# Diagnosing early-onset neonatal sepsis in low-resource settings: development of a multivariable prediction model

**DOI:** 10.1101/2022.11.19.22282335

**Authors:** Samuel R. Neal, Felicity Fitzgerald, Simbarashe Chimhuya, Michelle Heys, Mario Cortina-Borja, Gwendoline Chimhini

## Abstract

**Objective:** To develop a clinical prediction model to diagnose neonatal sepsis in low-resource settings.

**Design:** Secondary analysis of data collected by the Neotree digital health system from 01/02/2019 to 31/03/2020. We used multivariable logistic regression with candidate predictors identified from expert opinion and literature review. Missing data were imputed using multivariate imputation and model performance was evaluated in the derivation cohort.

**Setting:** A tertiary neonatal unit at Sally Mugabe Central Hospital, Zimbabwe.

**Patients:** We included 2628 neonates aged <72 hours, gestation ≥ 32^+0^ weeks and birth weight ≥ 1500 grams.

**Interventions:** Participants received standard care as no specific interventions were dictated by the study protocol.

**Main outcome measures:** Clinical early-onset neonatal sepsis (within the first 72 hours of life), defined by the treating consultant neonatologist.

**Results:** Clinical early-onset sepsis was diagnosed in 297 neonates (11.3%). The optimal model included eight predictors: maternal fever, offensive liquor, prolonged rupture of membranes, neonatal temperature, respiratory rate, activity, chest retractions and grunting. Receiver operating characteristic analysis gave an area under the curve of 0.736 (95% confidence interval 0.701-0.772). For a sensitivity of 95% (92-97%), corresponding specificity was 11% (10-13%), positive predictive value 12% (11-13%), negative predictive value 95% (92-97%), positive likelihood ratio 1.1 (95% CI 1.0-1.1), and negative likelihood ratio 0.4 (95% CI 0.3-0.6).

**Conclusions:** Our clinical prediction model achieved high sensitivity with modest specificity, suggesting it may be suited to excluding early-onset sepsis. Future work will validate and refine this model before considering it for clinical use within the Neotree.

**What is already known on this topic:** Various clinical prediction models exist to diagnose neonatal sepsis. However, there is a paucity of literature on models in low-resource settings, particularly sub-Saharan Africa.

**What this study adds:** We developed a clinical prediction model to diagnose clinical early-onset neonatal sepsis with over 2,500 neonates in a lower middle-income, low-resource neonatal unit. Our model is easy to implement, does not require laboratory tests and achieved high sensitivity with modest specificity in the derivation cohort.

**How this study might affect research, practice or policy:** Our model could support less experienced healthcare professionals avoid unnecessary antibiotic therapy in the absence of immediate local senior support. Before implementation, this model must be externally validated and its impact on sepsis-related neonatal morbidity and mortality must be assessed in future studies.

## INTRODUCTION

Neonatal sepsis caused 15% of the 2.5 million neonatal deaths worldwide in 2018 and has a mortality rate of 110-190 per 1000 livebirths.[1, 2] It can be difficult to diagnose as the clinical features overlap with non-infectious diseases.[3] Failing to treat sepsis with timely antimicrobials increases the risk of death or disability, but empirical antimicrobial therapy in non-infected neonates contributes to antimicrobial resistance and adverse outcomes.[4, 5]

Isolating a pathogenic organism from a normally sterile site is the gold standard diagnostic method,[6] but has limitations. In low-resource settings (LRS), cultures and blood counts are often unavailable,[7] or turnaround times are too long to usefully inform management.[8, 9] Blood cultures have high sensitivity provided sufficient inoculate volume is obtained, but sampling can be difficult in unwell neonates.[10] Therefore, clinicians may diagnose sepsis and initiate empirical antimicrobial therapy despite negative cultures, based on clinical presentation, risk factors and/or raised inflammatory markers. This is often called ‘culture-negative’ sepsis and up to 16 times more neonates receive antibiotics for culture-negative sepsis than for sepsis with a positive culture.[11] Diagnostic challenges are increased in LRS where early neonatal care may be led by less experienced healthcare professionals (HCPs) without immediate local senior support.[8]

Clinical prediction models combine patient or disease characteristics to estimate the probability of a diagnosis or outcome.[12] Models to diagnose neonatal sepsis may improve diagnostic accuracy and rationalise antibiotic use. In LRS, they could provide clinical decision support for less experienced HCPs, especially if models do not require laboratory tests. Several existing models estimate the probability of neonatal sepsis,[13] for example, the Kaiser Permanente Early-Onset Sepsis Calculator.[14] Unfortunately, few studies have investigated models in LRS, particularly in sub-Saharan Africa.[13]

Our primary objective was to develop a clinical prediction model to diagnose neonatal sepsis in a LRS neonatal unit, with the aim to support less experienced HCPs make this diagnosis.

## METHODS

We describe methods according to the TRIPOD statement (Additional File 1).[15] Further methods are in Additional File 2 and accompanying R code at https://doi.org/10.5281/zenodo.6969912.

### Source of data

We performed secondary analysis of data from the Neotree at the neonatal unit of Sally Mugabe Central Hospital (SMCH), Zimbabwe. Data were collected over 14 months from 01/02/2019 to 31/03/2020.

The Neotree is an open-source digital health system for newborn care in LRS.[16, 17] It combines evidence-based clinical decision support, education and digital data capture at admission and discharge. It is embedded in routine practice at three neonatal units in sub-Saharan Africa (Kamuzu Central Hospital, Malawi; SMCH, Zimbabwe; and Chinhoyi Provincial Hospital, Zimbabwe).

On admission, HCPs complete an admission form using the Neotree application on an Android tablet. The application guides assessment of the neonate and collects predefined data. At discharge or after neonatal death, HCPs complete an outcome form, which includes the final diagnoses or cause(s) of death after review by a consultant neonatologist (Additional File 2, section 1).

### Participants

SMCH has the largest of three tertiary neonatal units in Zimbabwe, with 100 cots. It admits neonates born within the hospital and accepts national referrals for specialist surgical care.

We included neonates with chronological age <72 hours, ≥ 32^+0^ weeks gestation at birth, and birth weight ≥ 1500 grams. We excluded non-first-born multiples and those with a diagnosis of major congenital anomaly, no outcome form completed, or anomalous admission durations (e.g. date of discharge before date of admission).

### Outcome

The primary outcome was clinical early-onset neonatal sepsis (EOS), defined as sepsis with onset within the first 72 hours of life, as diagnosed by the treating consultant neonatologist and recorded on the outcome form as one or more of: (i) primary discharge diagnosis, (ii) additional problem during admission, (iii) primary cause of death, or (iv) contributory cause of death. No specific actions were performed to blind outcome assessment.

### Predictors

We identified candidate predictors through a modified Delphi method study,[18] and literature review.[13] We mapped these predictors to available Neotree data, yielding 22 candidate predictors (Additional File 2, section 2). No specific actions were performed to blind predictor assessment.

### Statistical analysis

Analyses were performed in RStudio version 2022.02.0+443 (R version 4.1.3).[19, 20] No specific sample size calculations were performed.

#### Data preparation

We linked admission and outcome forms using the Fellegi-Sunter framework of probabilistic record linkage (Additional File 2, section 4).[21, 22] We imputed missing values using multivariate imputation by chained equations.[23] Data were assumed to be missing at random and we created 40 imputed datasets (Additional File 2, section 6).

#### Model development and specification

We used multivariable logistic regression to predict diagnosis of clinical EOS. For convenience, model selection was performed in one dataset randomly selected from all imputed datasets. First, we fitted a ‘full’ main effects model containing all candidate predictors assuming linearity of continuous predictors and additivity at the predictor scale. We excluded categorical variables with skewed distributions (<5% category prevalence in either outcome group) if Fisher’s exact test was non-significant (*p* ≥ 0.05) for the *m* × *n* contingency table. Otherwise, skewed categorical predictors were retained, and smaller categories combined into an ‘other’ category. Next, we compared plausible variations to the full model, selecting the model which minimised both Akaike’s information criterion (AIC) and the Bayesian information criterion (BIC) as the ‘optimal’ model (Additional File 2, section 8). We explored non-linear effects of continuous predictors with natural cubic spline functions (2 to 10 degrees of freedom) and polynomial transformations (second-degree to fifth-degree polynomials). We tested for the biologically plausible interaction between birth weight and gestational age. Finally, we fitted the optimal model across all imputed datasets and obtained pooled regression coefficients and their standard errors (SEs) using Rubin’s rules.[24]

#### Model performance

We evaluated performance of the optimal model in the derivation cohort. Discrimination was quantified by plotting a receiver operating characteristic (ROC) curve in each imputed dataset. We pooled the area under the curve (AUC) and variance across imputed datasets using Rubin’s rules.[24] Discrimination was visualised with a box plot and density plot of the distributions of predicted probabilities for each observed outcome group (in the single dataset used for model selection). We calculated Yates’ discrimination slope as the absolute difference in mean predicted probabilities between the two observed outcome groups.[25] Sensitivity, specificity, predictive values, and likelihood ratios of the optimal model were estimated in the single dataset used for model selection. These metrics are presented for the ‘optimal’ probability threshold according to Youden’s *J* statistic,[26] and for thresholds corresponding to sensitivities of 80, 85, 90 and 95%. Confidence intervals for likelihood ratios and Yates’ discrimination slope were obtained using bootstrap with 10,000 resamples (basic method or normal approximation).[27]

### Research ethics approval

Research ethics approval was granted by the University College London Research Ethics Committee (16915/001, 5019/004), Medical Research Council Zimbabwe (MRCZ/A/2570), and Sally Mugabe Central Hospital Ethics Committee (250418/48). The ‘session ID’ numbers shown in Additional File 2 were generated at the time of data import and, thus, were not known to anyone outside of the research group.

## RESULTS

### Participants

Of the 3577 neonates with matched admission and outcome records, 2628 (73.5%) were included (Figure 1). Mean gestational age was 38.0 (SD = 2.5) weeks, mean birth weight 2889 (SD = 703) grams, and 221 (8.4%) neonates died (Table 1). In total, 297 had clinical EOS (11.3%, incidence 113 per 1000 admissions).

**Table 1.** Characteristics of the study participants.

| Characteristics | Overall | No sepsis | Sepsis | *p*-value |
| --- | --- | --- | --- | --- |
| *n* | 2628 | 2331 | 297 |  |
| Admission |  |  |  |  |
| Gestational age, weeks | 38.0 (2.5) | 38.0 (2.5) | 38.4 (2.3) | 0.005 |
| Birth weight, grams | 2889 (703) | 2881 (716) | 2950 (595) | 0.067 |
| Sex |  |  |  | 0.7 |
| Male | 1503 (57%) | 1338 (57%) | 165 (56%) |  |
| Female | 1122 (43%) | 990 (42%) | 132 (44%) |  |
| Unsure | 3 (0.1%) | 3 (0.1%) | 0 (0.0%) |  |
| Type of birth |  |  |  | 0.032 |
| Singleton | 2496 (95%) | 2205 (95%) | 291 (98%) |  |
| First-born twin | 127 (4.8%) | 121 (5.2%) | 6 (2.0%) |  |
| First-born triplet | 2 (<0.1%) | 2 (<0.1%) | 0 (0.0%) |  |
| Mode of delivery |  |  |  | 0.074 |
| SVD | 1889 (72%) | 1663 (71%) | 226 (76%) |  |
| Elective C-section | 136 (5.2%) | 124 (5.3%) | 12 (4.0%) |  |
| Emergency C-section | 561 (21%) | 510 (22%) | 51 (17%) |  |
| Instrumental | 42 (1.6%) | 34 (1.5%) | 8 (2.7%) |  |
| Postnatal age |  |  |  | <0.001 |
| < 2 hours of life | 1001 (38%) | 901 (39%) | 100 (34%) |  |
| 2-24 hours of life | 1257 (48%) | 1136 (49%) | 121 (41%) |  |
| 24-48 hours of life | 235 (9.0%) | 181 (7.8%) | 54 (18%) |  |
| 48-72 hours of life | 110 (4.2%) | 91 (3.9%) | 19 (6.5%) |  |
| Outcome |  |  |  |  |
| Admission duration | 2.3 [1.3-4.9] | 2.1 [1.2-4.1] | 6.0 [3.5-8.8] | <0.001 |
| Death | 221 (8.4%) | 184 (7.9%) | 37 (12%) | 0.008 |

Data are presented as mean (SD), *n* (%) or median [quartile 1 - quartile 3]. *P*-values are from Welch's two-sample *t*-test for gestational age and birth weight; the Wilcoxon-Mann-Whitney *U* test for admission duration; Pearson's chi-squared test for postnatal age at admission and death; and Fisher's exact test for sex, type of birth, and mode of delivery. C-section = caesarean section; SVD = spontaneous vaginal delivery.

**Figure 1.**
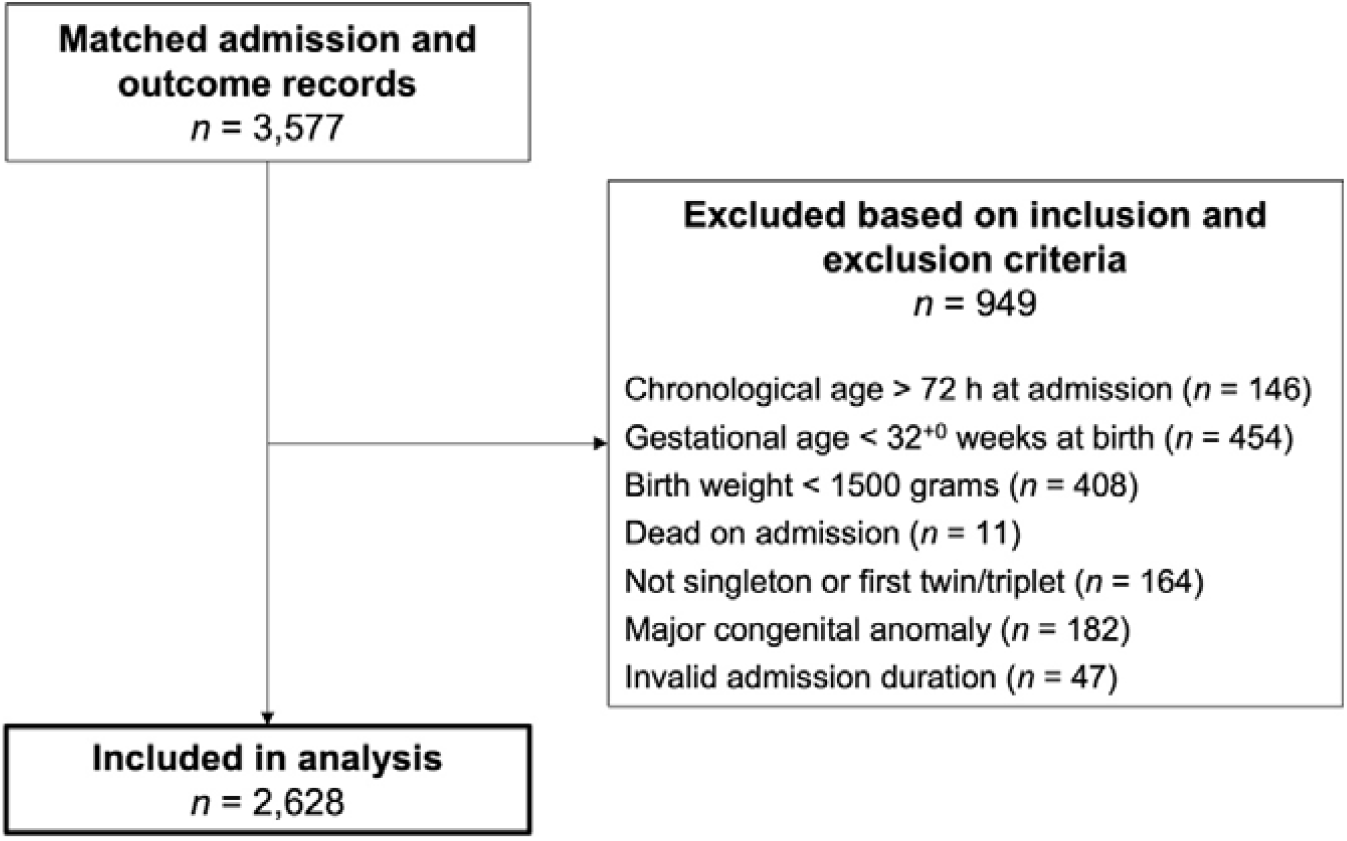
Flow diagram summarising participant inclusion and exclusion. Participants could fulfil multiple inclusion and/or exclusion criteria, therefore, the sum of participants excluded based on each criterion exceeds 949.

### Missing data

In total, 14 variables had missing values. All variables had <1% missing values except temperature (31%) and birth weight (1.2%). Time since the start of data collection predicted missing temperature (odds ratio [OR] 0.96, 95% CI 0.96-0.96, *p* < 0.001) as limited thermometers were available early in the study. Missing temperature was not associated with clinical EOS (OR 0.79, 95% CI 0.60-1.03, *p* = 0.084).

### Model development

From the set of 22 candidate predictors (Table 2), eight were excluded due to <5% category prevalence with a non-significant Fisher’s exact test (cyanosis, seizures, fontanelle, colour, abdominal distention, omphalitis, abnormal skin appearance, and history of vomiting). Three of the five categories for activity had a prevalence of <5% in either outcome group but Fisher’s exact test indicated a significant difference in the distribution between the two groups (*p* < 0.001). Activity was retained as a predictor and the three smaller categories were collapsed into one ‘other’ group.

**Table 2.** Distributions of candidate predictors in the study cohort.

| Candidate predictor | Overall | No sepsis | Sepsis | *p*-value |
| --- | --- | --- | --- | --- |
| *n* | 2628 | 2331 | 297 |  |
| Infant risk factors |  |  |  |  |
| Gestational age, weeks | 38.0 [37.0-40.0] | 38.0 [37.0-40.0] | 38.0 [37.0-40.0] | 0.032 |
| Birth weight, grams | 2950 [2400-3350] | 2900 [2400-3350] | 3000 [2600-3350] | 0.035 |
| Maternal risk factors |  |  |  |  |
| Maternal fever | 14 (0.5%) | 8 (0.3%) | 6 (2.0%) | 0.003 |
| Offensive liquor | 163 (6.2%) | 131 (5.6%) | 32 (11%) | 0.001 |
| PROM | 303 (12%) | 257 (11%) | 46 (15%) | 0.027 |
| Infant clinical features |  |  |  |  |
| Grunting at triage | 750 (29%) | 654 (28%) | 96 (32%) | 0.13 |
| Cyanosis at triage* | 69 (2.6%) | 60 (2.6%) | 9 (3.0%) | 0.6 |
| Seizures at triage* | 14 (0.5%) | 10 (0.4%) | 4 (1.3%) | 0.064 |
| Respiratory rate, breaths/min | 56 [48-68] | 56 [48-68] | 60 [50-72] | <0.001 |
| Heart rate, beats/min | 138 [126-146] | 138 [126-146] | 139 [127-150] | 0.011 |
| Temperature, Celsius | 36.5 [36.0-37.0] | 36.5 [36.0-36.9] | 36.9 [36.2-38.0] | <0.001 |
| Fontanelle* |  |  |  | 0.9 |
| Flat | 2,608 (99%) | 2,312 (99%) | 296 (100%) |  |
| Sunken | 10 (0.4%) | 9 (0.4%) | 1 (0.3%) |  |
| Bulging | 10 (0.4%) | 10 (0.4%) | 0 (0%) |  |
| Activity† |  |  |  | <0.001 |
| Alert | 2,152 (82%) | 1,933 (83%) | 219 (74%) |  |
| Lethargic | 382 (15%) | 327 (14%) | 55 (19%) |  |
| Irritable | 62 (2.4%) | 45 (1.9%) | 17 (5.7%) |  |
| Seizures | 14 (0.5%) | 9 (0.4%) | 5 (1.7%) |  |
| Coma | 18 (0.7%) | 17 (0.7%) | 1 (0.3%) |  |
| Nasal flaring | 912 (35%) | 791 (34%) | 121 (41%) | 0.023 |
| Chest retractions | 986 (38%) | 848 (36%) | 138 (46%) | <0.001 |
| Grunting | 421 (16%) | 360 (15%) | 61 (21%) | 0.029 |
| Work of breathing |  |  |  | <0.001 |
| Normal | 1,405 (54%) | 1,263 (55%) | 142 (48%) |  |
| Mildly increased | 413 (16%) | 378 (16%) | 35 (12%) |  |
| Moderately increased | 614 (24%) | 529 (23%) | 85 (29%) |  |
| Severely increased | 170 (6.5%) | 139 (6.0%) | 31 (11%) |  |
| Colour* |  |  |  | 0.11 |
| Pink | 2,507 (95%) | 2,220 (95%) | 287 (97%) |  |
| Pale | 10 (0.4%) | 7 (0.3%) | 3 (1.0%) |  |
| Blue | 62 (2.4%) | 58 (2.5%) | 4 (1.3%) |  |
| Yellow | 49 (1.9%) | 46 (2.0%) | 3 (1.0%) |  |
| Abdominal distention* | 28 (1.1%) | 26 (1.1%) | 2 (0.7%) | 0.8 |
| Omphalitis* | 6 (0.2%) | 4 (0.2%) | 2 (0.7%) | 0.14 |
| Abnormal skin* | 27 (1.0%) | 23 (1.0%) | 4 (1.3%) | 0.5 |
| Vomiting* |  |  |  | 0.3 |
| No | 2,605 (99%) | 2,309 (99%) | 296 (100%) |  |
| Yellow | 7 (0.3%) | 7 (0.3%) | 0 (0%) |  |
| Bilious | 13 (0.5%) | 13 (0.6%) | 0 (0%) |  |
| Blood-stained | 3 (0.1%) | 2 (<0.1%) | 1 (0.3%) |  |

\*Eliminated from the final set of candidate predictors due to very skewed distributions. †The three smallest categories of activity were collapsed into one ‘other’ category for model development. Data are presented as median [quartile 1 - quartile 3] for continuous predictors or *n* (%) for categorical predictors. *P*-values are from the Wilcoxon-Mann-Whitney *U* test for continuous predictors and Fisher’s exact test for categorical predictors. Distributions are presented for the observed data only, before multiple imputation of missing values.

Therefore, 14 candidate predictors were considered for model development. Of these, 12 had a significant univariable association with clinical EOS (Table 3). The strongest univariable predictor was maternal fever (OR 5.99, 95% CI 2.06-17.4). Neither birth weight (OR 1.14, 95% CI 0.96-1.35) nor grunting at triage (OR 1.23, 95% CI 0.95-1.59) predicted clinical EOS in univariable models.

**Table 3.**
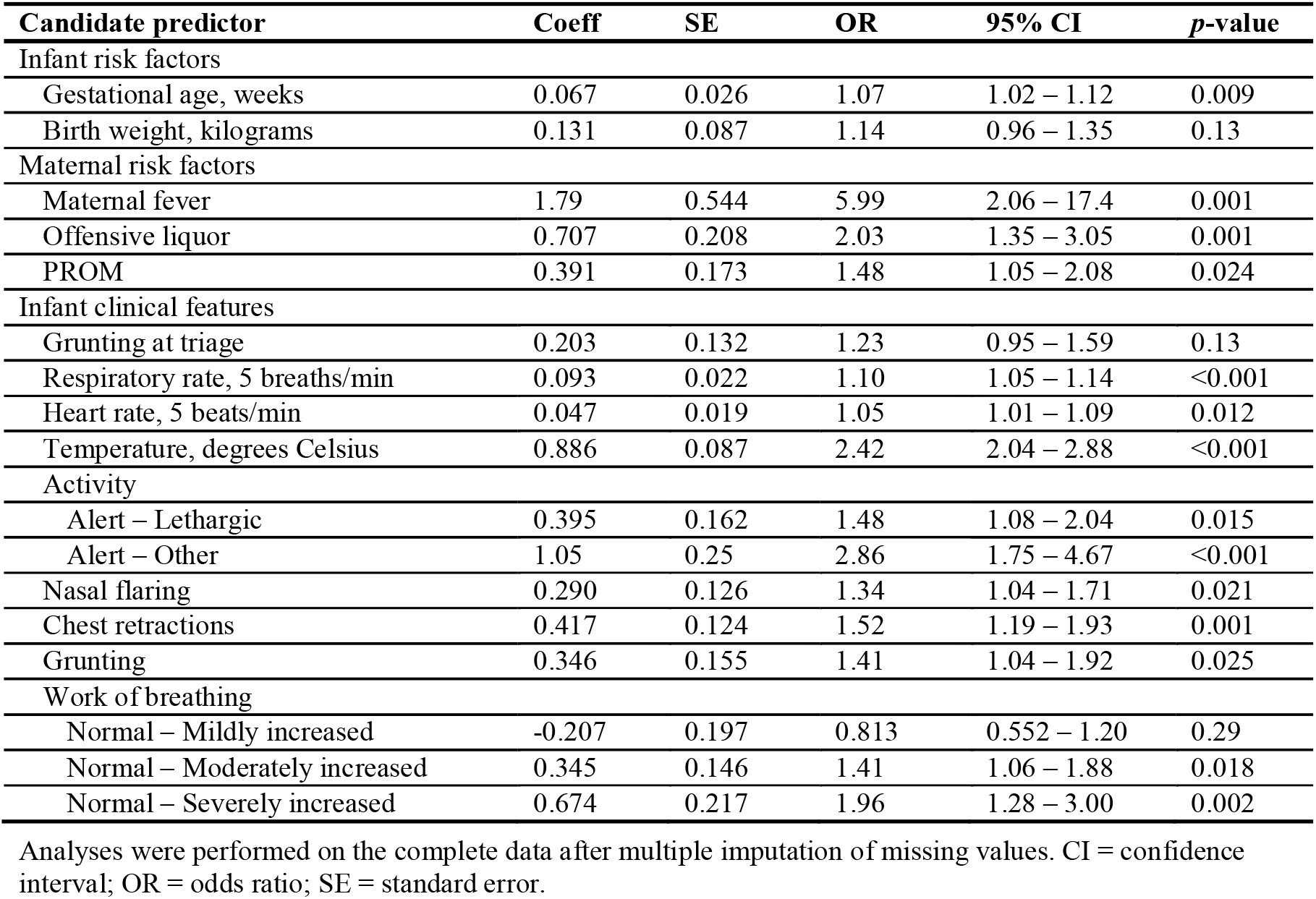
Univariable association between candidate predictors and outcome.

| **Candidate predictor** | **Coeff** | **SE** | **OR** | **95% CI** | **p-value** |
| --- | --- | --- | --- | --- | --- |
| Infant risk factors |  |  |  |  |  |
| Gestational age, weeks | 0.067 | 0.026 | 1.07 | 1.02 – 1.12 | 0.009 |
| Birth weight, kilograms | 0.131 | 0.087 | 1.14 | 0.96 – 1.35 | 0.13 |
| Maternal risk factors |  |  |  |  |  |
| Maternal fever | 1.79 | 0.544 | 5.99 | 2.06 – 17.4 | 0.001 |
| Offensive liquor | 0.707 | 0.208 | 2.03 | 1.35 – 3.05 | 0.001 |
| PROM | 0.391 | 0.173 | 1.48 | 1.05 – 2.08 | 0.024 |
| Infant clinical features |  |  |  |  |  |
| Grunting at triage | 0.203 | 0.132 | 1.23 | 0.95 – 1.59 | 0.13 |
| Respiratory rate, 5 breaths/min | 0.093 | 0.022 | 1.10 | 1.05 – 1.14 | <0.001 |
| Heart rate, 5 beats/min | 0.047 | 0.019 | 1.05 | 1.01 – 1.09 | 0.012 |
| Temperature, degrees Celsius | 0.886 | 0.087 | 2.42 | 2.04 – 2.88 | <0.001 |
| Activity |  |  |  |  |  |
| Alert – Lethargic | 0.395 | 0.162 | 1.48 | 1.08 – 2.04 | 0.015 |
| Alert – Other | 1.05 | 0.25 | 2.86 | 1.75 – 4.67 | <0.001 |
| Nasal flaring | 0.290 | 0.126 | 1.34 | 1.04 – 1.71 | 0.021 |
| Chest retractions | 0.417 | 0.124 | 1.52 | 1.19 – 1.93 | 0.001 |
| Grunting | 0.346 | 0.155 | 1.41 | 1.04 – 1.92 | 0.025 |
| Work of breathing |  |  |  |  |  |
| Normal – Mildly increased | -0.207 | 0.197 | 0.813 | 0.552 – 1.20 | 0.29 |
| Normal – Moderately increased | 0.345 | 0.146 | 1.41 | 1.06 – 1.88 | 0.018 |
| Normal – Severely increased | 0.674 | 0.217 | 1.96 | 1.28 – 3.00 | 0.002 |

Analyses were performed on the complete data after multiple imputation of missing values. CI = confidence interval; OR = odds ratio; SE = standard error.

Among plausible multivariable models, a model containing eight of the 14 candidate predictors was selected as the optimal model (Additional File 2, section 8). Fitting non-linear effects for temperature or birth weight, or allowing for an interaction between birth weight and gestational age, did not improve fit.

### Model specification

The optimal model included eight predictors: temperature at admission, respiratory rate, maternal fever during labour, offensive liquor, premature rupture of membranes, activity, chest retractions, and grunting (Table 4). It can be written as

**Table 4.** Predictors and their pooled regression coefficients and odds ratios for the optimal model.

| **Candidate predictor** | **Coeff** | **SE** | **OR** | **95% CI** | **p-value** |
| --- | --- | --- | --- | --- | --- |
| *Intercept* | -39.4 | 3.52 |  |  |  |
| Temperature, degrees Celsius | 0.987 | 0.095 | 2.68 | 2.23 – 3.23 | <0.001 |
| Respiratory rate, 5 breaths/min | 0.055 | 0.026 | 1.06 | 1.00 – 1.11 | 0.037 |
| Maternal fever | 1.44 | 0.612 | 4.21 | 1.27 – 14.0 | 0.019 |
| Offensive liquor | 0.543 | 0.228 | 1.72 | 1.10 – 2.69 | 0.017 |
| PROM | 0.360 | 0.192 | 1.43 | 0.98 – 2.09 | 0.06 |
| Activity (Alert – Lethargic) | 0.586 | 0.184 | 1.80 | 1.25 – 2.58 | 0.002 |
| Activity (Alert – Other) | 0.840 | 0.286 | 2.32 | 1.32 – 4.06 | 0.003 |
| Chest retractions | 0.406 | 0.172 | 1.50 | 1.07 – 2.10 | 0.019 |
| Grunting | 0.179 | 0.186 | 1.20 | 0.83 – 1.72 | 0.3 |

Analyses were performed on the complete data after multiple imputation of missing values. CI = confidence interval; OR = odds ratio; SE = standard error.

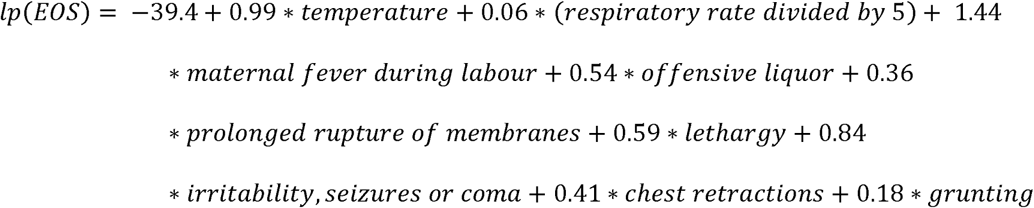

where *lp(EOS)* is the logit transformation of the probability of clinical EOS. The probability of clinical EOS (*Pr(EOS)*) is thus given by the inverse-logit function

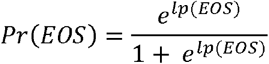

### Model performance

The pooled AUC was 0.736 (95% CI 0.701-0.772) (Figure 2). Median predicted probability was higher for observed cases with EOS than without EOS but there was significant overlap (Figure 3). Yates’ discrimination slope was 0.11 (95% CI 0.064-0.13).

**Figure 2.**
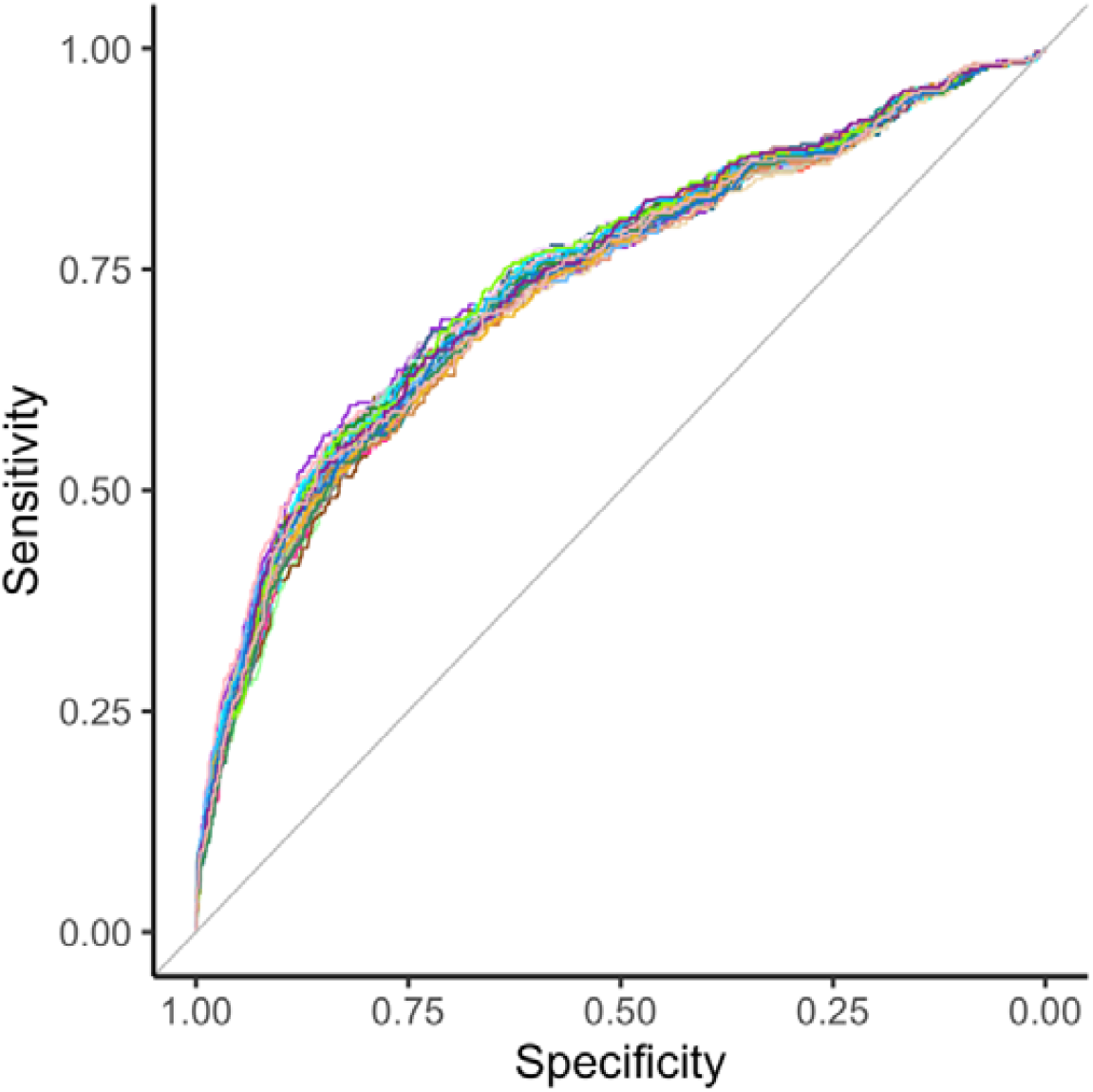
Receiver operating characteristic curve for the optimal model in each of the 40 imputed datasets. Pooled area under the curve (AUC) = 0.736 (95% confidence interval 0.701 – 0.772).

**Figure 3.**
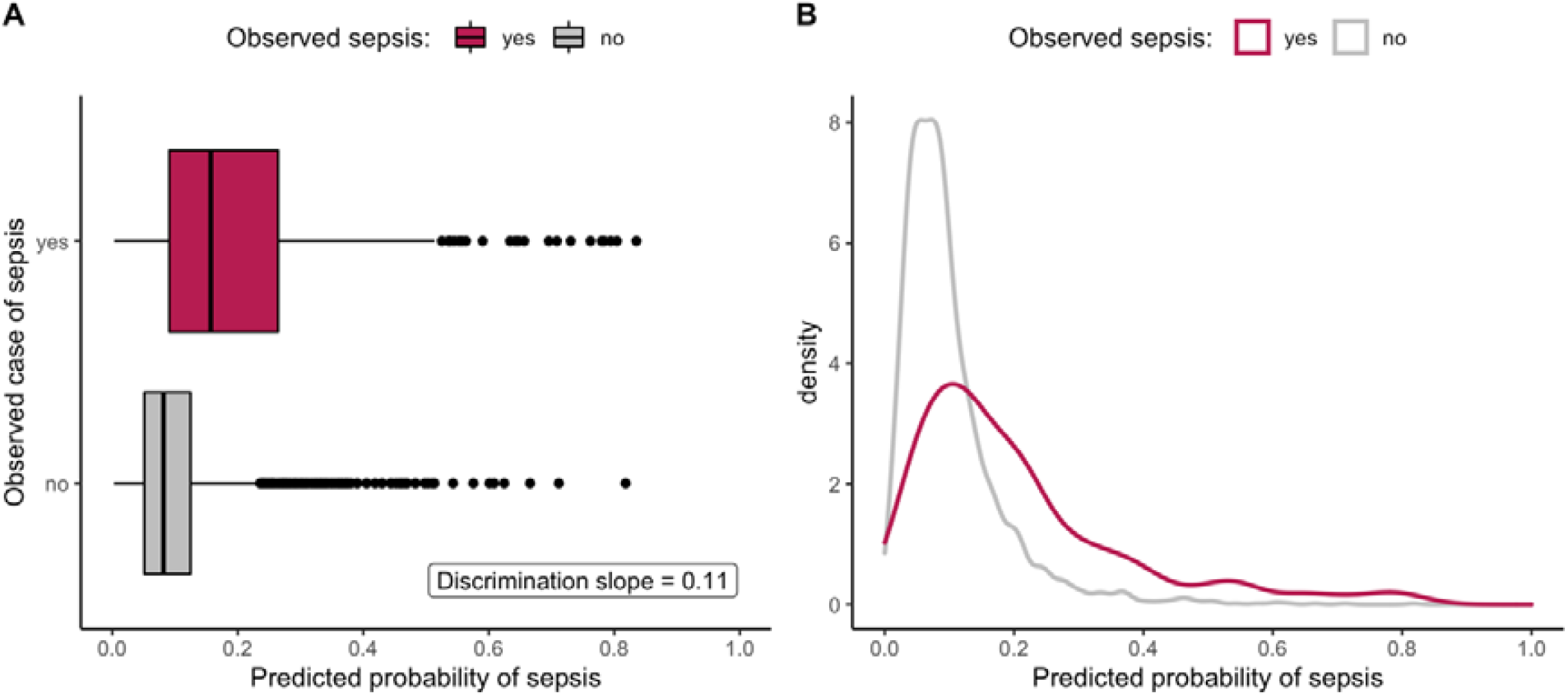
Boxplot (panel A) and density plot (panel B) of predicted probabilities of early-onset sepsis by observed outcome for the optimal model in the single dataset used for model selection.

The ‘optimal’ classification threshold was 0.121 (i.e. 12.1% predicted probability of clinical EOS) yielding sensitivity 65% (95% CI 59-70%) and specificity 74% (95% CI 72-75%) (Table 5). For a sensitivity of 95%, the corresponding classification threshold was 0.034 giving sensitivity 95% (95% CI 92-97%) and specificity 11% (95% CI 10-13%) (Table 5).

**Table 5.** Model performance at several classification thresholds of predicted probability.

| Threshold | Sens | Spec | PPV | NPV | LR+ | LR- |
| --- | --- | --- | --- | --- | --- | --- |
| 0.121* | 65 (59 – 70) | 74 (72 – 75) | 24 (21 – 27) | 94 (93 – 95) | 2.4 (1.6 – 2.9) | 0.5 (0.4 – 0.6) |
| 0.075 | 81 (76 – 85) | 44 (42 – 46) | 15 (14 – 17) | 95 (93 – 96) | 1.4 (1.0 – 1.6) | 0.4 (0.4 – 0.5) |
| 0.067 | 85 (80 – 88) | 38 (36 – 40) | 15 (13 – 17) | 95 (94 – 96) | 1.4 (1.2 – 1.6) | 0.4 (0.2 – 0.5) |
| 0.047 | 90 (86 – 93) | 22 (20 – 24) | 13 (12 – 14) | 95 (92 – 96) | 1.2 (0.9 – 1.2) | 0.4 (0.3 – 0.6) |
| 0.034 | 95 (92 – 97) | 11 (10 – 13) | 12 (11 – 13) | 95 (92 – 97) | 1.1 (1.0 – 1.1) | 0.4 (0.3 – 0.6) |

\*The ‘optimal’ threshold according to Youden’s *J* statistic. Data are presented for the single dataset used for model selection. Numbers in brackets represent the 95% confidence intervals. LR+ = positive likelihood ratio; LR- = negative likelihood ratio; PPV = positive predictive value; NPV = negative predictive value; sens = sensitivity; spec = specificity.

## DISCUSSION

We developed a clinical prediction model to diagnose clinical EOS that can be applied in LRS. The optimal model included eight predictors: three perinatal risk factors (maternal fever during labour, offensive liquor, and prolonged rupture of membranes), and five clinical signs in the neonate (temperature, respiratory rate, activity on neurological examination, chest retractions, and grunting). Using a classification threshold for high sensitivity, this model had a relatively low specificity in the derivation cohort.

### Interpretation

Incidence of clinical EOS in this cohort was 113 per 1000 admissions. This is greater than other estimates from low-income and middle-income countries, but there is marked heterogeneity between relatively few studies worldwide. For example, a 2019 meta-analysis estimated global EOS incidence of 31.1 per 1000 livebirths (95% CI 8.98-102.22; *I*^2^ 99.9%).[28]

Our model shares predictors with existing models for neonatal sepsis.[13] However, it does not need results from laboratory tests so is more applicable to LRS. Models exist for EOS that do not require laboratory tests (some of which have been validated in LRS), but data are limited to a few small studies. For example, Weber et al. developed a score with 14 clinical features to predict neonatal sepsis, meningitis, pneumonia or hypoxemia in neonates presenting to health facilities in four LRS countries.[29] Validation in the subgroup of 285 neonates aged ≤6 days of life showed a sensitivity of 95% with a specificity of 26% if one or more clinical features were present.[29] Comparison of performance between models is challenging as studies infrequently report readily comparable metrics (e.g. AUC) and often present performance at selected probability thresholds.

The Kaiser Permanente Early-Onset Sepsis Calculator has gained interest for managing neonates born at ≥ 34 weeks’ gestation.[30, 31] It combines maternal perinatal risk factors with the neonate’s clinical appearance to provide management recommendations based on the estimated probability of EOS. Meta-analyses suggest its use reduces rates of admission, antibiotic use and use of laboratory tests, without increased mortality (although some authors have voiced concerns about ‘missed’ or delayed diagnoses).[32–35] All included studies in these meta-analyses were performed in high-income countries.

The Kaiser Permanente calculator does not require results of laboratory tests but may be ill-suited to LRS. First, the baseline incidences of EOS used are lower than in most LRS: 0.1-4.0 per 1000 livebirths for the calculator,[30] compared to 31.1 per 1000 livebirths (95% CI 8.98-102.22) in LRS.[28] Second, the calculator was developed in a population where Group B streptococcus (GBS) is the predominant organism in EOS and where antenatal GBS screening is performed routinely. The microbiology of EOS differs in LRS. *Staphylococcus aureus, Klebsiella* species and *Escherichia coli* are relatively more common isolates than GBS.[36, 37] Therefore, risk factors such as maternal GBS carriage and intrapartum antibiotic status are less relevant or unmeasured in LRS. Finally, descriptors used for categories of clinical presentation (“clinical illness”, “equivocal” and “well appearing”) include interventions such as mechanical ventilation, which are not useful measures of illness in neonatal units where these interventions are unavailable. Two studies have validated the Kaiser Permanente calculator in middle-income countries with variable results.[38, 39] No studies have validated the calculator in low-income countries or sub-Saharan Africa.

### Implications

Our model is easy to apply in LRS. It includes clinical predictors and risk factors that are simple to identify by any cadre of HCP with minimal additional training. Acceptable thresholds of sensitivity and specificity will vary by clinical context. High sensitivity is important to avoid missing a true case of sepsis, but higher specificity would reduce risks associated with inappropriate antimicrobial therapy. During resource shortages, lower sensitivity might be favoured for higher specificity to allow treatment of neonates with the highest probability of EOS.

The high sensitivity with modest specificity of our model suggests it may be suited to excluding EOS. Approximately 300 neonates are admitted each month to SMCH.[40] At a sensitivity of 95% with our EOS incidence of 113 per 1000 admissions, we would expect one or two true cases of EOS to be missed per month. The corresponding negative predictive value of our model at this sensitivity was 95% (95% CI 92-97%). This might reassure HCPs antibiotics are not required.

### Limitations

First, the Neotree collects data at admission and upon discharge or death. This limits granular analysis where the timing of clinical features or interventions is important. We restricted our analysis to EOS and to neonates admitted within 72 hours of birth so the association between clinical presentation at admission and final diagnoses was clearer. It is plausible neonates admitted for ‘safekeeping’ (while their mother received inpatient care) could have unremarkable clinical appearance and vital signs on admission but develop symptoms of sepsis a few hours or days later.

Second, no specific actions were performed to blind assessment of the outcome. As we performed secondary analysis of data from a quality improvement project, the consultant neonatologist is unlikely to have been biased in their classification of EOS.

Third, although blood culture is the gold standard method for diagnosing EOS, erratic supplies of lab reagents meant we could not assess the correlation between positive blood cultures and the consultant neonatologists’ diagnosis of EOS.

Fourth, relatively small sample size caused imprecise estimates of the effects of low prevalence predictors: history of maternal fever had the largest effect size in our optimal model with OR 4.21 and wide 95% CI 1.27-14.0. Similarly, we could not evaluate the predictive ability of some identified candidate predictors; for example, only 10 neonates (0.4%) had a bulging fontanelle. Large sample sizes would be needed to determine if inclusion of these features is beneficial.

Finally, we present model performance in the derivation data to maximise sample size for model development. Predictions made on the derivation cohort can be optimistic due to overfitting.[12]

## Conclusions

Our prediction model to diagnose clinical EOS includes eight predictors: three perinatal risk factors (maternal fever during labour, offensive liquor, and prolonged rupture of membranes), and five clinical signs in the neonate (temperature, respiratory rate, activity on neurological examination, chest retractions, and grunting). With high sensitivity it achieved modest specificity in the derivation cohort suggesting it may be suited to excluding EOS, which could support HCPs’ decisions to withhold antibiotics in non-septic neonates. Our future work will include (1) validating and refining this model; (2) evaluating the acceptability and feasibility of implementing this model via the Neotree; and (3) evaluating the impact of implementing this model on sepsis-related neonatal morbidity and mortality.

## Data Availability

An open-source, anonymised research database is planned as part of the wider Neotree project. Currently, sharing of deidentified individual participant data will be considered on a case-by-case basis.

## Abbreviations

AIC: Akaike’s information criterion
AUC: area under the curve
BIC: Bayesian information criterion
CI: confidence interval
EOS: early-onset neonatal sepsis
GBS: Group B streptococcus
HCP: healthcare professional
IQR: interquartile range
LRS: low-resource settings
OR: odds ratio
ROC: receiver operating characteristic
SD: standard deviation
SE: standard error
SMCH: Sally Mugabe Central Hospital.

## ACKNOWLEDGMENTS

We thank Dr David Musorowegomo, Dr Hannah Gannon and Ms Heather Chesters for assistance with the literature review. We thank Dr Liam Shaw and Mr Yali Sassoon for technical assistance exporting and manipulating Neotree data. We thank the wider Neotree team including Dr Caroline Crehan and Mr Tim Hull-Bailey for valuable discussions throughout the study. We also thank all the staff in the neonatal unit at Sally Mugabe Central Hospital, especially Dr Christopher Pasi (Chief Executive Officer), Dr Hopewell Mungani (Clinical Director), Ms Prisca Nyamapfeni, Matron Alice Mudzingwa and Matron Dade Pedzisai for local support. Finally, we are grateful to all the babies and families who participated.

## AUTHORS’ CONTRIBUTIONS STATEMENT

Dr Samuel Neal designed the study protocol, conducted the literature review, carried out the analyses, drafted the initial manuscript, and reviewed and revised the manuscript.

Drs Michelle Heys and Felicity Fitzgerald conceptualised the study, led the implementation of Neotree in Zimbabwe, supervised the analyses, and critically reviewed and revised the manuscript.

Drs Gwendoline Chimhini and Simbarashe Chimhuya led the implementation of Neotree in Zimbabwe, provided the data, contributed to study conception, and critically reviewed and revised the manuscript.

Professor Mario Cortina-Borja supervised the analyses, and critically reviewed and revised the manuscript.

All authors approved the final manuscript as submitted and agree to be accountable for all aspects of the work.

