## Additional File 2 for "Diagnosing early-onset neonatal sepsis in low-resource settings: development of a multivariable prediction model"

Neal SR et al.

06/08/2022

#### Contents


**1 Neotree data collection** **2**

**2 Candidate predictors** **3**

**3 Preliminary data cleaning** **5**

**4 Record linkage** **8**

**5 Further data cleaning** **17**

**6 Missing data** **45**

**7 Descriptive statistics** **51**

**8 Model development and performance** **58**


Some supplementary data have been redacted or replaced by fictitious examples (where indicated) to remove identifying information for publication.

### 1 Neotree data collection

#### 1.1 Example screens from the Neotree app

Below are example screens from the Neotree app showing data capture and integrated education on neonatal care.

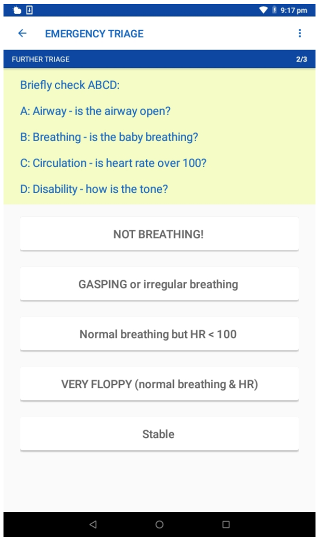


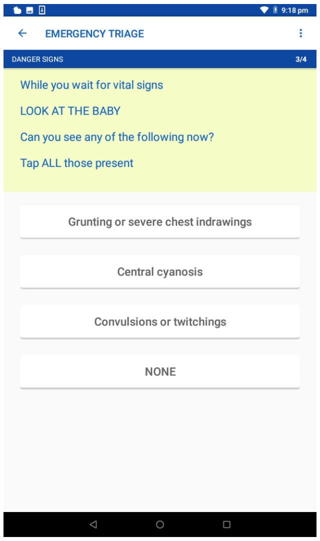


Note: A third screenshot has been redacted to comply with the medRxiv identifiable information policy. It showed a screen with an image of a neonate demonstrating neutral airway position and suction.

#### 1.2 Neotree data pipeline at Sally Mugabe Central Hospital

The below flow diagram summarises the current Neotree data pipeline.

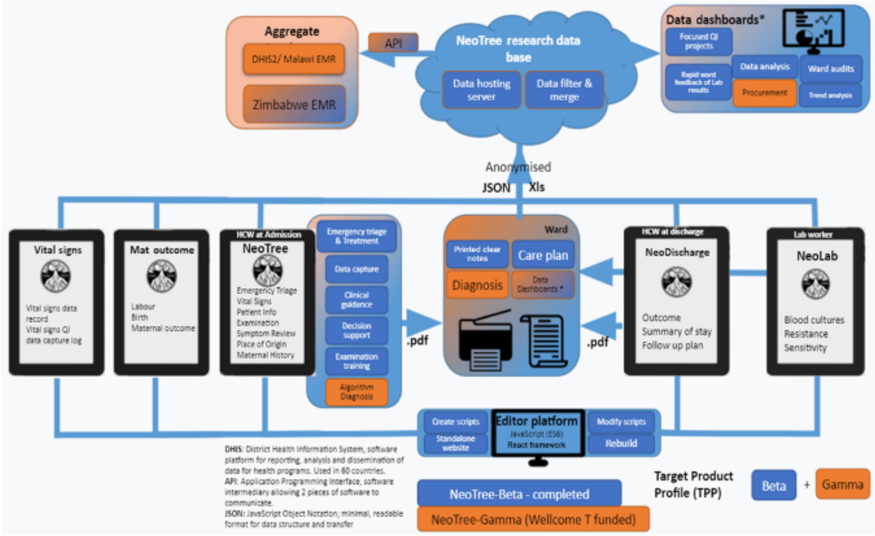


*Reprinted from Heys et al. 2022 (Learning Health Systems, DOI: 10.1002/lrh2.10310) in accordance with the CC BY 4.0 license.*

#### 2 Candidate predictors

| Predictors from literature review & expert opinion | Mapping to Neotree data (verbatim) |
|---|---|
| **Risk factors** | |
| Maternal fever >38°C in labor | "Tap all risk factors for sepsis present (some you have already asked about) these RFs will guide us on antibiotics" -> "Maternal fever in labour" |
| Prolonged rupture of membranes >18 hours | "When did the membranes rupture? (spontaneously or artificially)" -> "Did the membranes rupture?" -> "Yes" -> "How long between ROM and birth?" -> ">18 hours" |
| | "Tap all risk factors for sepsis present (some you have already asked about) these RFs will guide us on antibiotics" -> "PROM more than 18 hrs" |
| Foul smelling amniotic fluid | "Tap all risk factors for sepsis present (some you have already asked about) these RFs will guide us on antibiotics" -> "Offensive Liquor" |
| Gestation <32 weeks | "Gestation of the baby to the nearest week?" -> "Gestational age at birth (weeks)" -> *weeks (integer)* |
| Birth weight <1500g | "Look for birth weight in the obstetric record; Infants >24hrs old need a weight on the day of admission" -> "Birth Weight (g)" / "Admission Weight (g) (if different)" -> *grams (integer)* |
| **Signs and symptoms** | |
| Neonatal temperature >37.5°C | "Temperature (degs C)" -> *degrees Celsius (decimal, 1DP)* |
| Boil or abscess | "Examine the baby's skin" -> "Big Boil / Abscess" |
| Grunting, severe respiratory distress or moderate to severely increased work of breathing | "Look at the baby. Can you see any of the following now?" -> "Grunting or severe chest indrawings" |
| | "Tap all that are present (more than one if necessary)" -> "Nasal flaring" / "Chest in-drawings" / "Grunting" |
| | "How severe is the work of breathing" -> "Mild" / "Moderate" / "Severe" |
| Lethargy | "How is the baby's activity?" -> "Lethargic, quiet, decreased activity" |
| Umbilical redness or umbilicus draining pus | "Describe the umbilicus" -> "Red skin all around umbilicus" |
| Deep jaundice | "What colour is the baby?" -> "Yellow" |
| Tachypnoea >60 breaths per minute | "Tap the timer above to count the number of breaths in 30 seconds" -> *breaths per minute (integer)* |
| Convulsions, twitching or abnormal movements | "Look at the baby. Can you see any of the following now?" -> "Convulsions or twitchings" |
| | "How is the baby's activity?" -> "Seizures, convulsions, or twitchings" |
| Many or severe skin pustules | "Examine the baby's skin" -> "Pustules all over" |

| | |
|---|---|
| Bilious vomiting with severe abdominal distention | “Has the baby been vomiting” -> “Vomiting bright green” |
| | “Softly palpate the abdomen in all 4 quadrants” -> “Distended” |
| Bulging fontanelle | “Feel the fontanelle” -> “Bulging” |
| Not moving when stimulated | “How is the baby’s activity?” -> “Coma (unresponsive)” |
| Swollen red eyelids with pus | (No corresponding data collected by Neotree) |
| Central cyanosis | “Look at the baby. Can you see any of the following now?” -> “Central cyanosis” |
| | “What colour is the baby?” -> “Blue” |
| Pallor | “What colour is the baby?” -> “White” |
| Tachycardia >160 beats per minute | “Heart rate (beats/min)” -> *beats per minute (integer)* |

*DP = decimal place; PROM = prolonged rupture of membranes; RF = risk factor; ROM = rupture of membranes*

### 3 Preliminary data cleaning

We applied several preliminary cleaning steps to the raw imported data.

- Number of rows in raw admission data frame = 99468
- Number of rows in raw outcome data frame = 105139

#### 3.1 Removing duplicate entries

We defined exact duplicates as entries where values for all variables were identical to one or more other entries. This occurs when data are exported from a study tablet before previous data have been erased, resulting in some entries being exported in duplicate.

Number of duplicate entries:

```
## # A tibble: 2 x 2
##   form      duplicates
##   <chr>          <int>
## 1 admission      94801
## 2 outcome       100476
```

#### 3.2 Recoding missing values

We recoded empty cells or cells containing strings that signify missingness as missing values using the following custom function:

```
## function (x)
## {
##     strings <- c("", "na", "n/a", "N/A", "NA", "Nil", "nil",
##         "-")
##     x[x %in% strings] <- NA
##     x
## }
```

#### 3.3 Standardising variables between admission & outcome forms

We standardised `mode of delivery` and `sex` between admission and outcome forms, so they can be used for record linkage.

Labels before standardisation:

```
## $`Mode of delivery (admission)`
## [1] "1" "2" "3" "4" "5" "6"
##
## $`Mode of delivery (outcome)`
## [1] "ECS"  "ElCS" "For"  "SVD"  "Vent"
##
## $`Sex (admission)`
## [1] "F"  "M"  "NS"
##
## $`Sex (outcome)`
## [1] "F" "M" "U"
```

Labels after standardisation:

```
## $`Mode of delivery (admission)`
## [1] "ECS"  "ElCS" "For"  "SVD"  "Vent"
##
## $`Mode of delivery (outcome)`
## [1] "ECS"  "ElCS" "For"  "SVD"  "Vent"
##
## $`Sex (admission)`
## [1] "F" "M" "U"
##
## $`Sex (outcome)`
## [1] "F" "M" "U"
```

##### 3.4 Removing entries without a healthcare worker identifier

We removed entries that had not been 'signed off' by a healthcare worker with their healthcare worker identifier (HCW ID) (commonly their initials). Entries without a HCW ID occur for several reasons, e.g. (1) a healthcare worker accidentally exits the app and starts a new form upon reopening it; (2) a healthcare worker is demonstrating how to use the app to another user so does not want to mark the form as a genuine entry.

Number of entries without a HCW ID:

```
## # A tibble: 2 x 2
##   form      `no HCW ID`
##   <chr>           <int>
## 1 admission         100
## 2 outcome            88
```

##### 3.5 Removing outcome form entries with invalid unique identifiers

Invalid UIDs were:

```
## # A tibble: 4 x 2
##   format                                freq
##   <chr>                                <int>
## 1 missing values                          24
## 2 strings of only zeros                   84
## 3 strings shorter than 4 characters long    12
## 4 strings containing words                 3
```

##### 3.6 Limiting entries to the study period

We removed entries outwith the study period. This included entries prior to 01/02/2019, which constituted the 'pilot period' of data collection for the Neotree at SMCH.

Data import and preliminary cleaning resulted in one data frame for admission forms and one data frame for outcome forms.

- Number of rows in final admission data frame = 4137
- Number of rows in final outcome data frame = 3935

##### 3.7 Flow diagram

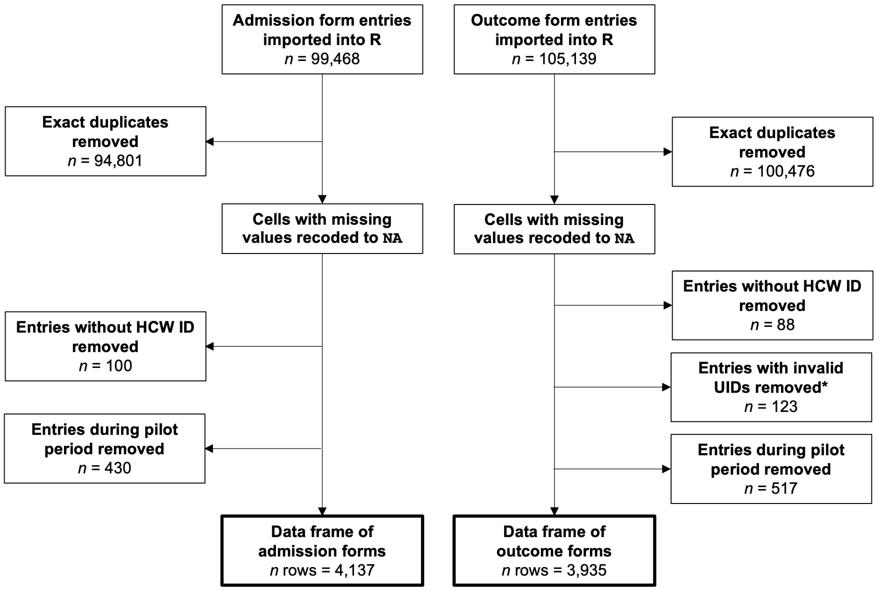


Figure 1: Flow diagram summarising preliminary data cleaning

### 4 Record linkage

At the time of our study, the Neotree app required users to manually enter the automatically generated admission unique identifier (UID) into a free-text field when completing the outcome form. Therefore, the outcome UID is liable to typographical errors and is not a 100% reliable key to link admission and outcome forms. Thus, we linked records using the Fellegi-Sunter framework of probabilistic record linkage.

#### 4.1 Create data frames for linkage

##### 4.1.1 Linkage variables

There are 8 variables common to both admission and outcome forms:

1. UID
2. Birth weight
3. Gestation at birth
4. Occipitofrontal circumference at admission
5. Length at admission
6. Mode of delivery
7. Sex
8. Place of birth

These have the following levels of missingness in the admission forms:

```
## # A tibble: 8 x 3
##   variable               n_miss pct_miss
##   <chr>                   <int>    <dbl>
## 1 Admission.PlaceBirth     3637  87.9
## 2 Admission.BW               68   1.64
## 3 Admission.OFC               5   0.121
## 4 Admission.Gestation         1   0.0242
## 5 Admission.Length            1   0.0242
## 6 Admission.UID_alphanum      0   0
## 7 Admission.ModeDelivery      0   0
## 8 Admission.Gender            0   0
```

And in the outcome forms:

```
## # A tibble: 8 x 3
##   variable                     n_miss pct_miss
##   <chr>                         <int>    <dbl>
## 1 Discharge.GestBirth            2499  63.5
## 2 Discharge.OFCDis                248   6.30
## 3 Discharge.LengthDis             231   5.87
## 4 Discharge.BirthPlace             13   0.330
## 5 Discharge.Delivery                1   0.0254
## 6 Discharge.NeoTreeID_alphanum      0   0
## 7 Discharge.BWTDis                  0   0
## 8 Discharge.SexDis                  0   0
```

Note `Place of birth` has 87.9% missing values in the admission forms. It is also coded differently between admission and outcome forms:

```
## $admission
## # A tibble: 5 x 2
##   levels definition
##   <chr>  <chr>
## 1 BBA    born before arrival
## 2 HC     health centre
## 3 Home   home
## 4 Hosp   hospital
## 5 TBA    traditional birth attendant
##
## $outcome
## # A tibble: 4 x 2
##   levels definition
##   <chr>  <chr>
## 1 H      home
## 2 HCH    Harare Central Hospital
## 3 OtH    other clinic in Harare
## 4 OtR    other clinic outside Harare
```

Although `Gestation at birth` has 63.5% missing values in the outcome forms, it is a numeric variable and, therefore, coded the same between admission and outcome forms:

```
## $admission
## [1] "40" "41" "33" "40" "40" "38"
##
## $discharge
## [1] "34" "32" "31" "27" "34" "34"
```

Thus, we used 7 variables for record linkage (excluding `Place of birth`):

1. Unique ID*
2. Birth weight
3. Gestation at birth
4. Occipitofrontal circumference at admission
5. Length at admission
6. Mode of delivery
7. Sex
8. ~~Place of birth~~

* **Special note on UID:** After the first month of the project, healthcare workers were told to only enter the first 3 and last 3 characters of the UID in the outcome form. This is because the UID was initially long and laborious to type and, hence, prone to error. Therefore, most outcome form UIDs are 6 characters long, except those from the first month of the project. To avoid confusion, we used a substring of the full UIDs (called `uidsub`) for linkage. This substring consists of the first 3 and last 3 characters of the UID, converted to lowercase. E.g. (fictitious example),

```
##      full       uidsub
## [1,] "AB123456" "ab1456"
## [2,] "AB789012" "ab7012"
```

##### 4.1.2 Linkage data frames

Below is a fictitious example of the data frame structure for record linkage:

```
## $admission
## # A tibble: 4 x 8
##   bw    gest  ofc   length mode  sex   session       uidsub
##   <chr> <chr> <chr> <chr>  <chr> <chr> <chr>         <chr>
## 1 3000  41    32    46     SVD   M     session 10000 ab1789
## 2 4000  40    37    46     ECS   F     session 10001 ab2567
## 3 1800  35    31    44     SVD   F     session 10002 ab3689
## 4 3500  40    33    48     SVD   M     session 10003 ab1478
##
## $outcome
## # A tibble: 4 x 8
##   bw    gest  ofc   length mode  sex   session        uidsub
##   <chr> <chr> <chr> <chr>  <chr> <chr> <chr>          <chr>
## 1 3320  <NA>  32    48     SVD   F     session 100000 cd3567
## 2 1900  32    32    47     ECS   F     session 100001 cd1378
## 3 1900  34    30    45     SVD   M     session 100002 cd8364
## 4 1300  32    29    39     ECS   M     session 100003 cd9246
```

#### 4.2 Perform record linkage

##### 4.2.1 Run linkage algorithm

We performed record linkage using the `fastLink` package by Enamorado, Fifield and Imai (https://github.com/kosukeimai/fastLink). Linkage is performed using the `fastLink::fastLink()` wrapper.

```r
set.seed(123)

matches_out <- fastLink(
  dfA = adm_link,
  dfB = dis_link,
  varnames = c("uidsub", "bw", "gest", "ofc", "length", "mode", "sex"),
  stringdist.match = c("uidsub"), # use string dist matching on uidsub
  stringdist.method = "jw", # Jaro-Winkler
  jw.weight = .10, # Jaro-Winkler weight for prefix
  partial.match = c("uidsub"), # allow partial matching for uidsub
  cut.a = 0.96, # full string-distance match cut point (Winkler, 1990)
  cut.p = 0.88, # partial string-distance match cut point (Winkler, 1990)
  dedupe.matches = TRUE, # enforces one-to-one matching
  cond.indep = TRUE, # assuming conditional independence for Fellegi-Sunter model
  return.all = TRUE # sets threshold.match to 0.0001
)
```

All other parameters were left as the default (see fastLink documentation).

##### 4.2.2 Determine thresholds

We plot the posterior probabilities (zeta) and their corresponding Fellegi-Sunter weights, as demonstrated by Weber. *Note that the y-axes values are displayed as the natural logarithm of 1 plus the number of records at each zeta or weight.*

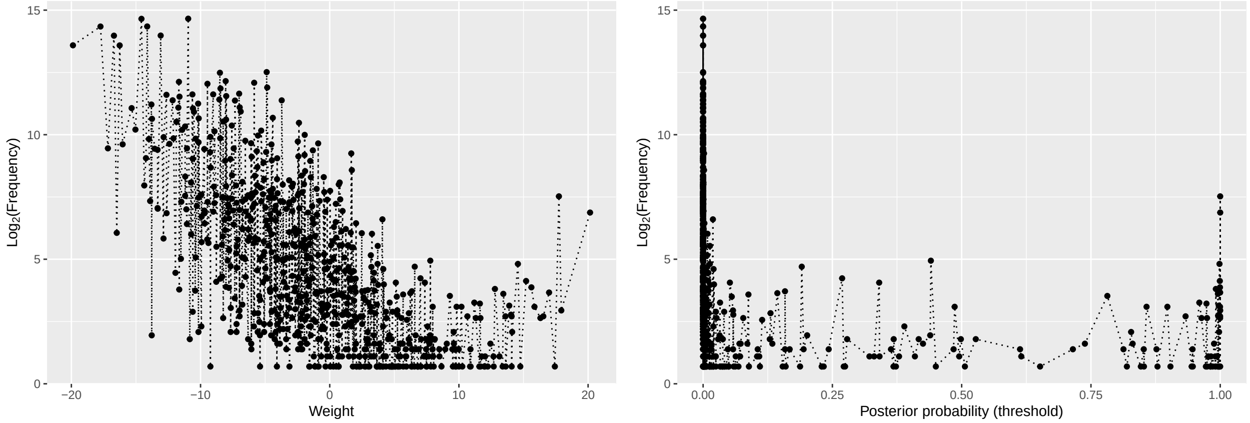


We then plot the frequencies of matches and non-matches, and the false positive rate (aka false detection rate [FDR]) and false negative rate (FNR) across the range of probability thresholds.

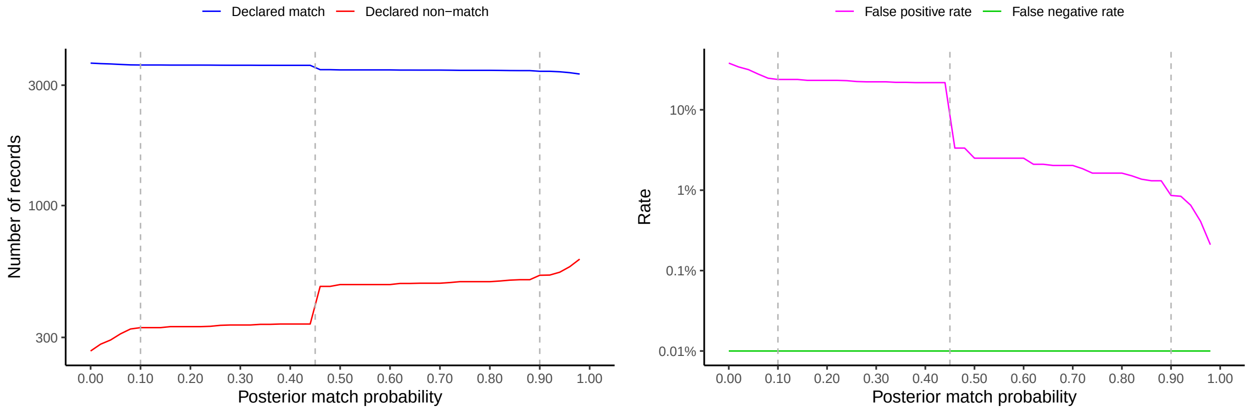


Considering the graphs, there appears to be an abrupt change in the number of matches vs. non-matches, and the FDR at zetas of ~0.10, ~0.45 and ~0.90 (*dotted lines*). The FNR appears essentially constant at 0.01% across all values of zeta.

It is most important to minimise the FDR (i.e. minimise the likelihood of declaring records a match when they are not a true match). Therefore, we set the threshold for declared matches very high (**zeta = 0.98**), which yielded an FDR <0.5%. We set the lower threshold (for declaring potential matches requiring manual review) at **zeta = 0.10**, based on the abrupt changes in the above graphs at this point.

- Zeta = 0.10 corresponds to Fellegi-Sunter weight = ~6.2.
- Zeta = 0.98 corresponds to Fellegi-Sunter weight = ~12.2.

Below are the zeta and Fellegi-Sunter weight plots with thresholds superimposed:

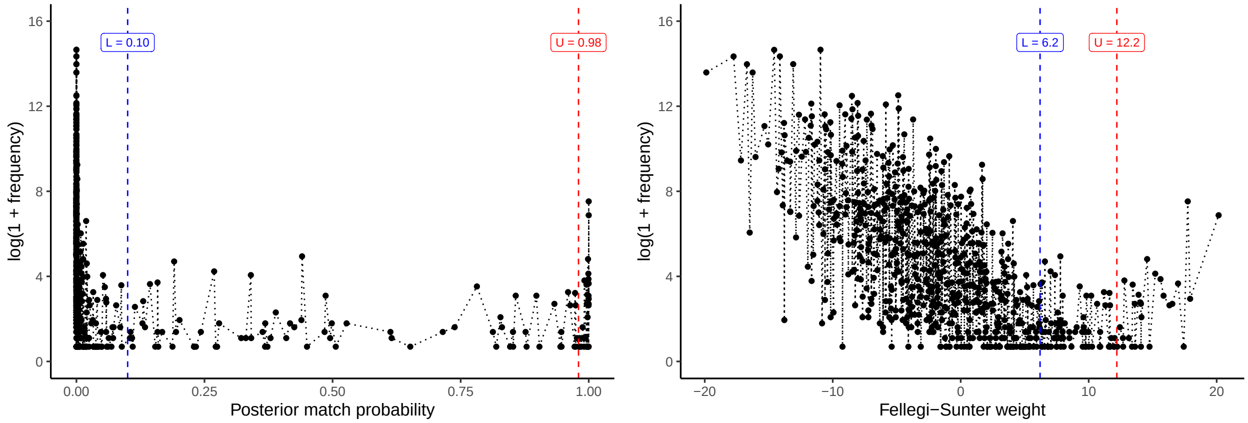


The chosen thresholds result in the following confusion tables:

```
## $lower_thres
## $lower_thres$confusion.table
##                        'True' Matches 'True' Non-Matches
## Declared Matches              3504.46             102.54
## Declared Non-Matches             0.27             327.73
##
## $lower_thres$addition.info
##                                results
## Max Number of Obs to be Matched 3935.00
## Sensitivity (%)                   99.99
## Specificity (%)                   76.17
## Positive Predicted Value (%)      97.16
## Negative Predicted Value (%)      99.92
## False Positive Rate (%)           23.83
## False Negative Rate (%)            0.01
## Correctly Classified (%)          97.39
## F1 Score (%)                      98.55
##
##
## $upper_thres
## $upper_thres$confusion.table
##                        'True' Matches 'True' Non-Matches
## Declared Matches              3320.70               1.30
## Declared Non-Matches             0.45             612.55
##
## $upper_thres$addition.info
##                                results
## Max Number of Obs to be Matched 3935.00
## Sensitivity (%)                   99.99
## Specificity (%)                   99.79
## Positive Predicted Value (%)      99.96
## Negative Predicted Value (%)      99.93
## False Positive Rate (%)            0.21
## False Negative Rate (%)            0.01
## Correctly Classified (%)          99.96
## F1 Score (%)                      99.97
```

This results in **285 records for manual review.** We deemed this to be an acceptable and pragmatic number of records to review manually.

###### 4.2.3 Get matches and potential matches at chosen thresholds

We subset the linkage data frames to return a data frame of matches and a data frame of potential matches using the `fastLink::getMatches()` function.

```
matches_list <- vector("list")

# With zeta >0.98 (matches)
matches_list$low <- getMatches(
  dfA = adm_link,
  dfB = dis_link,
  fl.out = matches_out,
  threshold.match = 0.98,
  combine.dfs = FALSE
)

# With zeta >0.10 (matches + potential matches)
matches_list$high <- getMatches(
  dfA = adm_link,
  dfB = dis_link,
  fl.out = matches_out,
  threshold.match = 0.10,
  combine.dfs = FALSE
)

# Session IDs for potential matches
matches_list$potential_adm <-
  matches_list$high$dfA.match$session[!matches_list$high$dfA.match$session
    %in% matches_list$low$dfA.match$session]

matches_list$potential_dis <-
  matches_list$high$dfB.match$session[!matches_list$high$dfB.match$session
                                       %in% matches_list$low$dfB.match$session]
```

We build this into a full data frame with all Neotree variables for matches and potential matches by merging on session ID (which uniquely identifies each completed admission or outcome form). N.B. `adm` and `dis` are the complete data frames of Neotree admission forms and outcome forms, respectively.

```
# Build into data frames
# Designated matches from fastLink
matches_list$matches <- tibble(
  Admission.session = matches_list$low$dfA.match$session,
  Discharge.session = matches_list$low$dfB.match$session
) %>%
  merge(adm, by = "Admission.session") %>%
  merge(dis, by = "Discharge.session")

# Potential matches from fastLink
matches_list$potentials <- tibble(
  Admission.session = matches_list$potential_adm,
  Discharge.session = matches_list$potential_dis
```

```
) %>%
  merge(adm, by = "Admission.session") %>%
  merge(dis, by = "Discharge.session")
```

There are **3322 declared matches, 285 declared potential matches, and 328 non-matches** from the Fellegi-Sunter linkage algorithm.

#### 4.3 Manual review of potential matches

Potential matches are manually reviewed to determine their true match status. We used several factors to make a clinical judgement, including:

- Admission and outcome UIDs - any discrepancies are plausible (e.g. likely to represent a typographical error).
- Admission date and outcome (discharge or death) date - congruent and plausible.
- Admission reason/diagnosis and discharge diagnosis or cause of death - congruent.
- A review of all other variables looking for any unique features on the admission and outcome form that might indicate a true match.

From manual review of the potential matches, we decided that **258** were true matches. Thus, there were **3580** declared matches at this stage.

##### 4.3.1 Quality checks

Finally, we performed several additional 'quality checks' to identify false-positive matches or other irregularities.

First, we checked for duplicate admission or outcome session IDs (i.e. duplicate completed admission or outcome forms) in the final linked dataset.

- Duplicated admission forms: $n = 0$
- Duplicated outcome forms: $n = 0$

Therefore, the one-to-one matching constraint was successful.

Next, we checked for duplicate admission or outcome UIDs in the final linked dataset.

- Duplicated admission UIDs: $n = 86$
- Duplicated outcome UIDs: $n = 108$

Looking at an extract from these duplicates, it was clear that they are unique babies with, for example, different birth weights, gestational ages, admission reasons, discharge diagnosis or outcome despite the same UID.

Finally, we checked for invalid admission durations (i.e. cases where the outcome date came before the admission date, or where the interval was unusually long).

Acceptable discrepancies:

- Outcome date $\leq$ 1 day prior to admission date - this could occur if the admission form was completed retrospectively shortly after the outcome form (e.g. if a baby was deceased on or soon after arrival to the neonatal unit).

Unacceptable discrepancies:

- Outcome date > 1 day prior to admission date
- Admission duration shown to be > 4 months - this is not a plausible admission duration for the neonatal unit at Sally Mugabe Central Hospital.

Distribution of admission durations:

```
##                           Min.                         1st Qu.
##          "-5d -21H -45M -2S"            "1d 6H 51M 52.25S"
##                         Median                            Mean
##              "2d 13H 2M 39S" "5d 6H 57M 15.7974860329414S"
##                        3rd Qu.                            Max.
##          "5d 14H 40M 59.25S"            "309d 14H 56M 13S"
```

- Outcome date prior to admission date: $n = 49$
  - $\leq$ 1 day prior: $n = 47$
  - $>$ 1 day prior: $n = 2$
- Admission duration shown to be >4 months: $n = 1$

We changed the status of these 3 cases to "non-match", to err on the side of caution. We felt these most likely represented false-positive matches.

```
## [1] "session 70565" "session 21083"
```

```
## [1] "session 10707"
```

New distribution of admission durations:

```
##                         Min.                         1st Qu.
##            "-16H -59M -25S"              "1d 6H 54M 10S"
##                       Median                            Mean
##             "2d 13H 7M 59S" "5d 5H 1M 47.4453452615999S"
##                      3rd Qu.                            Max.
##            "5d 14H 40M 46S"             "84d 22H 15M 49S"
```

**A total of 3577 record pairs were thus included in the final linked dataset.**

##### 4.3.2 Flow diagram

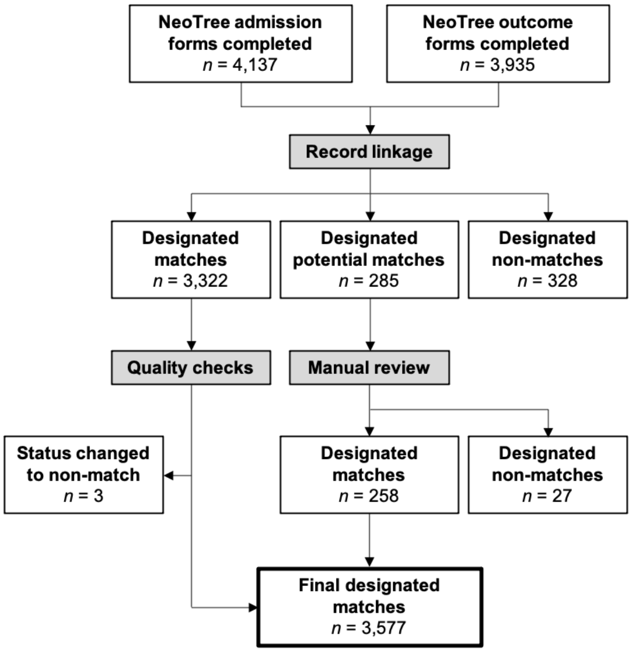


Figure 2: Flow diagram summarising record linkage

### 5 Further data cleaning

Elaboration on creating/extracting relevant variables required for model development from the Neotree dataset at Sally Mugabe Central Hospital.

#### 5.1 Data collected by admission forms

There are 7 sections to the Neotree admission form at SMCH, Zimbabwe.

1. Emergency triage & vital signs
2. Patient information
3. Examination
4. Symptom review
5. Place of origin
6. Maternal history
7. Provisional diagnoses

*N.B. Other data are collected by the app, but only relevant variables are detailed here.*

##### 5.1.1 Emergency triage & vital signs

The variables to be subset/created from this section are as follows:

| Parent variable | New variable(s) | Comments |
|---|---|---|
| Admission.DangerSigns | et_grunt | "Grun" (yes/no) |
| " " | et_cyanosis | "Cyan" (yes/no) |
| " " | et_seizures | "Conv" (yes/no) |
| Admission.RR | et_rr | (numeric) |
| Admission.HR | et_hr | (numeric) |
| Admission.Temperature | et_temp | (numeric) |
| Admission.BW | et_bw | (numeric) |
| Admission.AW | *informs* et_bw | (numeric) |

**5.1.1.1 Admission.DangerSigns** Categorical variable with four levels:

- Grun = "Grunting or severe chest indrawings"
- Cyan = "Central cyanosis"
- Conv = "Convulsions or twitchings"
- None

Recoded into three separate variables: `et_grunt`, `et_cyanosis` and `et_seizures`.

```
## [1] "Original variable"
```

```
##
##           Conv      Conv,Grun           Cyan           Grun Grun,Conv,Cyan
##             11              3             58           1067              1
##      Grun,Cyan           None           <NA>
##             82           2353              2
```

```
## [1] "New variables"
```

```
##  et_grunt    et_cyanosis et_seizures
##  no  :2422   no  :3434   no  :3560
##  yes :1153   yes : 141   yes :  15
##  NA's:   2   NA's:   2   NA's:   2
```

**5.1.1.2 Admission.RR** Continuous variable measured in breaths per minute.

- Some recorded values were very low (i.e. <20 breaths per minute).
  - On inspection, most died suggesting the recorded values were correct.
  - Some neonates were recorded as surviving to discharge with an initial RR $< 10$, despite receiving no resuscitation. This is implausible and their RR was set to missing.
- Similarly, some recorded values were very high (i.e. >100 breaths per minute).
  - After reviewing the distribution, we truncated these data to the 99.5th percentile, setting greater values to missing.

```
##    Min. 1st Qu.  Median    Mean 3rd Qu.    Max.    NA's
##     0.0    48.0    56.0    58.3    68.0   192.0       6
```

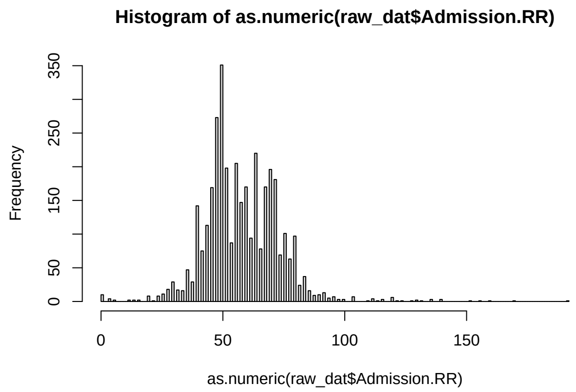


```
##    0%  0.5%    1%   50%   99% 99.5%  100%
##   0.0  13.7  24.0  56.0 104.0 120.0 192.0
```

```
## [1] "Lowest 10 values"
```

```
##
##  0  4  6 12 14 16 20 22 24 26
## 10  4  2  2  2  2  8  1  8 11
```

```
## [1] "Highest 20 values"
```

```
##
##  98 100 104 110 112 114 116 120 122 124 128 130 132 136 140 152 156 160 170 192
##   3   3   7   1   4   1   3   6   1   1   1   2   1   3   3   1   1   1   1   1
```

```
## [1] "Cases where RR <20"

## # A tibble: 22 x 3
##    Admission.RR Admission.Resus      Discharge.NeoTreeOutcome
##           <dbl> <chr>                <chr>
##  1           16 Stim,O2              NND
##  2            4 Stim,BVM,O2,Suc      NND
##  3            0 Stim,O2,Suc          NND
##  4            0 Stim,CPR,O2,BVM,Suc  NND
##  5            6 Stim,CPR,O2,BVM,Suc  NND
##  6           12 Stim,O2,BVM          NND
##  7            0 Stim,CPR,BVM         NND
##  8            6 None                 DC
##  9           16 Stim,O2              NND
## 10            0 CPR,Suc,O2,BVM       NND
## 11            0 Stim,CPR,O2,BVM,Suc  NND
## 12           12 Stim,BVM,O2,Suc      NND
## 13            0 Stim,BVM,O2,Suc      NND
## 14            4 None                 DC
## 15           14 Stim,CPR,O2,BVM,Suc  NND
## 16            4 Stim,CPR,O2,BVM,Suc  NND
## 17            0 None                 NND
## 18            0 Stim,CPR,O2,BVM,Suc  NND
## 19            0 Stim,CPR,BVM         NND
## 20            4 None                 DC
## 21           14 Stim,O2              DC
## 22            0 None                 NND

## [1] "New variable"

##      et_rr
##  Min.   :  0
##  1st Qu.: 48
##  Median : 56
##  Mean   : 58
##  3rd Qu.: 68
##  Max.   :120
##  NA's   :26
```

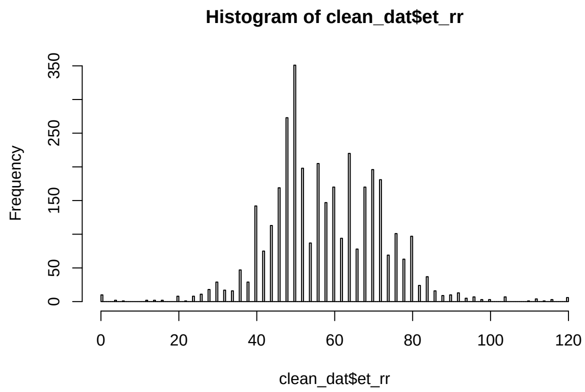

###### 5.1.1.3 Admission.HR

Continuous variable measured in beats per minute.

- Some recorded values were very low (i.e. <50 beats per minute).
  - On inspection, most died suggesting the recorded values were correct.
  - Some neonates were recorded as surviving to discharge with an initial HR < 20, despite receiving no resuscitation. This is implausible and their HR was set to missing.

```
##    Min. 1st Qu.  Median    Mean 3rd Qu.    Max.
##       0     125     138     136     146     252
```

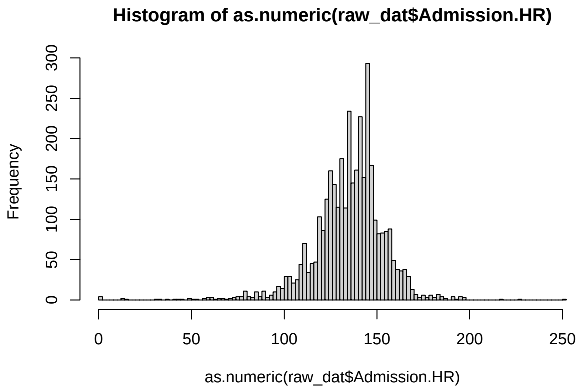


```
##      0%    0.1%      1%     50%     99%   99.9%    100%
##   0.000   8.064  74.760 138.000 179.000 198.000 252.000
```

```
## [1] "Lowest 10 values"
```

```
##
##  0 14 15 32 34 38 42 43 45 50
##  4  2  1  1  1  1  1  1  1  2
```

```
## [1] "Highest 20 values"
```

```
##
## 178 179 180 182 183 184 185 186 187 188 191 192 193 195 196 197 198 218 228 252
##   2   2   4   3   3   4   2   2   1   1   1   3   1   1   3   1   2   1   1   1
```

```
## [1] "Cases where HR <50"
```

```
## # A tibble: 13 x 4
##    Admission.HR Admission.RR Admission.Resus     Discharge.NeoTreeOutcome
##           <dbl> <chr>        <chr>               <chr>
##  1           32 60           O2,Suc              NND
##  2           14 70           None                DC
##  3            0 0            Stim,CPR,BVM        NND
##  4           38 26           Stim,CPR,O2,BVM     NND
##  5           42 44           Stim,CPR,O2,BVM,Suc NND
##  6           14 50           None                DC
```

```
##  7            0 20          Stim,CPR,O2,BVM,Suc NND
##  8            0 0           Stim,BVM,O2,Suc     NND
##  9           15 48          None                DC
## 10           43 4           Stim,CPR,O2,BVM,Suc NND
## 11           34 0           None                NND
## 12           45 26          Stim,CPR,BVM,Suc    NND
## 13            0 0           None                NND
```

```
## [1] "New variable"
```

```
##      et_hr
##  Min.   :  0
##  1st Qu.:125
##  Median :138
##  Mean   :136
##  3rd Qu.:146
##  Max.   :252
##  NA's   :3
```

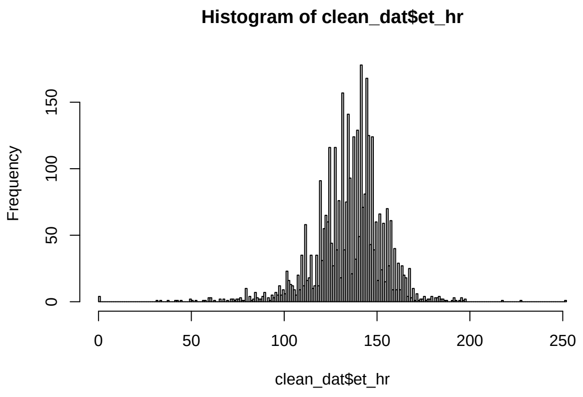


**5.1.1.4 Admission.Temperature** Continuous variable measured in degrees Celsius (to 0.1 precision).

```
##    Min. 1st Qu.  Median    Mean 3rd Qu.    Max.    NA's
##    30.0    36.0    36.5    36.4    37.0    41.0    1027
```

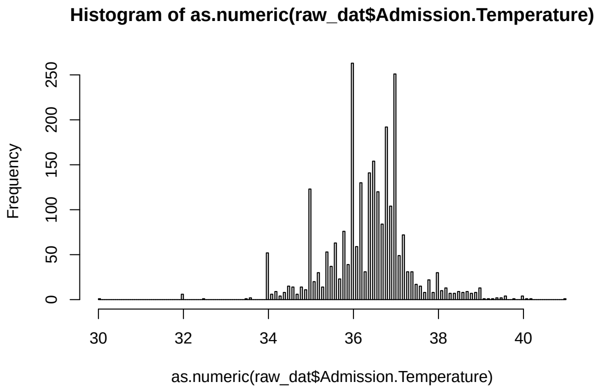

```
##     et_temp
##  Min.   :30.0
##  1st Qu.:36.0
##  Median :36.5
##  Mean   :36.4
##  3rd Qu.:37.0
##  Max.   :41.0
##  NA's   :1027
```

###### 5.1.1.5 Admission.BW & Admission.AW

Continuous variables measured in grams.

- Looking at the distributions of birth weight (BW) and admission weight (AW), some values are clearly invalid.
- It is important not to assume what these values should be (e.g., for "100" the true value may have been "1000", or perhaps "3100").

```
## [1] "Distribution of birth weight"
```

```
##    Min. 1st Qu.  Median    Mean 3rd Qu.    Max.    NA's 
##       2    1950    2700    2592    3200    5200      48
```

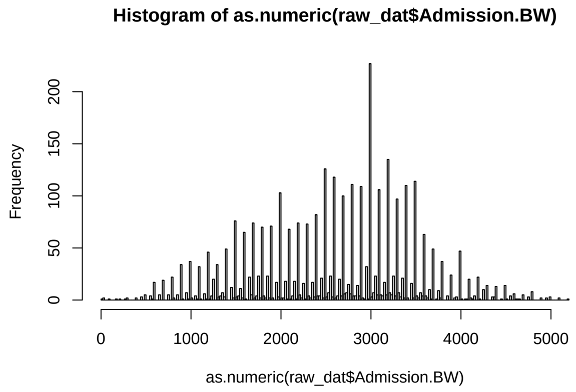


```
## [1] "Lowest values"
```

```
## 
##   2  35  36 100 180 220 270 300 400 450 500 550 580 600 650 690 700 750 800 805 
##   1   1   1   1   1   1   1   2   2   3   5   4   1  17   5   1  18   5  22   2 
## 850 900 920 945 950 955 
##   5  34   1   1   5   1
```

```
## [1] "Distribution of admission weight"
```

```
##    Min. 1st Qu.  Median    Mean 3rd Qu.    Max.    NA's 
##      40    1850    2600    2544    3200    5200    1877
```

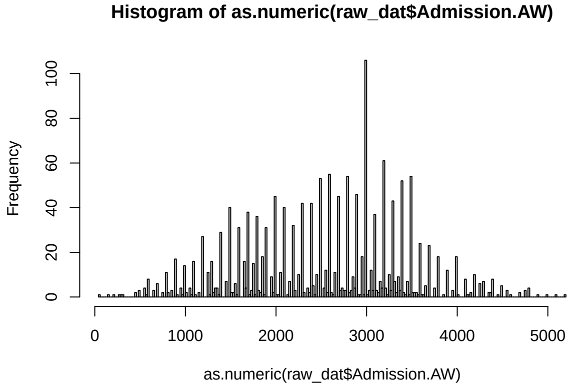


```
## [1] "Lowest values"
```

```
##
##  40 150 220 280 300 310 450 500 550 580 600 650 690 700 750 800 805 850 900 920
##   1   1   1   1   1   1   2   3   4   1   8   3   1   5   2  11   2   3  17   1
## 950
##   3
```

```
## [1] "Cases where BW or AW <500g"
```

```
## # A tibble: 18 x 2
##    Admission.BW Admission.AW
##           <dbl>        <dbl>
##  1         1500          150
##  2          100           NA
##  3          300          300
##  4         3100          310
##  5          300         3000
##  6          450          450
##  7          450           NA
##  8          400           NA
##  9         2800          280
## 10          450          450
## 11          700           40
## 12           35         3375
## 13            2           NA
## 14          400           NA
## 15           36         1340
## 16          270           NA
## 17          180         1800
## 18          220          220
```

Therefore, we assessed how many cases have BW and/or AW missing, and whether it is necessary to have two weight variables (i.e., do BW and AW substantially differ?):

```
## [1] "Birth weight missing"
```

```
## [1] 48
```

```
## [1] "Admission weight missing"
```

```
## [1] 1877
```

```
## [1] "Birth weight missing but admission weight NOT missing"
```

```
## [1] 28
```

```
## [1] "Cases where BW and AW differ (and AW not missing)"
```

```
## # A tibble: 32 x 2
##    Admission.BW Admission.AW
##           <dbl>        <dbl>
##  1         1500          150
##  2         3780         3300
##  3         3100          310
##  4         2120         2100
##  5         1700         1660
##  6         1650         1700
##  7         1540         1890
##  8         2488         2408
##  9          300         3000
## 10         3400         3408
## 11         1500         1350
## 12         2630         2603
## 13         3300         3700
## 14         1700         1550
## 15         1660         1600
## 16         3000         2880
## 17         2700         2600
## 18         1800         1500
## 19         1320         1750
## 20         4800         4560
## 21         1500         1600
## 22         2000         2200
## 23         4000         3900
## 24         2800          280
## 25         1900         1980
## 26         2910         2700
## 27         3100         3400
## 28          700           40
## 29           35         3375
## 30         1470         1275
## 31           36         1340
## 32          180         1800
```

- There are only 32 cases where BW and AW differ.
  - Examining these cases, the differences are relatively small (excluding cases where the value is obviously erroneous). Therefore, it is unnecessary to have a separate variable for AW, and BW will suffice.
- Some values were recorded as very low (i.e. <500g).

- If BW is consistent with the gestational age, the original value is retained.
- If BW is inconsistent with gestational age but AW is consistent, then `et_bw` takes the value of AW.
- Otherwise, if neither BW or AW consistent with gestational age (or AW missing), then original BW value retained and case will be excluded based on inclusion/exclusion criteria for birth weight (see below).
- *N.B. We used the UK-WHO Neonatal and Infant Close Monitoring Growth Chart 2009 to determine weights consistent with each gestational age.*

```
## [1] "New variable"
```

```
##      et_bw
##  Min.   :   2
##  1st Qu.:1950
##  Median :2700
##  Mean   :2595
##  3rd Qu.:3200
##  Max.   :5200
##  NA's   :48
```

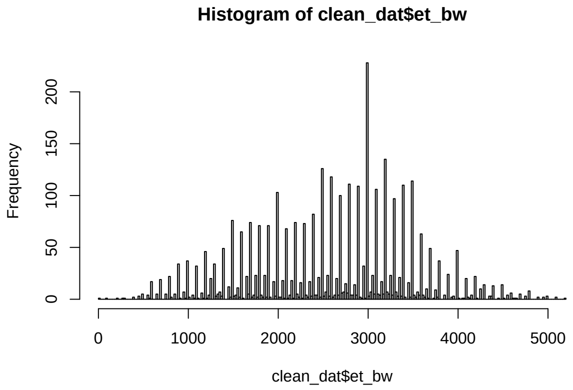


##### 5.1.2 Patient information

The variables to be subset/created from this section are as follows:

| Parent variable | New variable(s) | Comments |
|---|---|---|
| Admission.AdmReason | *informs* pi_bba | "BBA" (yes/no) |
| " " | pi_admreason | *takes original values* (factor) |
| Admission.UID | adm_uid | (string) |
| Admission.session | adm_session | (string) |
| Admission.DateTimeAdmission | adm_datetime | (date-time) |
| Admission.Gender | pi_sex | *takes original values* (factor) |
| Admission.AgeA/B/Cat/C | pi_age | (numeric) |
| Admission.TypeBirth | pi_type | (factor) |
| Admission.Gestation | pi_gest | (numeric) |

###### 5.1.2.1 Admission.AdmReason

Categorical variable with many levels. No changes made to original data.

```
##
##      AD     Apg      BA     BBA    Cong    Conv     DIB      DU      FD     Fev
##      25     392     141     139      46       7     511      10      63     132
##       G    HIVX       J     LBW     Mac     Mec     NTD       O      OM    Prem
##      96      10     155     246     128     240      25     261      18     149
## PremRDS    Risk    Safe     SPn    <NA>
##     443      86     251       3       0
```

```
##   pi_admreason
##  DIB    : 511
##  PremRDS: 443
##  Apg    : 392
##  O      : 261
##  Safe   : 251
##  LBW    : 246
##  (Other):1473
```

###### 5.1.2.2 Admission.UID & Admission.session

String variables. No changes made to original data.

- Admission.UID = the unique identifier for each baby, automatically generated by the Neotree app when a new admission form is created.
- Admission.UID_alphanum = Admission.UID but with non-alphanumeric characters removed. Used for record linkage.
- Admission.session = a unique number assigned to each row of data when imported from the raw JSON files (i.e., `seq_along(1:nrow(data))`). Can be used to merge columns from the other data frames if needed in later analyses.

###### 5.1.2.3 Admission.DateTimeAdmission

String variable representing a date. Converted to POSIXct object.

- The period prior to 1st February 2019 was a 'pilot period'.
  - During this period, healthcare workers were becoming accustomed to the Neotree app and only a subset of admissions and outcomes were recorded.

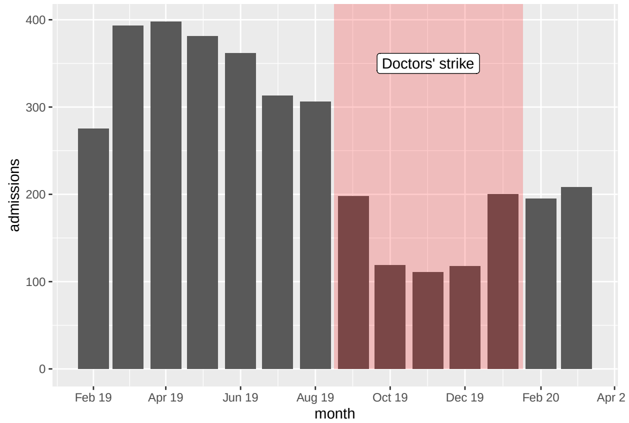

###### 5.1.2.4 Admission.Gender

Categorical variable with three levels.

- Male
- Female
- Unsure

No changes made to original data.

```
##
##    F    M    U <NA>
## 1608 1965    4    0
```

```
## [1] "New variable"
```

```
##  pi_sex
##  f:1608
##  m:1965
##  u:   4
```

###### 5.1.2.5 Admission.AgeA/B/Cat/C

Categorical or string variables representing age at admission:

- Admission.AgeA = Is the baby aged less than 1 week?
  - Binary categorical variable: yes (Y) or no (N)
- Admission.AgeB = If AgeA = yes, the baby's age to the nearest hour
  - String varible in the format `X days, Y hours`
- Admission.AgeCat = If AgeA = yes, the age category that the baby falls into
  - Categorical variable with 5 levels:
    * Fresh newborn (<2 hours-old)
    * Newborn 2-23 hours-old
    * Newborn 24-47 hours-old
    * Infant 48-71 hours-old
    * Infant  72 hours-old
- Admission.AgeC = If AgeA = no, the baby's age to the nearest day
  - String variable in the format `X days`

N.B. If the reason for admission is "dumped baby", then age is not recorded.

```
## # A tibble: 4 x 3
##   variable         n_miss pct_miss
##   <chr>             <int>    <dbl>
## 1 Admission.AgeC     3557   99.4
## 2 Admission.AgeB      657   18.4
## 3 Admission.AgeCat    109    3.05
## 4 Admission.AgeA       13    0.363
```

```
## [1] "Missing both AgeB and AgeCat"
```

```
## [1] 28
```

All age variables have a high proportion of missingness except Admission.AgeCat and Admission.AgeA.

- Since Admission.AgeA is a simple binary question of whether the baby is less than one week-old, using Admission.AgeCat is more informative.
- This means age will be a categorical variable rather than a continuous variable, but this is preferable to reduce the proportion of missing values.

We can transform Admission.AgeB into a continuous variable of age in hours, and then check to ensure Admission.AgeB is congruent with Admission.AgeCat:

```
## [1] "Admission.AgeB, original"
```

```
##  [1] "18 hours"         "1 day, 9 hours"   "1 hour"           "1 day, 9 hours"
##  [5] "16 hours"         "1 day, 5 hours"   "1 day, 5 hours"   "1 day, 15 hours"
##  [9] "6 hours"          "15 hours"         "13 hours"         "2 hours"
## [13] "19 hours"         "4 hours"          "11 hours"         "2 days, 18 hours"
## [17] "14 hours"         "11 hours"         "1 day, 3 hours"   NA
```

```
## [1] "Note some anomalies: negative values or >1 week-old"
```

```
##  [1] "-21 hours"                "-10 hours"
##  [3] "1 month5 days, 16 hours" "1 month1 day, 4 hours"
##  [5] "-17 hours"                "-10 hours"
##  [7] "-3 hours"                 "-18 hours"
##  [9] "-5 hours"                 "-11 hours"
## [11] "-23 hours"                "-20 hours"
## [13] "-20 hours"                "-23 hours"
## [15] "-23 hours"                "-23 hours"
## [17] "-20 hours"                "-5 hours"
## [19] "-10 hours"                "1 month4 days, 7 hours"
## [21] "-6 hours"                 "-22 hours"
## [23] "-21 hours"                "-22 hours"
## [25] "-5 hours"                 "-9 hours"
## [27] "-8 hours"                 "-10 hours"
```

```
## [1] "Check this new variable, age in hours"
```

```
## # A tibble: 10 x 2
##    Admission.AgeB  age_hours
##    <chr>               <dbl>
##  1 18 hours               18
##  2 1 day, 9 hours         33
##  3 1 hour                  1
##  4 1 day, 9 hours         33
##  5 16 hours               16
##  6 1 day, 5 hours         29
##  7 1 day, 5 hours         29
##  8 1 day, 15 hours        39
##  9 6 hours                 6
## 10 15 hours               15
```

```
##    age_hours
##  Min.   :  1.0
##  1st Qu.:  2.0
##  Median :  4.0
##  Mean   : 15.6
##  3rd Qu.: 18.0
##  Max.   :167.0
##  NA's   :685
```

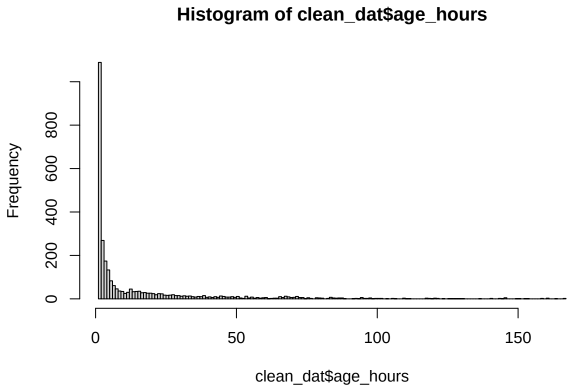


```
## [1] "Generate agecat_new based on age_hours values"
```

```
## [1] "Cases where agecat != agecat_new"
```

```
## [1] 259
```

```
## # A tibble: 6 x 5
##   Admission.AgeA age_hours agecat agecat_new Admission.AgeC
##   <chr>              <dbl> <chr>  <fct>      <chr>
## 1 Y                     33 NB24   NB48       <NA>
## 2 Y                     27 NB24   NB48       <NA>
## 3 Y                     95 INF72  INF        <NA>
## 4 Y                      1 NB24   FNB        <NA>
## 5 Y                     67 INF    INF72      <NA>
## 6 Y                     38 NB24   NB48       <NA>
```

There are some discrepancies between the age from Admission.AgeB (automatically generated by the app from date-time of birth and admission date-time) and the age category selected by the healthcare workers (recorded as Admission.AgeCat).

- These discrepancies occur in relatively few cases and likely represent a misunderstanding of the age category definitions by healthcare workers using the app.
- As Admission.AgeB is generated automatically by the app, it is less liable to errors than Admission.AgeCat.
- Therefore, the following rules will be applied to create the age variable:
  - Where Admission.AgeB is *not* missing, we use this variable to assign the age category.
  - Where Admission.AgeB is missing but Admission.AgeCat is *not* missing, we use the value of Admission.AgeCat.

– Where Admission.AgeCat is missing but Admission.Age == "N", then the baby is older than one week, so is assigned to the "infant" category.
– Where all the above are missing, the new age variable is missing.

```
## [1] 14
```

```
## # A tibble: 14 x 4
##    Admission.AgeA Admission.AgeB  agecat pi_age
##    <chr>          <chr>           <chr>  <chr>
##  1 N              15 hours        <NA>   NB24
##  2 N              2 days, 6 hours <NA>   INF72
##  3 N              2 days, 3 hours <NA>   INF72
##  4 N              14 hours        <NA>   NB24
##  5 N              1 day, 17 hours <NA>   NB48
##  6 N              21 hours        <NA>   NB24
##  7 N              1 hour          <NA>   FNB
##  8 N              1 day, 3 hours  <NA>   NB48
##  9 N              1 day, 20 hours <NA>   NB48
## 10 N              16 hours        <NA>   NB24
## 11 N              1 day, 3 hours  <NA>   NB48
## 12 N              1 day           <NA>   NB48
## 13 N              1 day, 2 hours  <NA>   NB48
## 14 N              1 day, 11 hours <NA>   NB48
```

```
##    pi_age
##  fnb  :1302
##  dol1 :1650
##  dol2 : 300
##  dol3 : 148
##  older: 146
##  NA's :  31
```

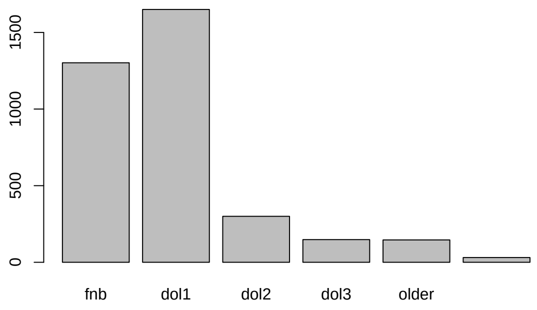


There are several cases where Admission.AgeA would suggest the baby is ≥ 1 week-old, yet Admission.AgeB (and, thus, pi_age) does not correlate with this. Admission.AgeB is likely the most accurate source of age and so this value will be used.

###### 5.1.2.6 Admission.TypeBirth

Categorical variable with six levels:

- Singleton

- Twin number 1
- Twin number 2
- Triplet number 1
- Triplet number 2
- Triplet number 3

No changes made to original data.

```
##
##    S  Tr1  Tr2  Tr3  Tw1  Tw2 <NA>
## 3217    6    5    6  187  153    3
```

```
##       pi_type
##  singleton:3217
##  twin1    : 187
##  twin2    : 153
##  triplet1 :   6
##  triplet2 :   5
##  triplet3 :   6
##  NA's     :   3
```

**5.1.2.7 Admission.Gestation** Continuous variable measured in weeks. No changes made to original data.

```
##    Min. 1st Qu.  Median    Mean 3rd Qu.    Max.    NA's
##    20.0    35.0    38.0    36.5    39.0    43.0       1
```

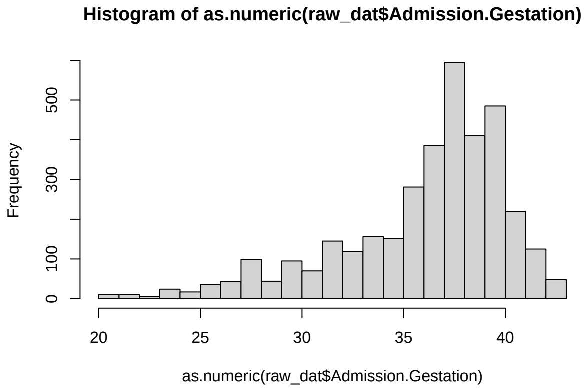


```
##     pi_gest
##  Min.   :20.0
##  1st Qu.:35.0
##  Median :38.0
##  Mean   :36.5
##  3rd Qu.:39.0
##  Max.   :43.0
##  NA's   :1
```

###### 5.1.3 Examination

The variables to be subset/created from this section are as follows:

| Parent variable | New variable(s) | Comments |
|---|---|---|
| Admission.Fontanelle | oe_fontanelle | *takes values* (factor) |
| Admission.Activity | oe_activity | *takes values* (factor) |
| Admission.SignsRD | oe_nasalflare | "NFL" (yes/no) |
| " " | oe_retractions | "CHI" (yes/no) |
| " " | oe_grunt | "GR" (yes/no) |
| Admission.WOB | oe_wob | *takes values* (factor), add "normal" if SignsRD == "None", NA if SignsRD missing |
| Admission.Colour | oe_colour | *takes values* (factor) |
| Admission.Abdomen | oe_abdodist | "Dist" (yes/no) |
| Admission.Umbilicus | oe_omphalitis | "Inf" (yes/no) |
| Admission.Skin | oe_abskin | not "None" (yes/no) |

###### 5.1.3.1 Admission.Fontanelle

Categorical variable with three levels:

- Bulging = "Bulging"
- Flat = "Flat"
- Sunken = "Sunken"

No changes made to original data.

```
##
## Bulg Flat Sunk <NA>
##   16 3546   15    0
```

```
##  oe_fontanelle
##  flat   :3546
##  sunken :  15
##  bulging:  16
```

###### 5.1.3.2 Admission.Activity

Categorical variable with five levels:

- Alert = "Alert, active, appropriate"
- Coma = "Coma (unresponsive)"
- Convulsions = "Seizures, convulsions, or twitchings"
- Irritable = "Irritable"
- Lethargic = "Lethargic, quiet, decreased activity"

No changes made to original data.

```
##
## Alert  Coma  Conv Irrit  Leth  <NA>
##  2791    48    16    77   645     0
```

```
##     oe_activity
##  alert    :2791
##  lethargic: 645
##  irritable:  77
##  seizures :  16
##  coma     :  48
```

###### 5.1.3.3 Admission.SignsRD

Categorical variable with five levels:

- Chest retractions = "Chest in-drawings"
- Grunting = "Grunting"
- Nasal flaring = "Nasal flaring"
- ~~Gasping = "Gasping"~~
- ~~Stridor = "Stridor"~~
- ~~Head nodding = "Head nodding"~~
- ~~Tracheal tug = "Tracheal tug"~~
- None

Of these, only the first three categories are candidate predictors for this study. No changes made to original data.

```
##
##                  CHI              CHI,GR          CHI,HN,NFL
##                  268                  71                   1
##              CHI,NFL          CHI,NFL,GR                Gasp
##                  488                 307                  45
##             Gasp,CHI         Gasp,CHI,GR        Gasp,CHI,NFL
##                   16                   7                  19
##      Gasp,CHI,NFL,GR             Gasp,GR     Gasp,HN,CHI,NFL
##                   20                   4                   1
##   Gasp,HN,CHI,NFL,GR            Gasp,NFL        Gasp,NFL,CHI
##                    3                   3                   6
##      Gasp,NFL,CHI,GR                  GR              HN,CHI
##                    4                  35                   4
##            HN,CHI,GR          HN,CHI,NFL       HN,CHI,NFL,GR
##                    5                  21                  48
##               HN,NFL          HN,NFL,CHI       HN,NFL,CHI,GR
##                    1                   1                   7
##            HN,NFL,GR                 NFL             NFL,CHI
##                    1                 189                  87
##           NFL,CHI,GR              NFL,GR              NFL,HN
##                   31                  44                   1
##           NFL,HN,CHI           NFL,HN,GR                None
##                    1                   1                1788
##                   ST              ST,CHI       ST,CHI,NFL,GR
##                    1                   1                   1
##                ST,HN              ST,NFL                  TT
##                    1                   1                   2
##               TT,CHI          TT,CHI,NFL       TT,CHI,NFL,GR
##                    4                   8                  10
##      TT,Gasp,CHI,NFL  TT,Gasp,CHI,NFL,GR TT,Gasp,HN,CHI,NFL,GR
##                    1                   2                   1
##   TT,Gasp,NFL,CHI,GR           TT,HN,CHI       TT,HN,CHI,NFL,GR
```

```
##                  1                  1                  6
##      TT,HN,NFL,CHI,GR           TT,NFL,CHI         TT,NFL,CHI,GR
##                  1                  3                  1
##         TT,NFL,HN,CHI      TT,ST,CHI,NFL,GR               <NA>
##                  1                  1                  1
```

```
##  oe_nasalflare oe_retractions oe_grunt
##  no  :2253     no  :2117      no  :2964
##  yes :1323     yes :1459      yes : 612
##  NA's:   1     NA's:   1      NA's:   1
```

**5.1.3.4 Admission.WOB** Categorical variable with three levels:

- Mildly increased work of breathing (WOB) = "Mild"
- Moderately increased WOB = "Moderate"
- Severely increased WOB = "Severe"

N.B. At the time of study, this variable was only completed if Admission.SignsRD was recorded as "nasal flaring", "chest retractions", "head nodding", "grunting", or "tracheal tug". A value was *not* entered if Admission.SignsRD was recorded as "gasping" or "stridor".

The following rules were applied to create the new WOB variable:

- NA if Admission.SignsRD is NA;
- "normal" if Admission.SignsRD == "none";
- NA if Admission.SignsRD == "gasping" or "stridor".

```
##
## Mild  Mod  Sev <NA>
##  520  885  339 1833
```

```
##       oe_wob
##  normal  :1788
##  mild    : 519
##  moderate: 885
##  severe  : 338
##  NA's    :  47
```

```
## [1] normal normal severe mild   mild   severe
## Levels: normal mild moderate severe
```

**5.1.3.5 Admission.Colour** Categorical variable with four levels:

- Pink = "Pink"
- Blue = "Blue"
- White = "White"
- Yellow = "Yellow"

No changes made to original data.

```
##
##  Blue  Pink White  Yell  <NA>
##   129  3353    21    74     0
```

```
##   oe_colour
##  pink  :3353
##  pale  :  21
##  blue  : 129
##  yellow:  74
```

###### 5.1.3.6 Admission.Abdomen

Categorical variable with eight levels:

- Distended = "Distended"
- ~~Hepatomegaly = "Hepatomegaly"~~
- ~~Splenomegaly = "Splenomegaly"~~
- ~~Abdominal mass = "Abdominal mass"~~
- ~~Gastroschisis = "Gastroschisis"~~
- ~~Omphalocele = "Omphalocele"~~
- ~~Prune belly = "Prune belly"~~
- Normal = "Soft and normal"

Of these, only abdominal distention is a candidate predictor for this study. No changes made to original data.

```
##
##            AbMass       AbMass,Dist       AbMass,PrunB              Dist
##                 4                 4                  1                45
##        Dist,PrunB            GSchis             HepMeg       HepMeg,Dist
##                 1                75                  2                 1
##              Norm              Omph          Omph,Norm        PrunB,Norm
##              3419                15                  1                 5
##       SplMeg,Dist SplMeg,Dist,HepMeg               <NA>
##                 1                 1                  2
```

```
##  oe_abdodist
##  no  :3522
##  yes :  53
##  NA's:   2
```

###### 5.1.3.7 Admission.Umbilicus

Categorical variable with four levels:

- Infected = "Red skin all around umbilicus"
- ~~Blood-stained = "Bleeding"~~
- ~~Meconium-stained = "Meconium stained"~~
- ~~Abnormal = "Abnormal looking"~~
- ~~Hernia = "Umbilical hernia"~~
- Normal = "Healthy and clean"

Of these, only omphalitis (i.e. "infected" umbilicus) is a candidate predictor for this study. No changes made to original data.

```
##
##      Abn   Abn,H      Bl    Bl,H       H     Inf Inf,Abn     Mec    Norm    <NA>
##       52       1       6       1       4      16       1      64    3432       0
```

```
## oe_omphalitis
## no :3560
## yes:  17
```

###### 5.1.3.8 Admission.Skin

Categorical variable with four levels:

- Pustules = "Pustules all over"
- Abscess = "Big boil/abscess"
- Rash = "Other skin rash"
- None = "Normal"

Due to distribution of categories, dichotomised into "abnormal skin" yes/no.

```
##
##      None      Rash Rash,PUST      <NA>
##      3540        36         1         0
```

```
## oe_abskin
## no :3540
## yes:  37
```

##### 5.1.4 Symptom review

The variables to be subset/created from this section are as follows:

| Parent variable | New variable(s) | Comments |
|---|---|---|
| Admission.Vomiting | hx_vomit | *modified values* (factor) |

###### 5.1.4.1 Admission.Vomiting

Categorical variable with five levels:

- Yes, vomiting = "Vomiting all feeds"
- Yes, green vomit = "Vomiting bright green"
- Yes, bloody vomit = "Vomiting with blood"
- Posseting = "Small milky possets after feeds (normal)"
- No vomiting = "NONE"

In the original variable, some cases were coded with multiple categories. The new variable was recoded to ensure mutually exclusive groups.

```
##
##        No      Poss       Yes Yes,YesGr     YesBl     YesGr      <NA>
##      3482        21        18         2         6        48         0
```

```
##    hx_vomit
## no      :3503
## yellow :  18
## bilious:  50
## bloody :   6
```

##### 5.1.5 Maternal history (obstetric history)

The variables to be subset/created from this section are as follows:

| Parent variable | New variable(s) | Comments |
|---|---|---|
| Admission.ROMlength | oh_prom2 | "PROM" (yes/no) |
| Admission.RFSepsis | oh_prom | "PROM" (yes/no) |
| " " | oh_matfever | "MF" (yes/no) |
| " " | oh_offliquor | "OL" (yes/no) |
| Both of the above | co_prom | "yes" if oh_prom OR oh_prom2 == "yes" (yes/no) |
| Admission.ModeDelivery | oh_delivery | *takes values* (factor) |

**5.1.5.1 Admission.ROMlength** Binary categorical variable:

- PROM = ">18 hours"
- NOPROM = "<18 hours"

No changes made to original data.

N.B. This is one of two PROM-related data points collected:

1. Admission.ROMlength (this variable)
2. Admission.RFSepsis (categorical variable with one catergory for PROM) - see below

```
##
## NOPROM   PROM   <NA>
##   1894    361   1322
```

```
##
##   no  yes <NA>
## 1894  361 1322
```

**5.1.5.2 Admission.RFSepsis** Categorical variable with seven levels:

- Prolonged rupture of membranes = "PROM more than 18 hrs"
- Maternal fever during labour = "Maternal fever in labour"
- Offensive liquor = "Offensive liquor"
- ~~Prematurity = "Prematurity <37 weeks"~~
- ~~Prolonged second stage of labour = "Prolonged second stage"~~
- ~~Born before arrival to hospital = "Born before arrival (BBA)"~~
- None

Of these, only the first three are candidate predictors for this study. Although prematurity is also a candidate predictor, this information is obtained more precisely from Admission.Gestation (*see above*).

No changes made to original data.

```
##
##                BBA            BBA,OL         BBA,Prem               MF     MF,BBA,Prem
##                127                 1               27                8               2
##       MF,Pr2nd,OL    MF,Pr2nd,PROM          MF,Prem      MF,Prem,BBA         MF,PROM
##                  1                 2                4                1               3
##        MF,PROM,OL MF,PROM,Prem,OL             NONE               OL         OL,Prem
##                  1                 1             2029               89               2
##              Pr2nd          Pr2nd,OL       Pr2nd,Prem       Pr2nd,PROM   Pr2nd,PROM,OL
##                 63                16                5               16               5
## Pr2nd,PROM,Prem              Prem         Prem,BBA          Prem,OL            PROM
##                  1               773               69               11             167
##           PROM,BBA     PROM,BBA,Prem          PROM,OL     PROM,OL,Prem       PROM,Prem
##                  1                 1               39                2             100
##      PROM,Prem,BBA      PROM,Prem,OL             <NA>
##                  2                 7                1
```

```
##  oh_prom     oh_matfever oh_offliquor
##  no  :3228   no  :3553   no  :3401
##  yes : 348   yes :  23   yes : 175
##  NA's:   1   NA's:   1   NA's:   1
```

**5.1.5.3 Creating a single variable to capture PROM** As mentioned above, there are two PROM-related data points collected:

1. Admission.ROMlength - now `oh_prom2` from above
2. `Admission.RFSepsis == "PROM"` - now `oh_prom` from above

Recoded into a single variable with "yes" if either of the above variables suggest the presence of PROM.

```
## [1] "Compare coding & distribution between both PROM variables..."
```

```
##  oh_prom     oh_prom2
##  no  :3228   no  :1894
##  yes : 348   yes : 361
##  NA's:   1   NA's:1322
```

```
##
##          no  yes
##   no   1881   36
##   yes    12  325
```

```
## [1] "New combined variable..."
```

```
##   no  yes
## 3193  384
```

**5.1.5.4 Admission.ModeDelivery** Categorical variable with five levels:

- Emergency caesarean section = "Emergency caesarean section"
- Elective caesarean section = "Elective caesarean section"

- Forceps = “Forceps extraction”
- Spontaneous vaginal delivery = “Spontaneous vaginal delivery”
- Ventouse = “Vacuum extraction”

No changes made to original data.

```
##
##  ECS ElCS  For  SVD Vent <NA>
##  726  186    1 2620   44    0
```

```
##       oh_delivery
##  svd        :2620
##  electiveCS : 186
##  emergencyCS: 726
##  forceps    :   1
##  ventouse   :  44
```

#### 5.2 Data collected by outcome forms

There are two groups of outcome variables to consider:

1. Participant demographics
2. Model outcome data

##### 5.2.1 Participant demographics

The variables to be subset/created from this section are as follows:

| Parent variable | New variable(s) | Comments |
|---|---|---|
| Discharge.session | dis_session | (string) |
| Discharge.NeoTreeID | dis_uid | (string) |
| Discharge.NeoTreeOutcome | outcome | *takes values* (factor) |
| Discharge.DateTimeDischarge | outcome_datetime | (date-time) |
| Discharge.DateTimeDeath | outcome_datetime | (date-time) |
| *several* | adm_dur | (period) |

###### 5.2.1.1 Discharge.NeoTreeID & Discharge.session

String variables.

- Discharge.NeoTreeID = the unique identifier for each baby, automatically generated by the Neotree app when a new admission form is created. Entered manually by the healthcare worker completing the outcome form.
- Discharge.NeoTreeID_alphanum = the unique identifier but with non-alphanumeric characters removed. Used for record linkage.
- Discharge.session = a unique number assigned to each row of data when imported from the raw JSON files (i.e., `seq_along(1:nrow(data))`). Can be used to merge columns from other data frames if required in future analyses.

No changes made to original data.

```
##    dis_uid          dis_session
##  Length:3577        Length:3577
##  Class :character   Class :character
##  Mode  :character   Mode  :character
```

###### 5.2.1.2 Discharge.NeoTreeOutcome

Categorical variable with five levels:

- Discharged = "Discharged"
- Death = "Died"
- Transferred within the hospital = "Transferred to other ward"
- Transferred to another hospital or facility = "Transferred to other hospital"
- Absconded = "Absconded"

Dichotomised into died/discharged. For this study, we considered a participant to be discharged if any outcome other than "death" was recorded.

```
##
##  ABS   DC  NND  TRH  TRO <NA>
##    3 2887  679    6    2    0
```

```
##        outcome
##  died      : 679
##  discharged:2898
```

###### 5.2.1.3 Discharge.DateTimeDischarge & Discharge.DateTimeDeath

String variables representing dates.

```
## [1] "Ensure outcome matches date variable recorded..."
```

```
## [1] "Discharge.DateTimeDischarge missing..."
```

```
## [1] 2900
```

```
## [1] "Discharge.DateTimeDeath missing..."
```

```
## [1] 681
```

```
## [1] "Both missing..."
```

```
## [1] 4
```

There are 4 cases where both a discharge date and date of death are recorded. For these, we used the date corresponding to the recorded outcome.

###### 5.2.1.4 Admission duration

It is useful to have a variable denoting the admission duration for each participant. Calculated from the admission and outcome dates.

```
##    Min. 1st Qu.  Median    Mean 3rd Qu.    Max.
##  -0.708   1.288   2.547   5.204   5.612  85.053
```

```
## [1] 48
```

There are 48 cases where admission duration is $\leq 0$.

- These most likely represent errors when inputting the admission and/or outcome date.
- Although a tolerance of outcome date $\leq 1$ day prior to admission date was allowed for record linkage, cases with negative admission durations were excluded from the main analysis because this anomaly questioned the accuracy of some other variables for that participant, e.g., chronological age (which is calculated automatically within the app from birth date-time and admission date-time).

#### 5.3 Model outcome data

The primary outcome was early-onset sepsis, defined as sepsis with onset within the first 72 hours of life, as diagnosed by the treating consultant neonatologist.

##### 5.3.1 Supporting variables

The variables required to create the outcome variable are as follows:

| Variable | Comments |
|---|---|
| Discharge.DIAGDIS1 | Primary discharge diagnosis |
| Discharge.DIAGDIS1OT | Free text field if primary discharge diagnosis == "other" |
| Discharge.OthProbs | Other problems during admission |
| Discharge.OthProbsOth | Free text field if other problems == "other" |
| Discharge.CauseDeath | Primary cause of death |
| Discharge.CauseDeathOther | Free text field if primary cause of death == "other" |
| Discharge.ContCauseDeath | Contributory cause(s) of death |
| Discharge.ContCauseDeathOth | Free text field of contributory cause of death == "other" |

```
##
##    AN   BBA    BI    BO   CHD  DEHY  EONS    FD     G   HIE  HIVX HIVXH HIVXL
##     4    95    11     4     8     8   197    38     8   376    11    15    48
##  JAUN   LBW  LONS    MA   Mac    MD    NB   OCA    OM   OTH    PN    PR PRRDS
##   231   126    26   119   134     4    40    29    12   314     9   166   269
##    Ri  Safe   TTN  Twin  <NA>
##    72   220   294    11   678
```

```
##
##    ASP     CA   EONS Gastro    HIE   LONS    MAS    NEC    OTH     PN     PR
##     22     17     35     75    117     10      5      2     63      6     39
##  PRRDS   <NA>
##    288   2898
```

```
## [1] "Ensure all discharges have discharge diagnosis recorded..."
```

```
## [1] 0
```

```
## [1] 0
```

```
## [1] "Ensure all deaths have cause of death recorded..."
```

```
## [1] 0
```

```
## [1] 0
```

```
## [1] "New variables..."
```

```
##    diagnosis    diagnosis_other     diagnosis2   diagnosis2_other  
##  HIE    : 376   Length:3577        NONE   :1449   Length:3577       
##  OTH    : 314   Class :character   OTH    : 231   Class :character  
##  TTN    : 294   Mode  :character   LBW    : 181   Mode  :character  
##  PRRDS  : 269                      JAU    : 147                     
##  JAUN   : 231                      HIVX   :  95                     
##  (Other):1415                      (Other): 795                     
##  NA's   : 678                      NA's   : 679                     
##   cause_death    cause_death_other   cause_death2  cause_death2_other
##  PRRDS  : 288   Length:3577        NONE   : 221   Length:3577       
##  HIE    : 117   Class :character   LBW    :  78   Class :character  
##  Gastro :  75   Mode  :character   OTH    :  45   Mode  :character  
##  OTH    :  63                      PRRDS  :  33                     
##  PR     :  39                      EONS   :  25                     
##  (Other):  97                      (Other): 277                     
##  NA's   :2898                      NA's   :2898                     
```

##### 5.3.2 Outcome variable (early-onset neonatal sepsis)

Binary categorical variable of early-onset sepsis yes/no.

First, we explored the free text fields for variations of "early-onset sepsis" that would need to be captured by the outcome variable:

```
# Explore free text (too long to print in full):

# clean_dat %>%
#   select(diagnosis_other) %>%
#   filter(grepl("sep|eons|early", diagnosis_other, ignore.case = T))
#
# clean_dat %>%
#   select(diagnosis2_other) %>%
#   filter(grepl("sep|eons|early", diagnosis2_other, ignore.case = T))
#
# clean_dat %>%
#   select(cause_death_other) %>%
#   filter(grepl("sep|eons|early", cause_death_other, ignore.case = T))
#
# clean_dat %>%
#   select(cause_death2_other) %>%
#   filter(grepl("sep|eons|early", cause_death2_other, ignore.case = T))
```

Relevant free text entries identified:

| Variable | Relevant free text entries |
|---|---|
| Discharge.DIAGDIS1OT | *None* |
| Discharge.OthProbsOth | "Early Onset Neonatal Sepsis" |
| Discharge.CauseDeathOther | "Early onset neonatal sepsis", "earlyonset neonatal sepsis" |
| Discharge.ContCauseDeathOth | *None* |

N.B. "Risk of sepsis", "unconfirmed sepsis" or "sepsis" were not included.

Next, we created the outcome variable.

```r
# Create variable
clean_dat <- clean_dat %>%
  mutate(sepsis = factor(
    case_when(
      # 1. Discharge diagnosis of EONS:
      diagnosis == "EONS" ~ "yes",
      # 2. Other discharge problem includes EONS:
      grepl("EONS", diagnosis2) ~ "yes",
      grepl("Early Onset Neonatal Sepsis", diagnosis2_other) ~ "yes",
      # 3. Cause of death of EONS:
      cause_death == "EONS" ~ "yes",
      grepl(
        "Early onset neonatal sepsis|earlyonset neonatal sepsis",
        cause_death_other
      ) ~ "yes",
      # 4. Contributory cause of death includes EONS:
      grepl("EONS", cause_death2) ~ "yes",
      # Else, no diagnosis of EONS:
      TRUE ~ "no"
    )
  ))

# Check new variable
clean_dat %>%
  select(sepsis) %>%
  summary()
```

```
##  sepsis
##  no :3170
##  yes: 407
```

##### 5.3.3 Inclusion and exclusion criteria

Our inclusion and exclusion criteria were:

| Inclusion criteria | Exclusion criteria |
|---|---|
| Chronological age <72 hours | Not singletons or first-born multiples |
| Gestation 32+0 weeks at birth | Died at admission to the unit (HR or RR = 0) |
| Birth weight 1500 grams | Major congenital anomalies* |
| - | Anomalous admission duration (<0 days) |

*Major congenital anomalies included congenital heart defects, open spina bifida, gastroschisis or omphalocele, and/or genetic syndromes.

The counts of participants excluded due to each criterion are:

```
## # A tibble: 7 x 2
##   criterion                           count
##   <chr>                               <int>
## 1 Admitted  72h of life                 146
## 2 Very premature                        454
```

```
## 3 Very low birth weight                    408
## 4 Dead on admission                         11
## 5 Not singleton or first-born multiple   164
## 6 Major congenital anomaly                 182
## 7 Anomalous admission duration              47
```

##### 5.3.4 Flow diagram of participant inclusion

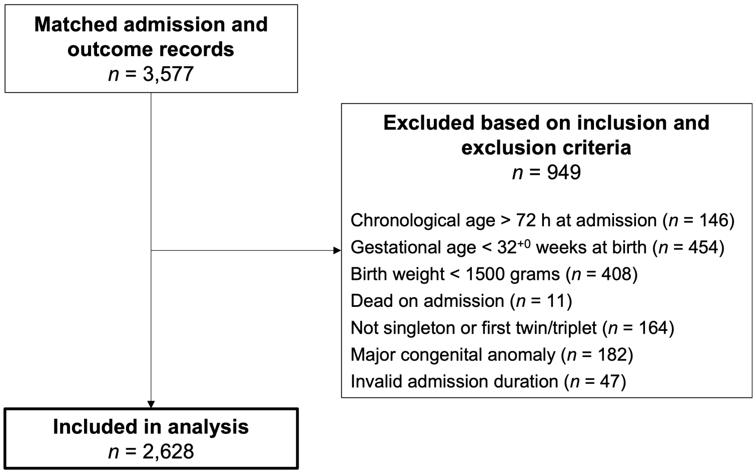

### 6 Missing data

Description of missing data analysis.

#### 6.1 Assess missingness

##### 6.1.1 Visualise data frame

A graphical representation of the data types and proportion of missing values for each variable is shown below. Ancillary variables that are not required for modelling are not shown. Variables in the data frame are plotted on the x-axis and each observation (i.e. participant) is plotted on the y-axis. Missing values are shaded grey.

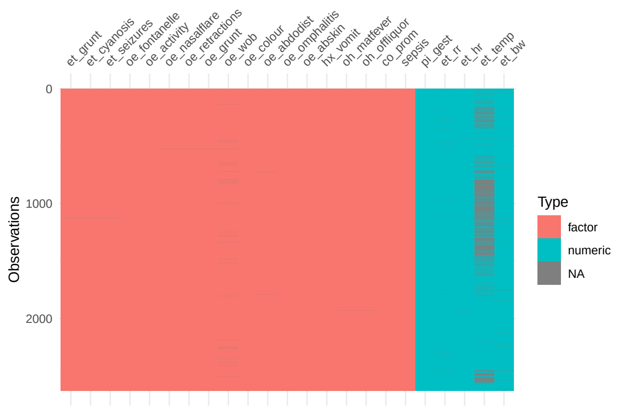


##### 6.1.2 Variable-wise missingness

The number and percentage of missing values for each variable is shown below. In total, 14 variables had missing values.

```
## # A tibble: 23 x 3
##    variable       n_miss pct_miss
##    <chr>           <int>    <dbl>
##  1 et_temp           814   31.0
##  2 et_bw              32    1.22
##  3 oe_wob             26    0.989
##  4 et_rr              22    0.837
##  5 et_hr               3    0.114
##  6 oe_abdodist         2    0.0761
##  7 et_grunt            1    0.0381
##  8 et_cyanosis         1    0.0381
##  9 et_seizures         1    0.0381
## 10 oe_nasalflare       1    0.0381
## 11 oe_retractions      1    0.0381
## 12 oe_grunt            1    0.0381
## 13 oh_matfever         1    0.0381
## 14 oh_offliquor        1    0.0381
## 15 pi_gest             0    0
## 16 oe_fontanelle       0    0
## 17 oe_activity         0    0
```

```
## 18 oe_colour          0   0
## 19 oe_omphalitis      0   0
## 20 oe_abskin          0   0
## 21 hx_vomit           0   0
## 22 co_prom            0   0
## 23 sepsis             0   0
```

##### 6.1.3 Case-wise missingness

Most participants had no missing data and, among those who did, the majority were only missing values for one predictor (most commonly temperature at admission).

```
## # A tibble: 6 x 3
##   n_miss_in_case n_cases pct_cases
##            <int>   <int>     <dbl>
## 1              0    1757   66.9
## 2              1     841   32.0
## 3              2      27    1.03
## 4              3       1    0.0381
## 5              4       1    0.0381
## 6              5       1    0.0381
```

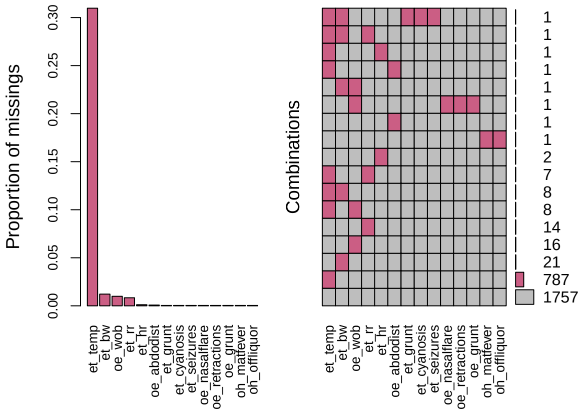


```
##
##  Variables sorted by number of missings:
##        Variable       Count
##         et_temp 0.309741248
##           et_bw 0.012176560
##          oe_wob 0.009893455
##           et_rr 0.008371385
##           et_hr 0.001141553
##     oe_abdodist 0.000761035
##        et_grunt 0.000380518
##     et_cyanosis 0.000380518
##     et_seizures 0.000380518
##   oe_nasalflare 0.000380518
##  oe_retractions 0.000380518
##        oe_grunt 0.000380518
##     oh_matfever 0.000380518
##    oh_offliquor 0.000380518
```

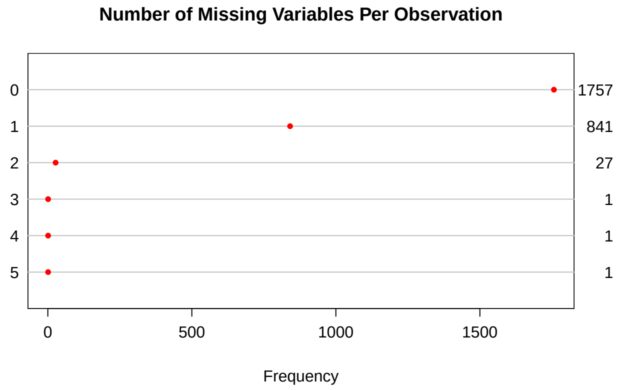


The dendrogram below shows predictors that were commonly missing together.

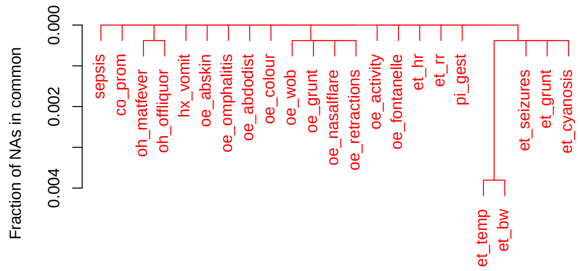


##### 6.1.4 Relationship between missing temperature and the study outcome

There was no evidence of an association between having a missing value for temperature at admission and the primary outcome of early-onset sepsis:

```
##
##        no  yes
##   0 1596  218
##   1  735   79
```

```
##
## Call:
## glm(formula = sepsis ~ na_temp, family = "binomial", data = dat)
##
## Deviance Residuals:
##     Min       1Q   Median       3Q      Max
## -0.506  -0.506  -0.506  -0.452   2.160
##
## Coefficients:
##             Estimate Std. Error z value Pr(>|z|)
## (Intercept)  -1.9908     0.0722  -27.57   <2e-16 ***
## na_temp      -0.2397     0.1387   -1.73    0.084 .
```

```
## ---
## Signif. codes:  0 '***' 0.001 '**' 0.01 '*' 0.05 '.' 0.1 ' ' 1
##
## (Dispersion parameter for binomial family taken to be 1)
##
##     Null deviance: 1854.2  on 2627  degrees of freedom
## Residual deviance: 1851.1  on 2626  degrees of freedom
## AIC: 1855
##
## Number of Fisher Scoring iterations: 4
```

| **Characteristic** | **OR** | **95% CI** | **p-value** |
|---|---|---|---|
| na_temp | 0.79 | 0.60, 1.03 | 0.084 |

##### 6.1.5 Relationship between missing temperature and time

Towards the start of the Neotree project, there was a limited number of thermometers available to measure temperature and, therefore, time since the start of the study is a plausible predictor of missingness.

Indeed, most missing values for temperature at admission occurred near the start of data collection. This suggests that temperature was missing at random (MAR) conditional on time since start of the project.

The matrix plot below shows missing values in red, with each participant sorted by their admission date (i.e. time since the start of data collection).

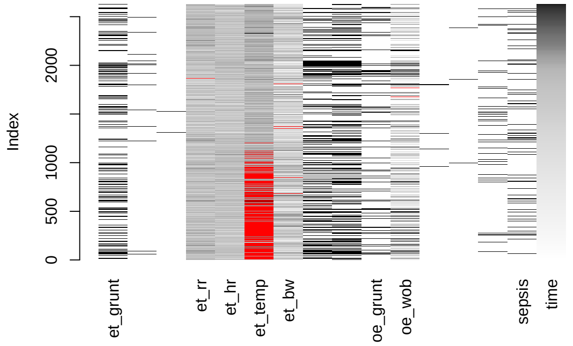


Furthermore, the below figure and a logistic regression analysis demonstrate that time since the start of data collection was a significant predictor of temperature at admission being missing.

Notably, the average recorded temperature was approximately 0.5°C higher during the first 100 days compared to the rest of the data collection period. It is plausible that, during the first 100 days, healthcare workers were more likely to record temperature for ‘sicker’ babies who were thus more likely to have an elevated temperature. Nevertheless, a wide range of participant characteristics were collected by the Neotree app and were included in the imputation model.

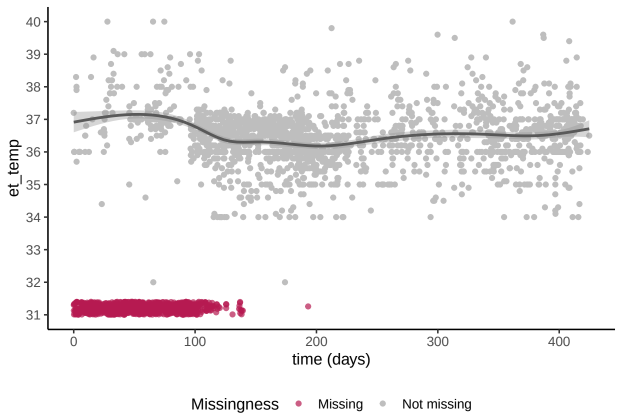


```
##
## Call:
## glm(formula = na_temp ~ time, family = "binomial", data = dat)
##
## Deviance Residuals:
##     Min       1Q   Median       3Q      Max
## -2.6405  -0.3237  -0.0218   0.3853   2.9314
##
## Coefficients:
##             Estimate Std. Error z value Pr(>|z|)
## (Intercept)  3.45556    0.16478    21.0   <2e-16 ***
## time        -0.04006    0.00172   -23.2   <2e-16 ***
## ---
## Signif. codes:  0 '***' 0.001 '**' 0.01 '*' 0.05 '.' 0.1 ' ' 1
##
## (Dispersion parameter for binomial family taken to be 1)
##
##     Null deviance: 3252.9  on 2627  degrees of freedom
## Residual deviance: 1462.6  on 2626  degrees of freedom
## AIC: 1467
##
## Number of Fisher Scoring iterations: 7
```

| **Characteristic** | **OR** | **95% CI** | **p-value** |
|---|---|---|---|
| time | 0.96 | 0.96, 0.96 | <0.001 |

#### 6.2 Impute missing values

The imputation model contained all candidate predictors, the outcome of sepsis, and ancillary variables included in the descriptive analysis or that were determined to predict missingness (i.e. time, see above).

Data were assumed to be MAR and 40 imputed datasets were created with 20 iterations. There is no consensus on the optimal number of imputations for multiple imputation, but 40 was chosen based on 33.1% of participants having at least one missing value.

The performance of the imputation model is shown below:

```
## [1] "Imputation method for each variable..."
```

```
##         pi_gest          et_bw    oh_matfever   oh_offliquor        co_prom
##              ""          "pmm"       "logreg"       "logreg"             ""
##        et_grunt          et_rr          et_hr        et_temp    oe_activity
##        "logreg"          "pmm"          "pmm"          "pmm"             ""
##   oe_nasalflare oe_retractions       oe_grunt         oe_wob    et_cyanosis
##        "logreg"       "logreg"       "logreg"      "polyreg"       "logreg"
##     et_seizures  oe_fontanelle      oe_colour     oe_abdodist  oe_omphalitis
##        "logreg"             ""             ""       "logreg"             ""
##       oe_abskin       hx_vomit         sepsis           time         pi_sex
##              ""             ""             ""             ""             ""
##          pi_age        outcome
##       "polyreg"             ""
```

```
## [1] "Diagnostic plots..."
```

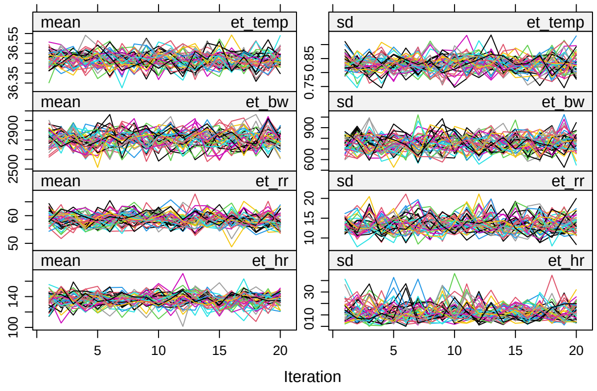


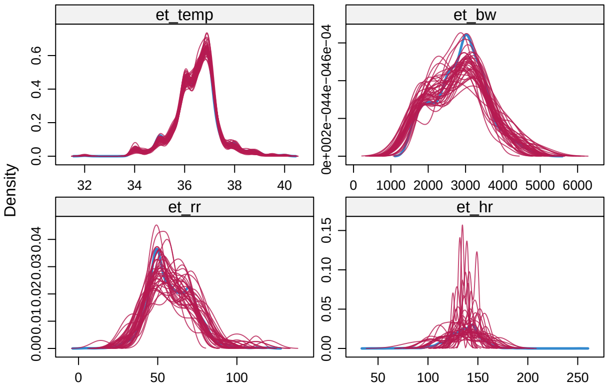


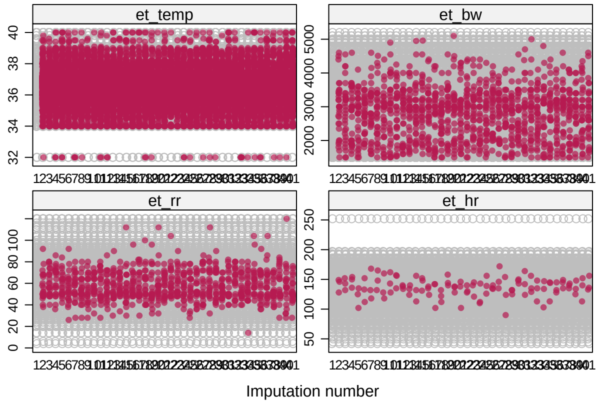

### 7 Descriptive statistics

Descriptive analysis of included participants. Data are presented for the observed data only (i.e. before MICE) using pairwise deletion of missing values.

#### 7.1 Distribution of continuous variables

```
## $pi_gest
```

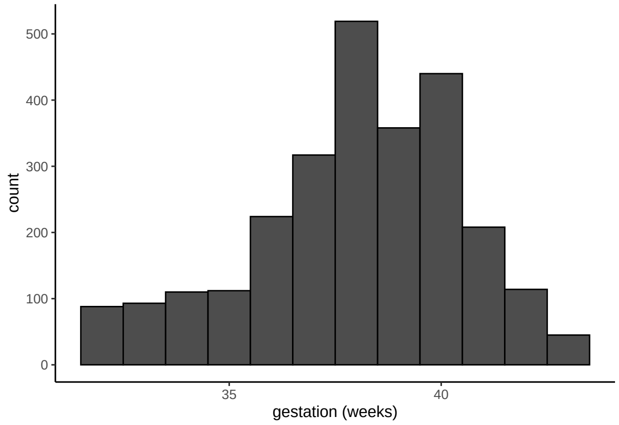


```
##
## $et_bw
```

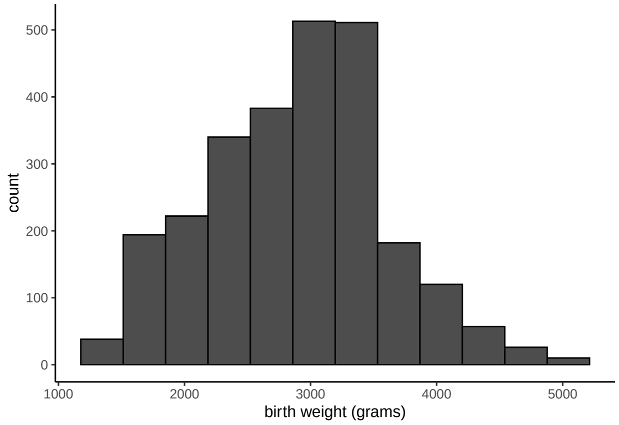


```
##
## $adm_dur
```

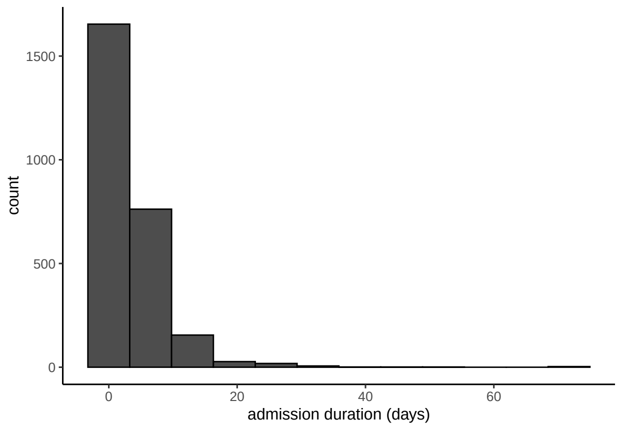


#### $pi_gest

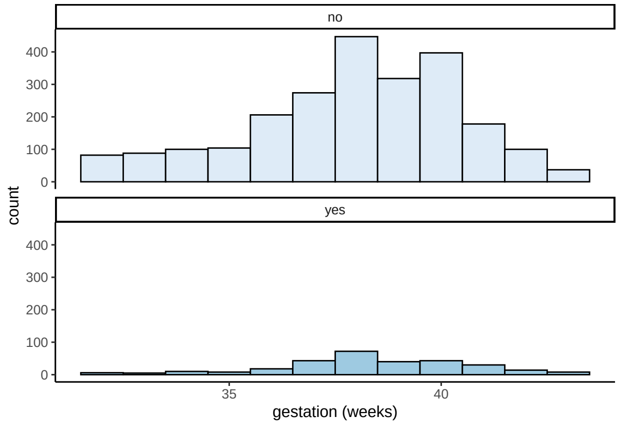


##
## $et_bw

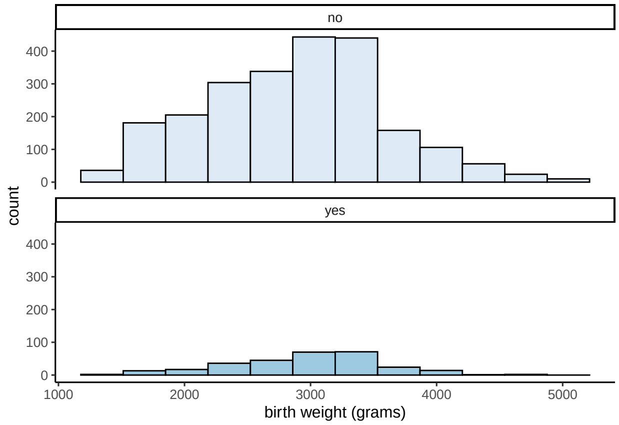


##
#### $adm_dur

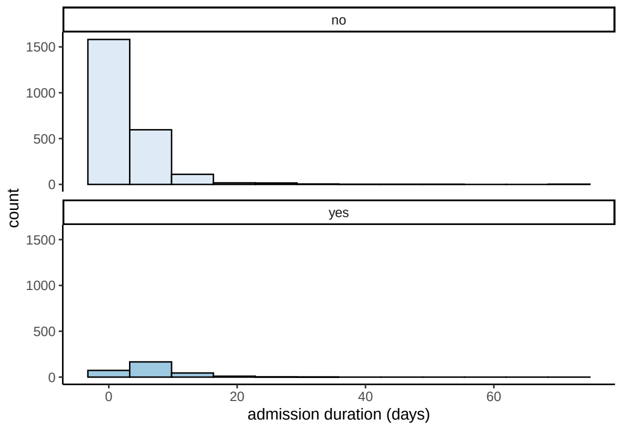


“Gestation” and “birth weight” are approximately normally distributed, while “admission duration” is very right-skewed.

#### 7.2 Table 1

Table summarising the characteristics of included participants:

| Characteristic | Overall, N = 2,628 | no, N = 2,331 | yes, N = 297 | p-value |
|---|---|---|---|---|
| Sex, n (%) | | | | 0.7 |
| f | 1,122 (43%) | 990 (42%) | 132 (44%) | |
| m | 1,503 (57%) | 1,338 (57%) | 165 (56%) | |
| u | 3 (0.1%) | 3 (0.1%) | 0 (0%) | |
| Gestational age, mean weeks (SD) | 38.00 (2.50) | 37.96 (2.52) | 38.36 (2.29) | 0.005 |
| Birth weight, mean grams (SD) | 2,889 (703) | 2,881 (716) | 2,950 (595) | 0.067 |
| Chronological age, n (%) | | | | <0.001 |
| fnb | 1,001 (38%) | 901 (39%) | 100 (34%) | |
| dol1 | 1,257 (48%) | 1,136 (49%) | 121 (41%) | |
| dol2 | 235 (9.0%) | 181 (7.8%) | 54 (18%) | |
| dol3 | 110 (4.2%) | 91 (3.9%) | 19 (6.5%) | |
| Type of birth, n (%) | | | | 0.032 |
| singleton | 2,496 (95%) | 2,205 (95%) | 291 (98%) | |
| twin1 | 127 (4.8%) | 121 (5.2%) | 6 (2.0%) | |
| triplet1 | 2 (<0.1%) | 2 (<0.1%) | 0 (0%) | |
| Mode of delivery, n (%) | | | | 0.074 |
| svd | 1,889 (72%) | 1,663 (71%) | 226 (76%) | |
| electiveCS | 136 (5.2%) | 124 (5.3%) | 12 (4.0%) | |
| emergencyCS | 561 (21%) | 510 (22%) | 51 (17%) | |
| instrumental | 42 (1.6%) | 34 (1.5%) | 8 (2.7%) | |
| Admission duration, median days [Q1-Q3] | 2.3 [1.3-4.9] | 2.1 [1.2-4.1] | 6.0 [3.5-8.8] | <0.001 |
| Death, n (%) | 221 (8.4%) | 184 (7.9%) | 37 (12%) | 0.008 |

p-values are from Welch’s two-sample t-test for gestational age and birth weight; the Wilcoxon-Mann-Whitney U test for admission duration; Pearson’s chi-squared test for age at admission and death; and Fisher’s exact test for sex, type of birth and mode of delivery.

Data are presented for the observed data only (i.e. before MICE) using pairwise deletion of missing values. The number of missing values for each variable in the above table are as follows:

```
## # A tibble: 9 x 3
```

```
##   variable    n_miss pct_miss
##   <chr>        <int>    <dbl>
## 1 et_bw           32    1.22
## 2 pi_age          25    0.951
## 3 pi_type          3    0.114
## 4 pi_sex           0    0
## 5 pi_gest          0    0
## 6 oh_delivery      0    0
## 7 adm_dur          0    0
## 8 outcome          0    0
## 9 sepsis           0    0
```

#### 7.3 Distribution of candidate predictors

| **Characteristic** | **Overall**, N = 2,628 | **no**, N = 2,331 | **yes**, N = 297 | **p-value**[1] |
|---|---|---|---|---|
| pi_gest | 38.00 [37.00-40.00] | 38.00 [37.00-40.00] | 38.00 [37.00-40.00] | 0.032 |
| et_bw | 2,950 [2,400-3,350] | 2,900 [2,400-3,350] | 3,000 [2,600-3,350] | 0.035 |
| oh_matfever | 14 (0.5%) | 8 (0.3%) | 6 (2.0%) | 0.003 |
| oh_offliquor | 163 (6.2%) | 131 (5.6%) | 32 (11%) | 0.001 |
| co_prom | 303 (12%) | 257 (11%) | 46 (15%) | 0.027 |
| et_grunt | 750 (29%) | 654 (28%) | 96 (32%) | 0.13 |
| et_cyanosis | 69 (2.6%) | 60 (2.6%) | 9 (3.0%) | 0.6 |
| et_seizures | 14 (0.5%) | 10 (0.4%) | 4 (1.3%) | 0.064 |
| et_rr | 56 [48-68] | 56 [48-68] | 60 [50-72] | <0.001 |
| et_hr | 138 [126-146] | 138 [126-146] | 139 [127-150] | 0.011 |
| et_temp | 36.50 [36.00-37.00] | 36.50 [36.00-36.90] | 36.90 [36.20-38.00] | <0.001 |
| oe_fontanelle | | | | 0.9 |
| flat | 2,608 (99%) | 2,312 (99%) | 296 (100%) | |
| sunken | 10 (0.4%) | 9 (0.4%) | 1 (0.3%) | |
| bulging | 10 (0.4%) | 10 (0.4%) | 0 (0%) | |
| oe_activity | | | | <0.001 |
| alert | 2,152 (82%) | 1,933 (83%) | 219 (74%) | |
| lethargic | 382 (15%) | 327 (14%) | 55 (19%) | |
| irritable | 62 (2.4%) | 45 (1.9%) | 17 (5.7%) | |
| seizures | 14 (0.5%) | 9 (0.4%) | 5 (1.7%) | |
| coma | 18 (0.7%) | 17 (0.7%) | 1 (0.3%) | |
| oe_nasalflare | 912 (35%) | 791 (34%) | 121 (41%) | 0.023 |
| oe_retractions | 986 (38%) | 848 (36%) | 138 (46%) | <0.001 |
| oe_grunt | 421 (16%) | 360 (15%) | 61 (21%) | 0.029 |
| oe_wob | | | | <0.001 |
| normal | 1,405 (54%) | 1,263 (55%) | 142 (48%) | |
| mild | 413 (16%) | 378 (16%) | 35 (12%) | |
| moderate | 614 (24%) | 529 (23%) | 85 (29%) | |
| severe | 170 (6.5%) | 139 (6.0%) | 31 (11%) | |
| oe_colour | | | | 0.11 |
| pink | 2,507 (95%) | 2,220 (95%) | 287 (97%) | |
| pale | 10 (0.4%) | 7 (0.3%) | 3 (1.0%) | |
| blue | 62 (2.4%) | 58 (2.5%) | 4 (1.3%) | |
| yellow | 49 (1.9%) | 46 (2.0%) | 3 (1.0%) | |
| oe_abdodist | 28 (1.1%) | 26 (1.1%) | 2 (0.7%) | 0.8 |
| oe_omphalitis | 6 (0.2%) | 4 (0.2%) | 2 (0.7%) | 0.14 |
| oe_abskin | 27 (1.0%) | 23 (1.0%) | 4 (1.3%) | 0.5 |

| | | | | |
|---|---|---|---|---|
| hx_vomit | | | | 0.3 |
| no | 2,605 (99%) | 2,309 (99%) | 296 (100%) | |
| yellow | 7 (0.3%) | 7 (0.3%) | 0 (0%) | |
| bilious | 13 (0.5%) | 13 (0.6%) | 0 (0%) | |
| bloody | 3 (0.1%) | 2 (<0.1%) | 1 (0.3%) | |

[1]Data are presented as median [Q1-Q3] for continuous predictors or n (%) for categorical predictors. p-values are from the Wilcoxon-Mann-Whitney U test for continuous predictors and Fisher's exact test for categorical predictors.

Data are presented for the observed data only (i.e. before MICE) using pairwise deletion of missing values. The number of missing values for each variable in the above table are as follows:

```
## # A tibble: 22 x 3
##    variable     n_miss pct_miss
##    <chr>         <int>    <dbl>
##  1 et_temp         814  31.0
##  2 et_bw            32   1.22
##  3 oe_wob           26   0.989
##  4 et_rr            22   0.837
##  5 et_hr             3   0.114
##  6 oe_abdodist       2   0.0761
##  7 oh_matfever       1   0.0381
##  8 oh_offliquor      1   0.0381
##  9 et_grunt          1   0.0381
## 10 et_cyanosis       1   0.0381
## # ... with 12 more rows
```

##### 7.3.1 Box plots of continuous candidate predictors

The box plots below show the distribution of the continuous candidate predictors between participants with and without sepsis.

```
## $pi_gest
```

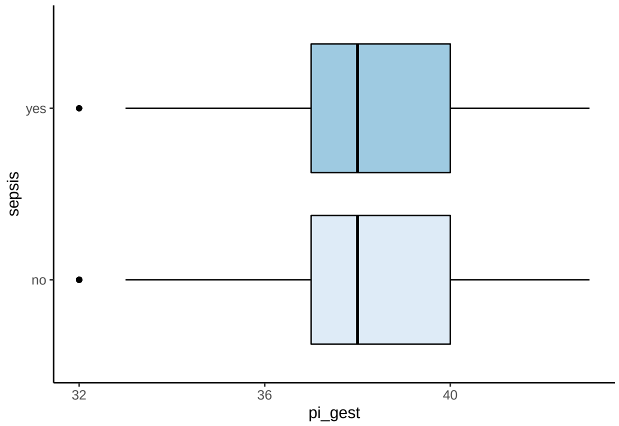


```
##
## $et_bw
```

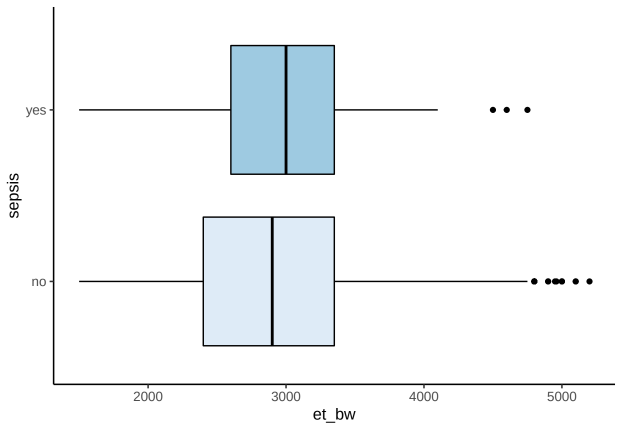


```
##
## $et_rr
```

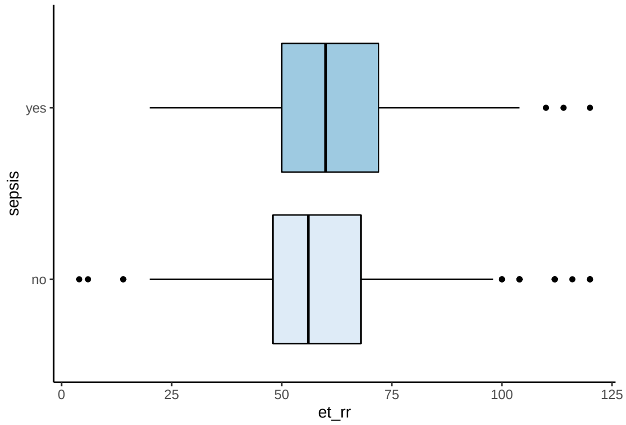


```
##
## $et_hr
```

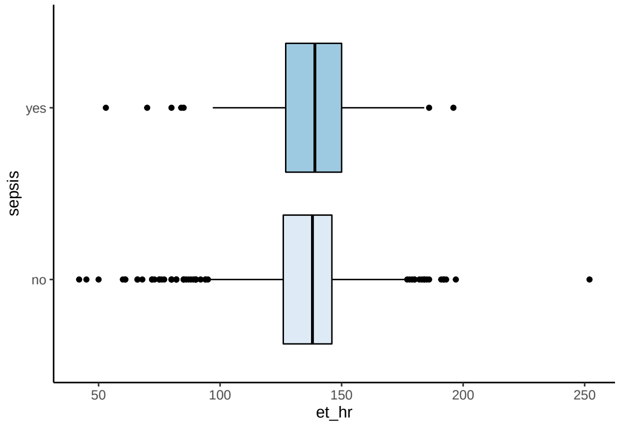


```
##
## $et_temp
```

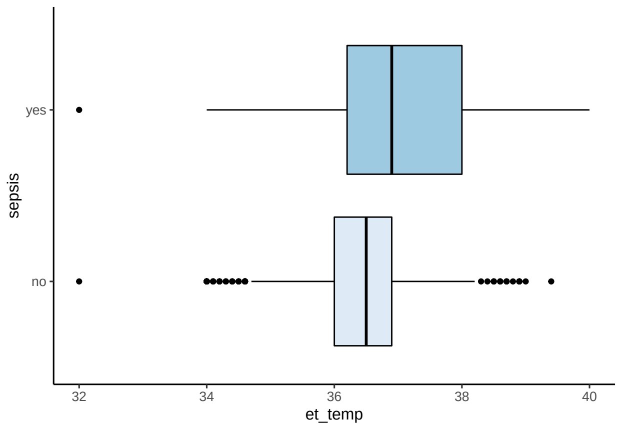
yes
no
sepsis
32
34
36
38
40
et_temp

### 8 Model development and performance

Description of model development.

#### 8.1 Univariable association of candidate predictors with EOS

Below is a univariable logistic regression showing the univariable association between each candidate predictor and the outcome of EOS. The results are pooled across all imputed datasets.

*N.B. To make interpretation easier, birth weight has been converted to kilograms, respiratory rate and heart rate have been divided by 5 (i.e. 5 breaths per minute), and "activity" has been collapsed into "alert", "lethargic", or "other".*

```
## # A tibble: 17 x 7
##    predictor              beta    SE    OR   LCL   UCL     p
##    <fct>                 <dbl> <dbl> <dbl> <dbl> <dbl> <dbl>
##  1 pi_gest               0.067 0.026 1.07  1.02   1.12 0.009
##  2 et_bw                 0.131 0.087 1.14  0.961  1.35 0.133
##  3 oh_matfeveryes        1.79  0.544 5.99  2.06  17.4  0.001
##  4 oh_offliquoryes       0.707 0.208 2.03  1.35   3.05 0.001
##  5 co_promyes            0.391 0.173 1.48  1.05   2.08 0.024
##  6 et_gruntyes           0.203 0.132 1.23  0.945  1.59 0.126
##  7 et_rr                 0.093 0.022 1.10  1.05   1.14 0
##  8 et_hr                 0.047 0.019 1.05  1.01   1.09 0.012
##  9 et_temp               0.886 0.087 2.42  2.04   2.88 0
## 10 oe_activitylethargic  0.395 0.162 1.48  1.08   2.04 0.015
## 11 oe_activityother      1.05  0.25  2.86  1.75   4.67 0
## 12 oe_nasalflareyes      0.29  0.126 1.34  1.04   1.71 0.021
## 13 oe_retractionsyes     0.417 0.124 1.52  1.19   1.93 0.001
## 14 oe_gruntyes           0.346 0.155 1.41  1.04   1.92 0.025
## 15 oe_wobmild           -0.207 0.197 0.813 0.552  1.20 0.293
## 16 oe_wobmoderate        0.345 0.146 1.41  1.06   1.88 0.018
## 17 oe_wobsevere          0.674 0.217 1.96  1.28   3.00 0.002
```

#### 8.2 Model selection

##### 8.2.1 Randomly select a single imputed dataset

To facilitate comparison between models, we randomly select a single imputed dataset (from the 40 imputations) and use this imputation throughout model selection.

```
set.seed(37)
rand <- floor(runif(1, min = 1, max = 30))
rand
```

```
## [1] 16
```

```
si <- as_tibble(complete(imp, rand))
```

##### 8.2.2 Assess linearity assumption

**8.2.2.1 Histograms** We first assessed the linearity assumption – that the outcome of sepsis is modelled by a linear combination of predictors – graphically, by plotting histograms of the proportion of included neonates with sepsis per decile of each continuous predictor.

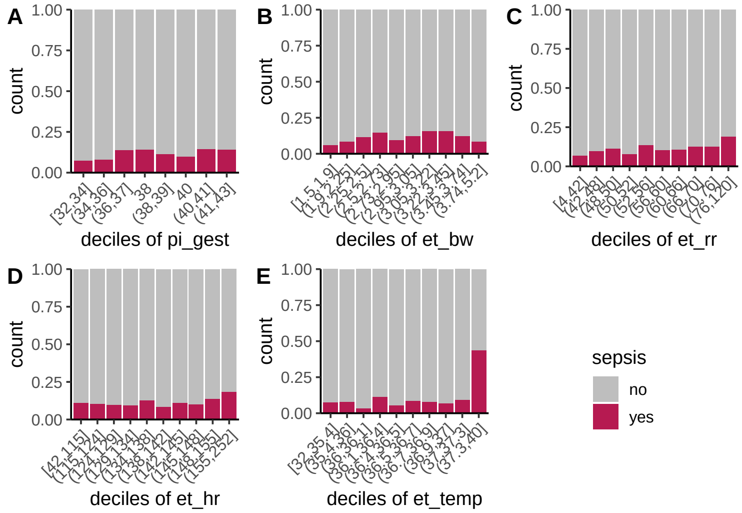


If the relationship between the predictor and the probability of EOS were linear, we would expect the proportion of cases of sepsis to increase or decrease at a constant rate across deciles. Therefore, the above figure suggests some non-linearity for all continuous candidate predictors but most pronounced for temperature.

**8.2.2.2 Splines** We explored non-linear effects of continuous predictors by fitting univariable logistic regression models to predict the outcome of sepsis and modelling each continuous predictor as a natural cubic spline (NCS) function with varying degrees of freedom from 1 (linear) to 10.

We plotted the AIC and BIC of these models for each predictor to visually determine the optimal degrees of freedom for the NCS function.

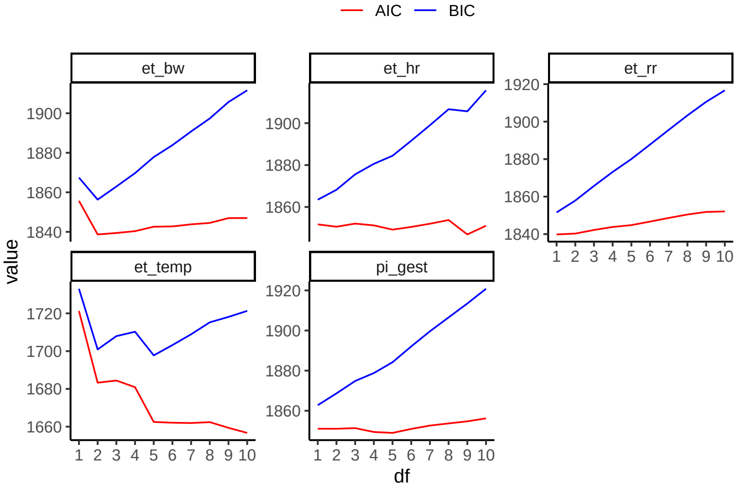


The above figure shows that the AIC and BIC increased monotonically or remained approximately constant across all degrees of freedom for heart rate, respiratory rate and gestational age. This suggests that using the untransformed predictor (i.e. assuming linearity) resulted in a better model than defining these predictors with natural cubic splines.

However, for birth weight, minimum values for AIC and BIC were determined by a natural cubic spline with 2 degrees of freedom (top left panel, above). Similarly, for temperature, the BIC was minimal for natural cubic splines with 2 or 5 degrees of freedom before increasing monotonically. The AIC had minima at 5 or 7 degrees of freedom (bottom left panel, above).

The above figure suggests that transforming birth weight using a natural cubic spline with 2 degrees of freedom and transforming temperature using a natural cubic spline with 5 degrees of freedom produced the optimal univariable models of the natural cubic spline transformations explored.

**8.2.2.3 Polynomials** We further explored non-linear effects by modelling each continuous predictor with polynomial transformations instead of natural cubic spline functions.

Again, we plotted the AIC and BIC of these models for each predictor to visually determine the optimal degree of polynomial.

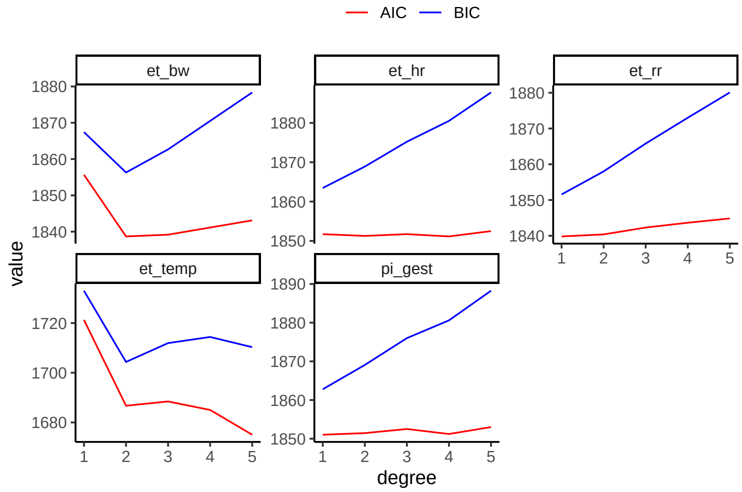


The above figure shows that the AIC and BIC increased monotonically or remained approximately constant across all degrees of polynomials for heart rate, respiratory rate and gestational age. This suggests that using the untransformed predictor (i.e. assuming linearity) resulted in a better model than transforming these predictors with polynomial functions.

However, for birth weight, minimum values for AIC and BIC were determined by a second-degree polynomial (top left panel, above). Similarly, for temperature, the BIC was minimal for a second-degree polynomial and the AIC was minimal for a second-degree or fifth-degree polynomial (bottom left panel, above).

The above figure suggests that transforming birth weight and temperature using a second-degree polynomial produced the optimal univariable models of the polynomial transformations explored.

**8.2.2.4 Univariable models with non-linear transformations - birth weight** Based on the above results, we fit a univariable model to predict early-onset sepsis with birth weight modelled as a natural cubic spline with 2 degrees of freedom.

| **Characteristic** | **log(OR)** | **SE** | **95% CI** | **p-value** |
|---|---|---|---|---|
| ns(et__bw, df = 2) | | | | |
| ns(et__bw, df = 2)1 | 1.5 | 0.468 | 0.62, 2.5 | 0.001 |
| ns(et__bw, df = 2)2 | -1.4 | 0.555 | -2.5, -0.34 | 0.013 |

```
##  50%
## 2.95
```

While both components of the spline were significant, their coefficients were unstable with large SEs.

Thus, we subsequently modelled birth weight as a second-degree polynomial.

| **Characteristic** | **log(OR)** | **SE** | **95% CI** | **p-value** |
|---|---|---|---|---|
| et__bw | | | | |
| et__bw | 5.7 | 3.70 | -1.6, 13 | 0.13 |
| et__bw$^2$ | -16 | 4.03 | -24, -8.4 | <0.001 |

This model suffered similar numerical issues. Adding random noise did not improve estimations in either the natural cubic spline or polynomial models:

| **Characteristic** | **log(OR)** | **SE** | **95% CI** | **p-value** |
|---|---|---|---|---|
| ns(et__bw__noise, df = 2) | | | | |
| ns(et__bw__noise, df = 2)1 | 1.6 | 0.496 | 0.67, 2.6 | 0.001 |
| ns(et__bw__noise, df = 2)2 | -1.4 | 0.569 | -2.5, -0.31 | 0.016 |

| **Characteristic** | **log(OR)** | **SE** | **95% CI** | **p-value** |
|---|---|---|---|---|
| et__bw__noise | | | | |
| et__bw__noise | 5.5 | 3.69 | -1.8, 13 | 0.14 |
| et__bw__noise$^2$ | -16 | 4.02 | -24, -8.2 | <0.001 |

Therefore, birth weight was assumed to be linear in subsequent models.

###### 8.2.2.5 Univariable models with non-linear transformations - temperature

Based on the above results, we fit a univariable model to predict early-onset sepsis with temperature modelled as a natural cubic spline with 5 degrees of freedom and with 2 degrees of freedom.

| **Characteristic** | **log(OR)** | **SE** | **95% CI** | **p-value** |
|---|---|---|---|---|
| ns(et__temp, df = 5) | | | | |
| ns(et__temp, df = 5)1 | -1.1 | 1.24 | -3.4, 1.8 | 0.4 |
| ns(et__temp, df = 5)2 | -2.3 | 1.32 | -4.7, 0.86 | 0.088 |
| ns(et__temp, df = 5)3 | 2.3 | 0.713 | 1.0, 3.9 | 0.001 |
| ns(et__temp, df = 5)4 | -0.93 | 2.80 | -6.2, 5.6 | 0.7 |
| ns(et__temp, df = 5)5 | 3.3 | 0.805 | 1.8, 5.1 | <0.001 |

```
##  20%  40%  60%  80%
## 36.0 36.4 36.7 37.0
```

| **Characteristic** | **log(OR)** | **SE** | **95% CI** | **p-value** |
|---|---|---|---|---|
| ns(et__temp, df = 2) | | | | |
| ns(et__temp, df = 2)1 | -1.4 | 1.36 | -4.0, 1.4 | 0.3 |
| ns(et__temp, df = 2)2 | 5.1 | 0.428 | 4.3, 6.0 | <0.001 |

```
##  50%
## 36.5
```

Similar numerical issues were encountered for these models as were encountered when fitting non-linear functions of birth weight.

Again, we subsequently modelled temperature as a second-degree polynomial and tried adding random noise, neither of which produced satisfactory models.

| **Characteristic** | **log(OR)** | **SE** | **95% CI** | **p-value** |
|---|---|---|---|---|
| et__temp | | | | |
| et__temp | 28 | 3.13 | 22, 34 | <0.001 |
| et__temp$^2$ | 19 | 2.89 | 14, 25 | <0.001 |

| **Characteristic** | **log(OR)** | **SE** | **95% CI** | **p-value** |
|---|---|---|---|---|
| ns(et_temp_noise, df = 2) | | | | |
| ns(et_temp_noise, df = 2)1 | -1.5 | 1.41 | -4.1, 1.5 | 0.3 |
| ns(et_temp_noise, df = 2)2 | 5.2 | 0.436 | 4.3, 6.0 | <0.001 |

| **Characteristic** | **log(OR)** | **SE** | **95% CI** | **p-value** |
|---|---|---|---|---|
| et_temp_noise | | | | |
| et_temp_noise | 27 | 3.13 | 21, 34 | <0.001 |
| et_temp_noise$^2$ | 19 | 2.87 | 13, 24 | <0.001 |

Therefore, temperature was also assumed to be linear in subsequent models.

##### 8.2.3 Selecting main effects

**8.2.3.1 Fit full main effects model (model M1)** We next fit a full main effects model to predict sepsis, including all 14 candidate predictors (those remaining after consideration of skewed predictor distributions). The AIC and BIC of this full model were the benchmark to which subsequent models were compared.

```
## sepsis ~ et_temp + et_rr + et_hr + et_bw + pi_gest + oh_matfever +
##     oh_offliquor + co_prom + et_grunt + oe_activity + oe_nasalflare +
##     oe_retractions + oe_grunt + oe_wob
```

```
##
## Call:
## glm(formula = main_form, family = "binomial", data = si)
##
## Deviance Residuals:
##    Min      1Q  Median      3Q     Max
## -2.005  -0.492  -0.384  -0.276   3.443
##
## Coefficients:
##                       Estimate Std. Error z value Pr(>|z|)
## (Intercept)          -3.90e+01   3.20e+00  -12.18   <2e-16 ***
## et_temp               9.48e-01   8.56e-02   11.08   <2e-16 ***
## et_rr                 6.17e-02   2.71e-02    2.28    0.023 *
## et_hr                -9.88e-04   1.95e-02   -0.05    0.960
## et_bw                -1.28e-01   1.25e-01   -1.03    0.305
## pi_gest               3.79e-02   3.34e-02    1.13    0.256
## oh_matfeveryes        1.47e+00   6.25e-01    2.35    0.019 *
## oh_offliquoryes       5.31e-01   2.28e-01    2.33    0.020 *
## co_promyes            3.67e-01   1.90e-01    1.93    0.054 .
## et_gruntyes          -3.05e-01   2.07e-01   -1.47    0.142
## oe_activitylethargic  4.54e-01   1.90e-01    2.40    0.017 *
## oe_activityother      6.91e-01   2.83e-01    2.44    0.015 *
## oe_nasalflareyes      1.02e-01   2.39e-01    0.43    0.669
## oe_retractionsyes     7.37e-01   3.23e-01    2.28    0.023 *
## oe_gruntyes           1.98e-01   2.19e-01    0.90    0.367
## oe_wobmild           -7.52e-01   3.83e-01   -1.96    0.049 *
## oe_wobmoderate       -2.56e-01   4.18e-01   -0.61    0.541
## oe_wobsevere          1.62e-01   5.08e-01    0.32    0.749
## ---
## Signif. codes:  0 '***' 0.001 '**' 0.01 '*' 0.05 '.' 0.1 ' ' 1
##
## (Dispersion parameter for binomial family taken to be 1)
```

```
##
##     Null deviance: 1854.2  on 2627  degrees of freedom
## Residual deviance: 1632.0  on 2610  degrees of freedom
## AIC: 1668
##
## Number of Fisher Scoring iterations: 5
```

```
##          AIC     BIC
## [1,] 1668.05 1773.78
```

This model assumed linearity of all continuous candidate predictors and additivity at the predictor scale. The regression coefficients and SEs of each predictor in this model (estimated in the single imputed dataset) are as follows:

| **Characteristic** | **OR** | **95% CI** | **p-value** |
|---|---|---|---|
| et_temp | 2.58 | 2.19, 3.06 | <0.001 |
| et_rr | 1.06 | 1.01, 1.12 | 0.023 |
| et_hr | 1.00 | 0.96, 1.04 | >0.9 |
| et_bw | 0.88 | 0.69, 1.12 | 0.3 |
| pi_gest | 1.04 | 0.97, 1.11 | 0.3 |
| oh_matfever | 4.33 | 1.22, 14.6 | 0.019 |
| oh_offliquor | 1.70 | 1.07, 2.63 | 0.020 |
| co_prom | 1.44 | 0.98, 2.08 | 0.054 |
| et_grunt | 0.74 | 0.49, 1.11 | 0.14 |
| oe_activity | | | |
| alert | 1.00 | | |
| lethargic | 1.57 | 1.08, 2.27 | 0.017 |
| other | 2.00 | 1.13, 3.43 | 0.015 |
| oe_nasalflare | 1.11 | 0.70, 1.79 | 0.7 |
| oe_retractions | 2.09 | 1.15, 4.10 | 0.023 |
| oe_grunt | 1.22 | 0.79, 1.87 | 0.4 |
| oe_wob | | | |
| normal | 1.00 | | |
| mild | 0.47 | 0.21, 0.97 | 0.049 |
| moderate | 0.77 | 0.33, 1.71 | 0.5 |
| severe | 1.18 | 0.42, 3.12 | 0.7 |

The highest VIF values were for the 'moderate' and 'severe' categories of work of breathing and retractions. All other VIF values were $< 5$. Pearson's chi-squared test showed that these two predictors were highly correlated with each other:

```
## # A tibble: 17 x 2
##    predictor              VIF
##    <chr>                <dbl>
##  1 oe_wobmoderate        8.26
##  2 oe_retractionsyes     6.06
##  3 oe_wobsevere          5.08
##  4 oe_wobmild            3.79
##  5 oe_nasalflareyes      3.20
##  6 et_gruntyes           2.18
##  7 oe_gruntyes           1.76
##  8 et_bw                 1.63
##  9 pi_gest               1.57
## 10 et_rr                 1.45
```

```
## 11 oe_activitylethargic  1.21
## 12 et_temp               1.21
## 13 oh_offliquoryes       1.09
## 14 co_promyes            1.08
## 15 et_hr                 1.08
## 16 oe_activityother      1.05
## 17 oh_matfeveryes        1.02
```

```
##
##       normal mild moderate severe
##   no    1431  131       77      3
##   yes      0  282      537    167
```

```
##
##  Pearson's Chi-squared test
##
## data:  table(si$oe_retractions, si$oe_wob)
## X-squared = 1947, df = 3, p-value <2e-16
```

**8.2.3.2 Models M2 & M2a** Next, we fit model M2 as the above full model (model M1), but without work of breathing (the predictor with the highest VIF in model M1). This model had a higher AIC compared to model M1, but a lower BIC. Removing work of breathing from the model also reduced collinearity between predictors.

```
## sepsis ~ et_temp + et_rr + et_hr + et_bw + pi_gest + oh_matfever +
##     oh_offliquor + co_prom + et_grunt + oe_activity + oe_nasalflare +
##     oe_retractions + oe_grunt
```

```
##
## Call:
## glm(formula = sepsis ~ et_temp + et_rr + et_hr + et_bw + pi_gest +
##     oh_matfever + oh_offliquor + co_prom + et_grunt + oe_activity +
##     oe_nasalflare + oe_retractions + oe_grunt, family = "binomial",
##     data = si)
##
## Deviance Residuals:
##    Min      1Q  Median      3Q     Max
## -1.934  -0.495  -0.389  -0.280   3.416
##
## Coefficients:
##                      Estimate Std. Error z value Pr(>|z|)
## (Intercept)          -39.0553     3.1937  -12.23   <2e-16 ***
## et_temp                0.9489     0.0852   11.13   <2e-16 ***
## et_rr                  0.0552     0.0267    2.07   0.0386 *
## et_hr                 -0.0024     0.0194   -0.12   0.9019
## et_bw                 -0.1216     0.1245   -0.98   0.3286
## pi_gest                0.0401     0.0333    1.20   0.2296
## oh_matfeveryes         1.3996     0.6140    2.28   0.0226 *
## oh_offliquoryes        0.5146     0.2261    2.28   0.0229 *
## co_promyes             0.3768     0.1895    1.99   0.0468 *
## et_gruntyes           -0.2461     0.1991   -1.24   0.2164
## oe_activitylethargic   0.5402     0.1811    2.98   0.0029 **
```

```
## oe_activityother       0.7470     0.2804    2.66   0.0077 **
## oe_nasalflareyes      -0.0213     0.1877   -0.11   0.9095
## oe_retractionsyes      0.4878     0.2034    2.40   0.0165 *
## oe_gruntyes            0.3449     0.2043    1.69   0.0914 .
## ---
## Signif. codes:  0 '***' 0.001 '**' 0.01 '*' 0.05 '.' 0.1 ' ' 1
##
## (Dispersion parameter for binomial family taken to be 1)
##
##     Null deviance: 1854.2  on 2627  degrees of freedom
## Residual deviance: 1644.3  on 2613  degrees of freedom
## AIC: 1674
##
## Number of Fisher Scoring iterations: 5
```

```
##          AIC     BIC
## [1,] 1674.27 1762.38
```

```
## # A tibble: 14 x 2
##    predictor              VIF
##    <chr>                <dbl>
##  1 oe_retractionsyes     2.42
##  2 et_gruntyes           2.04
##  3 oe_nasalflareyes      2.00
##  4 et_bw                 1.63
##  5 pi_gest               1.57
##  6 oe_gruntyes           1.55
##  7 et_rr                 1.39
##  8 et_temp               1.19
##  9 oe_activitylethargic  1.13
## 10 co_promyes            1.08
## 11 oh_offliquoryes       1.08
## 12 et_hr                 1.08
## 13 oe_activityother      1.04
## 14 oh_matfeveryes        1.02
```

For comparison, model M2a instead dropped retractions from model M1. This model had a slightly improved AIC compared to model M2, but a higher BIC.

```
## sepsis ~ et_temp + et_rr + et_hr + et_bw + pi_gest + oh_matfever +
##     oh_offliquor + co_prom + et_grunt + oe_activity + oe_nasalflare +
##     oe_grunt + oe_wob
```

```
##
## Call:
## glm(formula = sepsis ~ et_temp + et_rr + et_hr + et_bw + pi_gest +
##     oh_matfever + oh_offliquor + co_prom + et_grunt + oe_activity +
##     oe_nasalflare + oe_grunt + oe_wob, family = "binomial", data = si)
##
## Deviance Residuals:
##    Min      1Q  Median      3Q     Max
## -2.001  -0.492  -0.391  -0.279   3.469
##
```

```
## Coefficients:
##                      Estimate Std. Error z value Pr(>|z|)
## (Intercept)          -39.0775     3.2012  -12.21   <2e-16 ***
## et_temp                0.9496     0.0856   11.10   <2e-16 ***
## et_rr                  0.0648     0.0270    2.40    0.016 *
## et_hr                  0.0021     0.0194    0.11    0.914
## et_bw                 -0.1471     0.1244   -1.18    0.237
## pi_gest                0.0375     0.0334    1.12    0.261
## oh_matfeveryes         1.5304     0.6270    2.44    0.015 *
## oh_offliquoryes        0.5260     0.2277    2.31    0.021 *
## co_promyes             0.3758     0.1900    1.98    0.048 *
## et_gruntyes           -0.2214     0.2038   -1.09    0.277
## oe_activitylethargic   0.4658     0.1893    2.46    0.014 *
## oe_activityother       0.7155     0.2821    2.54    0.011 *
## oe_nasalflareyes       0.0261     0.2349    0.11    0.911
## oe_gruntyes            0.1593     0.2172    0.73    0.463
## oe_wobmild            -0.1941     0.2807   -0.69    0.489
## oe_wobmoderate         0.4129     0.2896    1.43    0.154
## oe_wobsevere           0.8900     0.3953    2.25    0.024 *
## ---
## Signif. codes:  0 '***' 0.001 '**' 0.01 '*' 0.05 '.' 0.1 ' ' 1
##
## (Dispersion parameter for binomial family taken to be 1)
##
##     Null deviance: 1854.2  on 2627  degrees of freedom
## Residual deviance: 1637.9  on 2611  degrees of freedom
## AIC: 1672
##
## Number of Fisher Scoring iterations: 5
```

```
##          AIC    BIC
## [1,] 1671.94 1771.8
```

**8.2.3.3 Models M3 & M4** Note that the sign of the regression coefficient for grunting at emergency triage (`et_grunt`) and nasal flaring in model M2 (above) was inconsistent with established subject knowledge of neonatal sepsis. We would expect the presence of these clinical features would increase the probability of sepsis, yet they had negative regression coefficients.

Therefore, model M3 was fitted as model M2, but without grunting at emergency triage or nasal flaring. This model had a slightly lower AIC and BIC compared to model M2.

```
## sepsis ~ et_temp + et_rr + et_hr + et_bw + pi_gest + oh_matfever +
##     oh_offliquor + co_prom + oe_activity + oe_retractions + oe_grunt
```

```
##
## Call:
## glm(formula = sepsis ~ et_temp + et_rr + et_hr + et_bw + pi_gest +
##     oh_matfever + oh_offliquor + co_prom + oe_activity + oe_retractions +
##     oe_grunt, family = "binomial", data = si)
##
## Deviance Residuals:
##    Min      1Q  Median      3Q     Max
## -1.859  -0.497  -0.390  -0.280   3.414
```

```
##
## Coefficients:
##                       Estimate Std. Error z value Pr(>|z|)
## (Intercept)          -39.11391    3.18931  -12.26   <2e-16 ***
## et_temp                0.95273    0.08506   11.20   <2e-16 ***
## et_rr                  0.05134    0.02594    1.98   0.0478 *
## et_hr                 -0.00169    0.01933   -0.09   0.9302
## et_bw                 -0.12153    0.12441   -0.98   0.3286
## pi_gest                0.03810    0.03329    1.14   0.2523
## oh_matfeveryes         1.38993    0.60746    2.29   0.0221 *
## oh_offliquoryes        0.52515    0.22542    2.33   0.0198 *
## co_promyes             0.37897    0.18909    2.00   0.0450 *
## oe_activitylethargic   0.53091    0.18070    2.94   0.0033 **
## oe_activityother       0.73425    0.27960    2.63   0.0086 **
## oe_retractionsyes      0.37793    0.16978    2.23   0.0260 *
## oe_gruntyes            0.23686    0.18293    1.29   0.1954
## ---
## Signif. codes:  0 '***' 0.001 '**' 0.01 '*' 0.05 '.' 0.1 ' ' 1
##
## (Dispersion parameter for binomial family taken to be 1)
##
##     Null deviance: 1854.2  on 2627  degrees of freedom
## Residual deviance: 1645.9  on 2615  degrees of freedom
## AIC: 1672
##
## Number of Fisher Scoring iterations: 5
```

```
##          AIC     BIC
## [1,] 1671.88 1748.24
```

Looking at the above model, the regression coefficient for heart rate was close to zero and it was not found to be a significant predictor in the model. Therefore, heart rate was dropped from model M3 to fit model M4. This model had a lower AIC and BIC compared to model M3. Also, this model had minimal collinearity between predictors.

```
## sepsis ~ et_temp + et_rr + et_bw + pi_gest + oh_matfever + oh_offliquor +
##     co_prom + oe_activity + oe_retractions + oe_grunt
```

```
##
## Call:
## glm(formula = sepsis ~ et_temp + et_rr + et_bw + pi_gest + oh_matfever +
##     oh_offliquor + co_prom + oe_activity + oe_retractions + oe_grunt,
##     family = "binomial", data = si)
##
## Deviance Residuals:
##    Min      1Q  Median      3Q     Max
## -1.854  -0.498  -0.389  -0.280   3.415
##
## Coefficients:
##                      Estimate Std. Error z value Pr(>|z|)
## (Intercept)          -39.1032     3.1868  -12.27   <2e-16 ***
## et_temp                0.9512     0.0833   11.42   <2e-16 ***
## et_rr                  0.0511     0.0258    1.98   0.0478 *
```

```
## et_bw                  -0.1214     0.1244   -0.98   0.3292
## pi_gest                 0.0381     0.0333    1.14   0.2522
## oh_matfeveryes          1.3928     0.6065    2.30   0.0216 *
## oh_offliquoryes         0.5252     0.2254    2.33   0.0198 *
## co_promyes              0.3795     0.1890    2.01   0.0447 *
## oe_activitylethargic    0.5316     0.1805    2.94   0.0032 **
## oe_activityother        0.7347     0.2796    2.63   0.0086 **
## oe_retractionsyes       0.3767     0.1692    2.23   0.0260 *
## oe_gruntyes             0.2371     0.1829    1.30   0.1948
## ---
## Signif. codes:  0 '***' 0.001 '**' 0.01 '*' 0.05 '.' 0.1 ' ' 1
##
## (Dispersion parameter for binomial family taken to be 1)
##
##     Null deviance: 1854.2  on 2627  degrees of freedom
## Residual deviance: 1645.9  on 2616  degrees of freedom
## AIC: 1670
##
## Number of Fisher Scoring iterations: 5
```

```
##          AIC     BIC
## [1,] 1669.89 1740.38
```

```
## # A tibble: 11 x 2
##    predictor              VIF
##    <chr>                <dbl>
##  1 oe_retractionsyes     1.68
##  2 et_bw                 1.62
##  3 pi_gest               1.56
##  4 et_rr                 1.31
##  5 oe_gruntyes           1.25
##  6 et_temp               1.14
##  7 oe_activitylethargic  1.12
##  8 co_promyes            1.08
##  9 oh_offliquoryes       1.08
## 10 oe_activityother      1.04
## 11 oh_matfeveryes        1.01
```

Note that two non-significant predictors were retained in the regression model (premature rupture of membranes and grunting on examination) as the sign of their regression coefficient was consistent with established knowledge and the corresponding *p*-values were reasonably small. Also, birth weight and gestational age were retained in the model despite being non-significant to test for interactions between these two predictors, as described ahead.

##### 8.2.4 Assess additivity assumption

We then assessed the additivity assumption – that the effects of predictors can be added at the linear predictor scale (and thus multiplied at the odds scale) - by assessing for a biologically plausible interaction between birth weight and gestational age.

**8.2.4.1 Interaction plots** There was a significant interaction between birth weight and gestational age in a logistic regression model of these two predictors predicting EOS:

```
## sepsis ~ et_bw * pi_gest

##
## Call:
## glm(formula = sepsis ~ et_bw * pi_gest, family = "binomial",
##     data = si)
##
## Deviance Residuals:
##    Min      1Q  Median      3Q     Max
## -0.693  -0.519  -0.502  -0.425   2.574
##
## Coefficients:
##                Estimate Std. Error z value Pr(>|z|)
## (Intercept)    -17.6505     4.2952   -4.11    4e-05 ***
## et_bw            5.0104     1.5822    3.17  0.00154 **
## pi_gest          0.4110     0.1141    3.60  0.00032 ***
## et_bw:pi_gest   -0.1311     0.0413   -3.17  0.00152 **
## ---
## Signif. codes:  0 '***' 0.001 '**' 0.01 '*' 0.05 '.' 0.1 ' ' 1
##
## (Dispersion parameter for binomial family taken to be 1)
##
##     Null deviance: 1854.2  on 2627  degrees of freedom
## Residual deviance: 1836.2  on 2624  degrees of freedom
## AIC: 1844
##
## Number of Fisher Scoring iterations: 5
```

A plot of this interaction is shown below. Panel A shows the logit of the probability of sepsis across all values of birth weight at six selected values of gestational age. Panel B shows the same interaction but displayed across all values of gestational age at four selected values of birth weight.

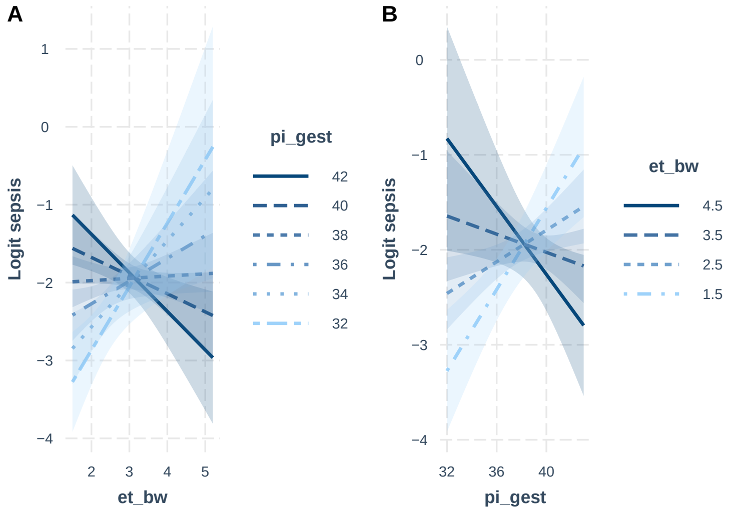


At lower birth weights, those with a higher gestational age appeared to have a greater probability of sepsis compared to those with lower gestational ages (panel A, above). However, at approximately 3200 grams, this relationship reversed, after which the probability of sepsis appeared higher for those with lower gestational ages. The above figure suggests that the probability of sepsis decreased with increasing birth weight for gestational ages > 38 weeks but increased with increasing birth weight for gestational ages < 38 weeks.

This relationship can also be interpreted such that, for lower gestational ages, those with higher birth weights had a greater probability of sepsis compared to those with lower birth weights (panel B, above). For higher gestational ages (above around 38 weeks), those with a higher birth weight had the lowest probability of sepsis.

**8.2.4.2 Models M5 & M5a** The interaction between birth weight and gestational age was included in the selected multivariable model M4 to produce model M5.

The main effects and the interaction term were significant for birth weight and gestational age in this model. However, the coefficients and standard errors were extreme for these terms, with large VIF values.

```
## sepsis ~ et_temp + et_rr + oh_matfever + oh_offliquor + co_prom +
##     oe_activity + oe_retractions + oe_grunt + et_bw + pi_gest +
##     et_bw:pi_gest
```

```
##
## Call:
## glm(formula = sepsis ~ et_temp + et_rr + oh_matfever + oh_offliquor +
##     co_prom + oe_activity + oe_retractions + oe_grunt + et_bw +
##     pi_gest + et_bw:pi_gest, family = "binomial", data = si)
##
## Deviance Residuals:
##    Min      1Q  Median      3Q     Max
## -1.723  -0.496  -0.386  -0.274   3.503
##
## Coefficients:
##                      Estimate Std. Error z value Pr(>|z|)
## (Intercept)          -52.4430     5.5372   -9.47   <2e-16 ***
## et_temp                0.9539     0.0836   11.42   <2e-16 ***
## et_rr                  0.0529     0.0259    2.04   0.0415 *
## oh_matfeveryes         1.3536     0.6044    2.24   0.0251 *
## oh_offliquoryes        0.4682     0.2261    2.07   0.0384 *
## co_promyes             0.3710     0.1895    1.96   0.0503 .
## oe_activitylethargic   0.5568     0.1807    3.08   0.0021 **
## oe_activityother       0.6909     0.2806    2.46   0.0138 *
## oe_retractionsyes      0.4082     0.1696    2.41   0.0161 *
## oe_gruntyes            0.2282     0.1830    1.25   0.2123
## et_bw                  4.9863     1.6892    2.95   0.0032 **
## pi_gest                0.3874     0.1214    3.19   0.0014 **
## et_bw:pi_gest         -0.1338     0.0442   -3.03   0.0025 **
## ---
## Signif. codes:  0 '***' 0.001 '**' 0.01 '*' 0.05 '.' 0.1 ' ' 1
##
## (Dispersion parameter for binomial family taken to be 1)
##
##     Null deviance: 1854.2  on 2627  degrees of freedom
## Residual deviance: 1636.1  on 2615  degrees of freedom
## AIC: 1662
##
## Number of Fisher Scoring iterations: 5
```

```
##          AIC     BIC
## [1,] 1662.07 1738.43
```

```
## # A tibble: 12 x 2
##    predictor                VIF
##    <chr>                  <dbl>
##  1 et_bw:pi_gest        349.
##  2 et_bw                265.
##  3 pi_gest               18.0
##  4 oe_retractionsyes      1.68
##  5 et_rr                  1.31
##  6 oe_gruntyes            1.24
##  7 et_temp                1.14
##  8 oe_activitylethargic   1.12
##  9 oh_offliquoryes        1.08
## 10 co_promyes             1.08
## 11 oe_activityother       1.04
## 12 oh_matfeveryes         1.01
```

Refitting this model but with birth weight and gestational age centred by subtracting their respective sample means from each observation greatly improved the estimates (model M5a). However, the main effects of these terms were no longer significant despite a significant interaction. This is consistent with the crossover interaction effect seen in the interaction plot shown previously.

```
## sepsis ~ et_temp + et_rr + oh_matfever + oh_offliquor + co_prom +
##     oe_activity + oe_retractions + oe_grunt + et_bw_centred +
##     pi_gest_centred + et_bw_centred:pi_gest_centred
```

```
##
## Call:
## glm(formula = sepsis ~ et_temp + et_rr + oh_matfever + oh_offliquor +
##     co_prom + oe_activity + oe_retractions + oe_grunt + et_bw_centred +
##     pi_gest_centred + et_bw_centred:pi_gest_centred, family = "binomial",
##     data = si)
##
## Deviance Residuals:
##    Min      1Q  Median      3Q     Max
## -1.723  -0.496  -0.386  -0.274   3.503
##
## Coefficients:
##                               Estimate Std. Error z value Pr(>|z|)
## (Intercept)                   -38.0075     3.0916  -12.29   <2e-16 ***
## et_temp                         0.9539     0.0835   11.42   <2e-16 ***
## et_rr                           0.0529     0.0259    2.04   0.0415 *
## oh_matfeveryes                  1.3536     0.6045    2.24   0.0251 *
## oh_offliquoryes                 0.4682     0.2261    2.07   0.0384 *
## co_promyes                      0.3710     0.1895    1.96   0.0503 .
## oe_activitylethargic            0.5568     0.1807    3.08   0.0021 **
## oe_activityother                0.6909     0.2806    2.46   0.0138 *
## oe_retractionsyes               0.4082     0.1696    2.41   0.0161 *
## oe_gruntyes                     0.2282     0.1830    1.25   0.2123
## et_bw_centred                  -0.0999     0.1239   -0.81   0.4200
## pi_gest_centred                 0.0010     0.0353    0.03   0.9774
## et_bw_centred:pi_gest_centred  -0.1338     0.0442   -3.03   0.0025 **
## ---
## Signif. codes:  0 '***' 0.001 '**' 0.01 '*' 0.05 '.' 0.1 ' ' 1
```

```
##
## (Dispersion parameter for binomial family taken to be 1)
##
##     Null deviance: 1854.2  on 2627  degrees of freedom
## Residual deviance: 1636.1  on 2615  degrees of freedom
## AIC: 1662
##
## Number of Fisher Scoring iterations: 5
```

```
##          AIC     BIC
## [1,] 1662.07 1738.43
```

```
## # A tibble: 12 x 2
##    predictor                      VIF
##    <chr>                        <dbl>
##  1 oe_retractionsyes             1.68
##  2 pi_gest_centred               1.52
##  3 et_bw_centred                 1.43
##  4 et_rr                         1.31
##  5 oe_gruntyes                   1.24
##  6 et_bw_centred:pi_gest_centred  1.16
##  7 et_temp                       1.14
##  8 oe_activitylethargic          1.12
##  9 oh_offliquoryes               1.08
## 10 co_promyes                    1.08
## 11 oe_activityother              1.04
## 12 oh_matfeveryes                1.01
```

Given that allowing for an interaction between birth weight and gestational age (model M5a) showed only minor improvements in the AIC and BIC compared to the model assuming additivity (model M4), we selected model M4 as it was the simpler model.

This decision was reinforced since the distributions of birth weight and gestational age in our cohort suggested that higher birth weights and gestational ages had a higher probability of sepsis than lower birth weights and gestational ages. This contradicted what is expected from established subject knowledge.

**8.2.4.3 Models M6 and M7** Since the interaction between birth weight and gestational age was no longer included in the model, model M4 was refitted but without gestational age (model M6) as the sign of its regression coefficient contradicted established knowledge and it was not significant in model M4. This improved both the AIC and BIC compared to model M4.

```
## sepsis ~ et_temp + et_rr + et_bw + oh_matfever + oh_offliquor +
##     co_prom + oe_activity + oe_retractions + oe_grunt
```

```
##
## Call:
## glm(formula = sepsis ~ et_temp + et_rr + et_bw + oh_matfever +
##     oh_offliquor + co_prom + oe_activity + oe_retractions + oe_grunt,
##     family = "binomial", data = si)
##
## Deviance Residuals:
##    Min      1Q  Median      3Q     Max
## -1.832  -0.498  -0.392  -0.280   3.398
```

```
##
## Coefficients:
##                      Estimate Std. Error z value Pr(>|z|)
## (Intercept)          -38.0449     3.0407  -12.51   <2e-16 ***
## et_temp                0.9552     0.0832   11.48   <2e-16 ***
## et_rr                  0.0517     0.0258    2.00   0.0452 *
## et_bw                 -0.0386     0.1004   -0.38   0.7004
## oh_matfeveryes         1.3756     0.6051    2.27   0.0230 *
## oh_offliquoryes        0.5378     0.2249    2.39   0.0168 *
## co_promyes             0.3735     0.1891    1.98   0.0482 *
## oe_activitylethargic   0.5410     0.1804    3.00   0.0027 **
## oe_activityother       0.7488     0.2790    2.68   0.0073 **
## oe_retractionsyes      0.3790     0.1693    2.24   0.0252 *
## oe_gruntyes            0.2366     0.1828    1.29   0.1956
## ---
## Signif. codes:  0 '***' 0.001 '**' 0.01 '*' 0.05 '.' 0.1 ' ' 1
##
## (Dispersion parameter for binomial family taken to be 1)
##
##     Null deviance: 1854.2  on 2627  degrees of freedom
## Residual deviance: 1647.2  on 2617  degrees of freedom
## AIC: 1669
##
## Number of Fisher Scoring iterations: 5
```

```
##         AIC     BIC
## [1,] 1669.2 1733.81
```

Finally, in model M7, we refitted model M6 without birth weight as the *p*-value for this term in model M6 was large.

```
## sepsis ~ et_temp + et_rr + oh_matfever + oh_offliquor + co_prom +
##     oe_activity + oe_retractions + oe_grunt
```

```
##
## Call:
## glm(formula = sepsis ~ et_temp + et_rr + oh_matfever + oh_offliquor +
##     co_prom + oe_activity + oe_retractions + oe_grunt, family = "binomial",
##     data = si)
##
## Deviance Residuals:
##    Min      1Q  Median      3Q     Max
## -1.846  -0.497  -0.393  -0.280   3.402
##
## Coefficients:
##                      Estimate Std. Error z value Pr(>|z|)
## (Intercept)          -37.9468     3.0286  -12.53   <2e-16 ***
## et_temp                0.9493     0.0818   11.61   <2e-16 ***
## et_rr                  0.0517     0.0258    2.00   0.0451 *
## oh_matfeveryes         1.3841     0.6051    2.29   0.0222 *
## oh_offliquoryes        0.5327     0.2245    2.37   0.0177 *
## co_promyes             0.3770     0.1889    2.00   0.0459 *
## oe_activitylethargic   0.5410     0.1804    3.00   0.0027 **
```

```
## oe_activityother       0.7446     0.2785    2.67   0.0075 **
## oe_retractionsyes      0.3866     0.1682    2.30   0.0215 *
## oe_gruntyes            0.2364     0.1828    1.29   0.1960
## ---
## Signif. codes:  0 '***' 0.001 '**' 0.01 '*' 0.05 '.' 0.1 ' ' 1
##
## (Dispersion parameter for binomial family taken to be 1)
##
##     Null deviance: 1854.2  on 2627  degrees of freedom
## Residual deviance: 1647.3  on 2618  degrees of freedom
## AIC: 1667
##
## Number of Fisher Scoring iterations: 5
```

```
##          AIC     BIC
## [1,] 1667.35 1726.09
```

```
## # A tibble: 9 x 2
##   predictor              VIF
##   <chr>                <dbl>
## 1 oe_retractionsyes     1.66
## 2 et_rr                 1.31
## 3 oe_gruntyes           1.25
## 4 oe_activitylethargic  1.12
## 5 et_temp               1.10
## 6 co_promyes            1.08
## 7 oh_offliquoryes       1.07
## 8 oe_activityother      1.04
## 9 oh_matfeveryes        1.01
```

##### 8.2.5 Selected model

Model M7 was favoured by both the AIC and BIC and was thus selected as the optimal model. This model included 8 of the 14 candidate predictors. The regression coefficients and SEs of each predictor in this model (estimated in the single imputed dataset) are as follows:

| **Characteristic** | **OR** | **95% CI** | **p-value** |
|---|---|---|---|
| et_temp | 2.58 | 2.21, 3.04 | <0.001 |
| et_rr | 1.05 | 1.00, 1.11 | 0.045 |
| oh_matfever | 3.99 | 1.17, 13.0 | 0.022 |
| oh_offliquor | 1.70 | 1.08, 2.62 | 0.018 |
| co_prom | 1.46 | 1.00, 2.09 | 0.046 |
| oe_activity | | | |
| alert | 1.00 | — | |
| lethargic | 1.72 | 1.20, 2.43 | 0.003 |
| other | 2.11 | 1.20, 3.58 | 0.008 |
| oe_retractions | 1.47 | 1.06, 2.05 | 0.022 |
| oe_grunt | 1.27 | 0.88, 1.81 | 0.2 |

#### 8.3 Model performance

##### 8.3.1 In the single imputed dataset

The ROC curve for the optimal model in the single imputed dataset (imputation number 16) is shown below.

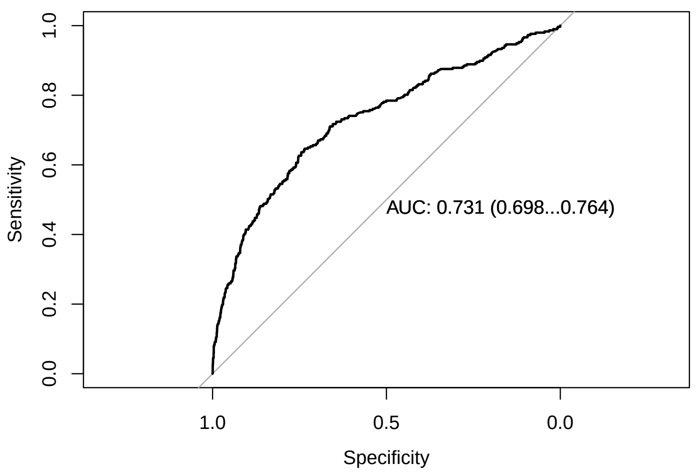


We calculated Yates' discrimination slope as the absolute difference in mean predicted probabilities between the two observed outcome groups. We obtained 95% confidence intervals using bootstrap (calculated using the normal approximation and 10,000 resamples).

```
##
## ORDINARY NONPARAMETRIC BOOTSTRAP
##
##
## Call:
## boot(data = si, statistic = yatesBootstrap, R = 10000, model = M7)
##
##
## Bootstrap Statistics :
##      original     bias    std. error
## t1* 0.105251 0.00581465   0.0178995

## BOOTSTRAP CONFIDENCE INTERVAL CALCULATIONS
## Based on 10000 bootstrap replicates
##
## CALL :
## boot.ci(boot.out = yates, type = c("norm", "perc"))
##
## Intervals :
## Level      Normal             Percentile
## 95%   ( 0.0644,  0.1345 )   ( 0.0781,  0.1476 )
## Calculations and Intervals on Original Scale
```

A boxplot and density plot of predicted probabilities of EOS by observed outcome are shown below. On average, the predicted probability was higher for observed cases of sepsis than observed cases without sepsis. Nevertheless, there was substantial overlap in predicted probabilities, with cases of sepsis with a low predicted probability (below the median for observed cases without sepsis) and cases without sepsis with a high predicted probability (above the median for observed cases with sepsis).

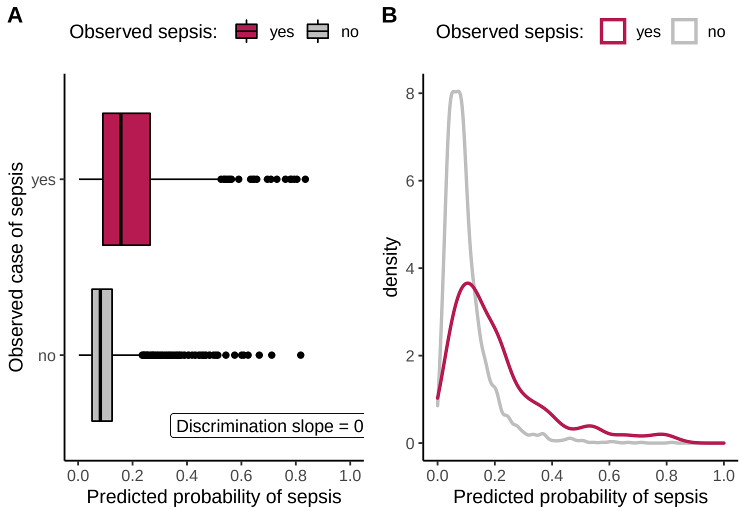


Performance of the optimal model in the selected imputed dataset at various thresholds of predicted probability are shown below. We obtained 95% confidence intervals for likelihood ratios using bootstrap (calculated using the empirical method and 10,000 resamples).

```
## $best
##      obs
## pred   yes   no
##   yes  192  617
##   no   105 1714
##
## $`0.8`
##      obs
## pred   yes   no
##   yes  239 1311
##   no    58 1020
##
## $`0.85`
##      obs
## pred   yes   no
##   yes  251 1448
##   no    46  883
##
## $`0.9`
##      obs
## pred   yes   no
##   yes  268 1816
##   no    29  515
##
## $`0.95`
##      obs
## pred   yes   no
##   yes  283 2068
##   no    14  263

## # A tibble: 5 x 19
```

```
##   thres  sens sens.lcl sens.ucl  spec spec.lcl spec.ucl   PPV PPV.lcl PPV.ucl
##   <dbl> <dbl>    <dbl>    <dbl> <dbl>    <dbl>    <dbl> <dbl>   <dbl>   <dbl>
## 1 0.121  64.6     58.9     70.1  73.5     71.7     75.3  23.7    20.8    26.8
## 2 0.075  80.5     75.5     84.8  43.8     41.7     45.8  15.4    13.7    17.3
## 3 0.067  84.5     79.9     88.4  37.9     35.9     39.9  14.8    13.1    16.6
## 4 0.047  90.2     86.3     93.4  22.1     20.4     23.8  12.9    11.5    14.4
## 5 0.034  95.3     92.2     97.4  11.3     10       12.6  12      10.7    13.4
##     NPV NPV.lcl NPV.ucl   PLR PLR.lcl PLR.ucl   NLR NLR.lcl NLR.ucl
##   <dbl>   <dbl>   <dbl> <dbl>   <dbl>   <dbl> <dbl>   <dbl>   <dbl>
## 1  94.2    93.1    95.3  2.44   1.58     2.88 0.481   0.402   0.595
## 2  94.6    93.1    95.9  1.43   1.04     1.57 0.446   0.378   0.547
## 3  95      93.5    96.4  1.36   1.21     1.59 0.409   0.219   0.476
## 4  94.7    92.4    96.4  1.16   0.9      1.22 0.442   0.334   0.632
## 5  94.9    91.7    97.2  1.07   0.976    1.12 0.418   0.265   0.592
```

The ‘optimal’ classification threshold according to Youden’s $J$ statistic was 0.120643.

##### 8.3.2 Pooled across all imputed datasets

The ROC curve for the optimal model in each of the 40 multiply imputed datasets is shown below.

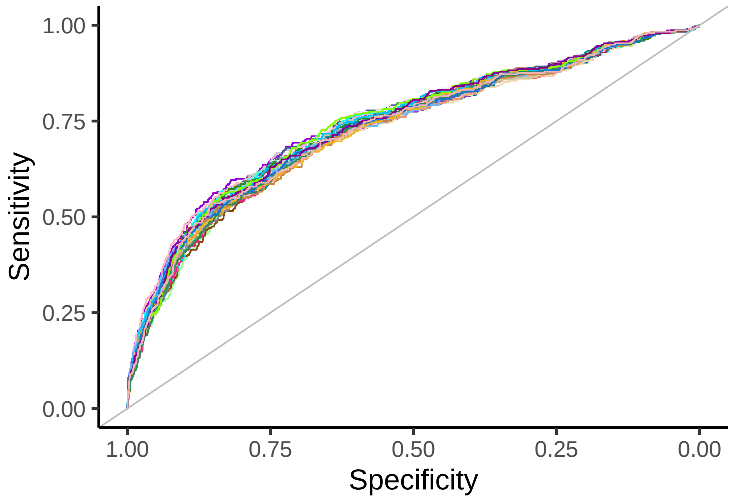


We then applied Rubin’s rules to get the pooled AUC across all imputed datasets.

```
##          auc      lcl      ucl
## [1,] 0.73648 0.700954 0.772006
```

The pooled AUC across the imputed datasets was 0.736 (95% CI 0.701-0.772%).

Finally, we estimated the regression coefficients and odds ratios for the optimal model, pooled across all imputed datasets:

```
## # A tibble: 10 x 7
##    predictor              beta    SE    OR   lcl   ucl p.value
##    <chr>                 <dbl> <dbl> <dbl> <dbl> <dbl>   <dbl>
##  1 (Intercept)          -39.4   3.52  0     0     0       0
##  2 et_temp                0.987 0.095  2.68  2.23  3.23   0
```

```
##  3 et_rr                  0.055 0.026  1.06  1     1.11  0.0373
##  4 oh_matfeveryes         1.44  0.612  4.21  1.27 14.0   0.0189
##  5 oh_offliquoryes        0.543 0.228  1.72  1.1   2.69  0.0174
##  6 co_promyes             0.36  0.192  1.43  0.98  2.09  0.0612
##  7 oe_activitylethargic   0.586 0.184  1.8   1.25  2.58  0.0015
##  8 oe_activityother       0.84  0.286  2.32  1.32  4.06  0.0033
##  9 oe_retractionsyes      0.406 0.172  1.5   1.07  2.1   0.0187
## 10 oe_gruntyes            0.179 0.186  1.2   0.83  1.72  0.337
```
